# Stress about Eviction or Loss of Housing and Child Mental Health

**DOI:** 10.1101/2024.06.28.24309688

**Authors:** Jamie L. Hanson

## Abstract

**Importance:** Eviction and housing loss are pressing public health concerns. Understanding how caregivers’ stress about eviction or loss of housing relates to specific childhood psychiatric issues across development is important.

**Objective:** To examine associations between stress about eviction or loss of housing and caregiver-reported child depression, anxiety, attention-deficit/hyperactivity disorder (ADHD), and behavioral problems, adjusting for sociodemographic factors.

**Design:** Cross-sectional analysis of a nationally representative survey, collected between July 2022 to January 2023.

**Setting:** United States.

**Participants:** Over 36,000 caregivers and children from a national survey.

**Main Outcomes and Measures:** Caregiver-reported child depression, anxiety, ADHD, and behavioral problems. Generalized linear mixed models were used to test associations with stress about eviction or loss of housing.

**Results:** Analyses from a sociodemographically diverse sample of 36,710 children indicated that stress about eviction or loss of housing was associated with 4-35% increased odds of internalizing psychopathology (i.e., depression and anxiety) in children. Relations were sometimes moderated by age, with stronger associations for younger children. No consistent relations emerged between stress about eviction or loss of housing and ADHD or behavioral problems after adjustment.

**Conclusions and Relevance:** This study provided new insights into how stress about eviction or loss of housing is differentially associated with internalizing versus externalizing psychopathology across child development. Prospective longitudinal research is still needed to fully understand these complex relations over time. Findings underscored the importance of policies and interventions to address housing instability and its mental health consequences for children.

**Key Points:** *Question:* What are the associations between caregivers’ stress about eviction or housing loss and children’s mental health outcomes across different ages?

*Findings:* In this cross-sectional study of 36,638 children, stress about eviction or housing loss was associated with increased odds of internalizing symptoms (e.g., depression; anxiety), with stronger relations seen for depression in younger children. No consistent associations were found with ADHD or behavioral problems after adjusting for different sociodemographic factors.

*Meaning:* Stress about eviction or housing loss may differentially impact children’s mental health outcomes, particularly internalizing symptoms in younger children. This underscores the importance of housing stability interventions for child mental health.

---

Eviction, foreclosures and housing instability presents significant threats to the well-being of millions of Americans each year^1,2^. Millions of families face evictions, removal from their homes^3^, or foreclosures^4^. This is of incredible import for public health and public policy, as adults under threat of eviction or foreclosure report and evince multiple negative physical and mental health outcomes^2,5,6^. Notably, eviction and foreclosures are not equally distributed across demographics– it disproportionately affects communities of color, the economically marginalized, and families with children in their home^7^. Despite these alarming facts, there are many open questions related to the impacts of eviction, foreclosures and housing loss on child mental health where deeper investigation is needed, especially regarding the etiology and occurrence of different psychiatric issues.

Research finds that eviction, foreclosures, and housing instability have significant health impacts on families and children, including premature birth, maternal depression, and parenting stress^8–11^. Housing loss and eviction have been linked to decreased social support^12^, food insecurity^13,14^, increased conflict in the home^15^, and harsh parenting^16–20^. These challenges may cascade to significantly impact child development and mental health^21–24^. For example, housing instability and experiencing multiple moves in childhood is associated with more externalizing problems^25^ – or aggression, rule-breaking, and other disruptive behaviors that are expressed outwardly through actions^26^. Similarly, household and residential changes in adolescence were related to depression and internalizing problems, or issues that are inwardly directed, with distress in internal thoughts and feelings^27^. Examined collectively, research generally suggests housing instability impacts child mental health, but further study is needed to understand the consequences of these experiences on child and adolescent functioning.

While existing research has provided insights into links between housing instability and child mental health, important gaps remain in fully understanding these relations. First and foremost, while individuals and families can be formally evicted via court order, informal evictions are much more common. Informal eviction is the most common reason (∼72%) for a forced move^28^, as landlords may pressure, intimidate, or deceive a renter to leave without court proceedings^29^. While foreclosures happen through court orders, there may be anticipatory shame, loss, and regret occurring as a borrow starts defaulting^30–32^. As such, it is important to think about the psychological pressures and concerns related to eviction or foreclosure to more fully understand these issues. Second, additional work is needed to examine eviction’s or foreclosure’s associations with mental health in relation to children’s age. Eviction or loss of housing at specific points in development could have larger impacts on health; for example, younger children may be more impacted by stress and experiences as: they are more dependent on caregivers and substantial brain development is occurring earlier in time^33,34^. Age may moderate the impact of eviction or foreclosures given that different forms of psychopathology show differential onsets in childhood versus adolescence. Finally, there is significant variability in how the impacts of eviction or foreclosures are isolated, and what confounding variables are used in adjusting statistical models. Nearly all projects include child demographics, while only some projects adjust for other developmental challenges common to families who face eviction (i.e., food insecurity; low birth weight). Moving forward, it will be critical to think about psychological pressures and anxiety related to eviction or loss of housing, while comprehensively adjusting statistical models and considering potential interactions with age.

To overcome these limitations, in this study, we used a national survey to test associations between caregivers’ stress about being evicted or housing loss and indicators of four forms of psychopathology– depression, anxiety, attention-deficit/hyperactivity disorder (ADHD), behavioral/conduct problems. We were interested in the main effect of stress about eviction or loss of housing; and the interactions between stress about eviction or loss of housing and child age to potentially understand differential developmental impacts of mental health. Given the wide-reaching impacts of eviction, we hypothesized: 1) Caregiver stress about eviction or loss of housing will be positively associated with risk for these four types of mental health problems; and 2) the association between caregiver stress about eviction or loss of housing and child mental health problems will be moderated by age, with greater relations seen in younger children.

## METHOD

### Participants

We leveraged the 2022 National Survey of Children’s Health (NSCH), an ongoing population-based survey that collected responses from over 50,000 caregivers in the United States^35,36^. Administered by the U.S. Census Bureau, the NSCH is a cross-sectional, high-quality survey and was collected across all 50 U.S. states between July 2022 and January 2023. The survey collected data from 54,103 households (∼1,061 per state). Parents or other knowledgeable adult caregivers answered questions about one specific child (0–17 years old) under their care. These caregivers completed a large battery of questions about their child, their demographics, their family circumstances, and multiple other domains.

### Measures

To measure stress about eviction or loss of housing, caregivers were asked one question about how often they were worried or stressed about being evicted, foreclosed on, or having their house condemned during the past year. Responses included “*Always, Usually, Sometimes, Rarely, Never*”. We reverse the coding of this item, so that higher scores indicated greater concern and stress about these issues. Related to mental health, we focused on caregiver reports of whether their children currently had, or they were previously told that their children had the following conditions: depression, anxiety, ADHD, or behavioral/conduct problems. We recoded these questions to be a binary indicator, with 1=“Currently has condition” and 0=“Does not have condition” or “Ever told, but does not currently have condition”.

Regarding statistical covariates, we adjusted models for a basic set of demographic factors, including child sex, child age, child race/ethnicity, family structure, caregiver’s highest education level, and household poverty level based on federal guidelines and caregiver-reported family income (as noted in Table 1). Race/ethnicity was based on parental self-report data on the selected child’s Hispanic origin and race, with imputation for missing cases. We also more stringently adjusted our statistical models for factors correlated with, but independent of, eviction stress. These included: the number of places the child lived in the past year, if the child ever experienced homelessness, premature birth, low birth weight, poor maternal mental health, food insecurity over the past year, and exposure to adverse childhood experiences. We employed these two sets of covariates motivated to balance under- and over-adjustment biases. While it is critical to rule out potential confounding variables, “overadjustment” in social science can skew estimates away from the true total causal effect, prevent consistent estimation of associations, and introduce additional errors (e.g., collider-stratification)^37,38^.

**Table 1.**
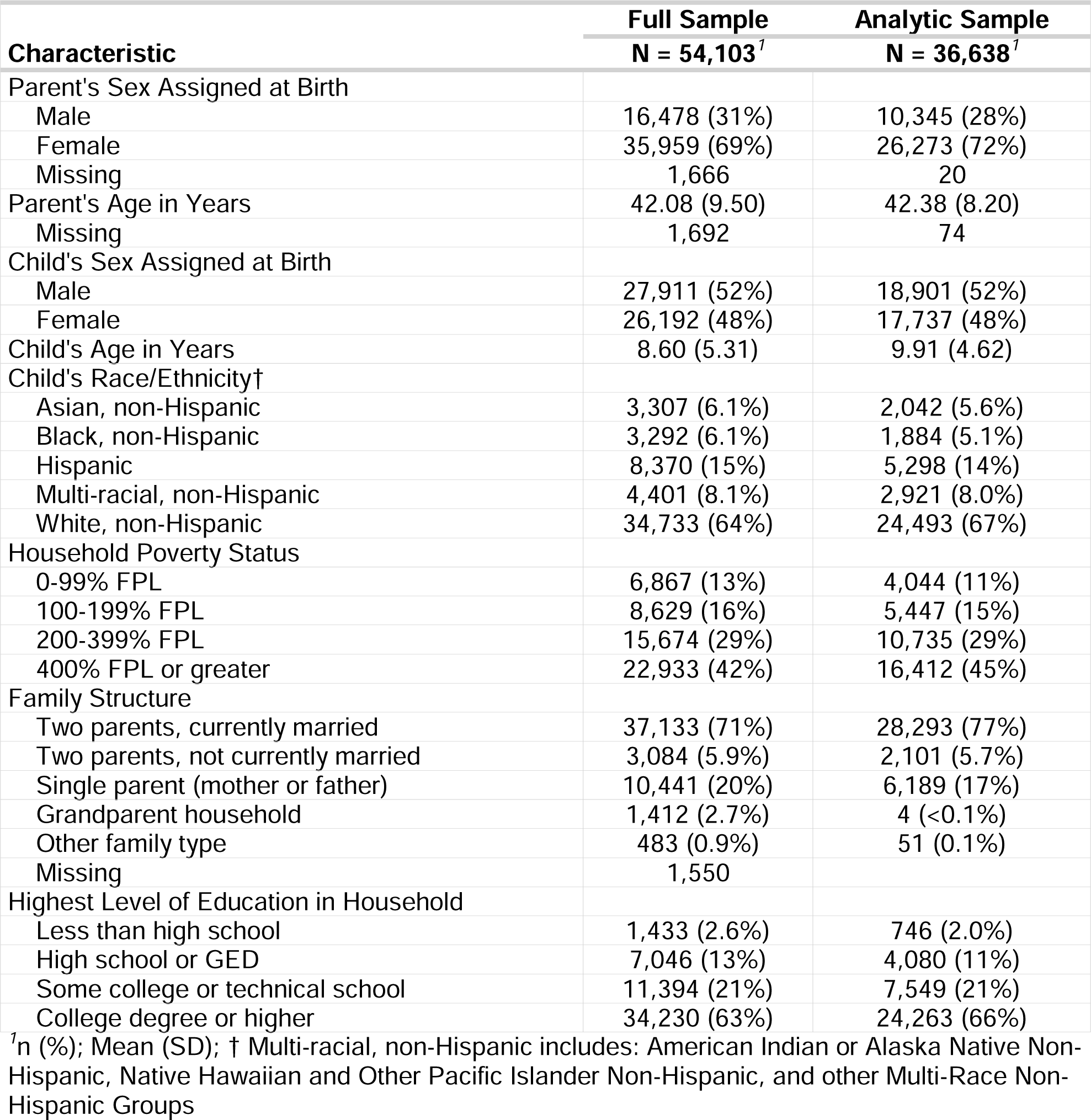
Descriptive statistics of NSCH variables for full and analytic samples used in the project.

This study adheres to the Strengthening the Reporting of Observational Studies in Epidemiology (STROBE) reporting guideline for observational studies^39^. Of note, this work did not meet the definition of human research and was, therefore, “exempt” from Institutional Review Board oversight at the University of Pittsburgh.

### Statistical Analyses

We ran separate generalized linear mixed models with a logistic link function, where each form of psychopathology (a binary indicator) was entered as the dependent variable; in base or stringently adjusted models, eviction stress and different covariates were entered as independent variables, and we included a random factor for geographic location (50 US states, plus the District of Columbia). These multiple statistical adjustments were a way to address potential sources of bias. For parsimony, we included the interaction between stress about eviction or loss of housing and age in all models. We calculated adjusted odds ratios and 95% confidence intervals (CI) for our independent variables of interest (stress about eviction or loss of housing; stress about eviction or loss of housing X age). For significant interactions, we tested differences for the simple slopes of the association between stress about eviction or loss of housing at different values of child age in relation to each form of psychopathology. Analyses were conducted using R version 4.4.0^40,41^ and only included participants with complete data for all variables across the two sets of covariates (*Analytic N=36638*).

## RESULTS

### Descriptive Statistics of the Sample

The full NSCH sample included 54,103 families, while the analytic sample included 36,638 families. Both samples were predominantly White, non-Hispanic, followed by Hispanic. Black, non-Hispanic and Asian, non-Hispanic participants comprised a smaller portion of the samples. The average age of the full sample was 8.6 years and for the analytic sample was 9.9 years.

Most participants in both samples came from families with two parents who were currently married. The highest level of education in the household for most participants was a college degree or higher. These demographic statistics are noted in Table 1.

### Models Examining the Presence of Depression or Anxiety

Our statistical models with a base set of covariates found several factors significantly associated with the presence of depression, including family structure, race/ethnicity, caregiver education, poverty status, and sex assigned at birth (as noted on the left side of Table 2). Stress about eviction or loss of housing was significantly associated with higher odds of current depression, with an adjusted OR of 1.35 [CI=1.27-1.44], z= 9.348, p<.001. The interaction of stress about eviction or loss of housing X age was significant (z=-2.415, p=0.016). The slope for stress about eviction or loss of housing was continually significant, but varied, at different levels of age (at lower levels of age −1 SD, CI=0.26-0.48, z=6.45, p<.001; at higher levels of age +1 SD CI=0.19-0.27, z=11.38, p<.001). More stringent model adjustment attenuated some of these connections, as shown on the right side of Table 2. Stress about eviction or loss of housing was again significantly associated with higher odds of depression, with an adjusted OR of 1.10 [CI=1.02-1.18], z=2.650, p=0.008. The interaction of stress about eviction or loss of housing X age was significant (z=-2.505, p=0.012). For these models, the slope for stress about eviction or loss of housing was significant in younger participants, but not for older participants (−1SD age CI=0.05-0.29, z=2.76, p=0.006; at higher levels of age +1 SD CI= −0.03-0.06 z=0.76, p=0.445). For graphically depiction, please see our *Supplemental Materials*.

**Table 2.**
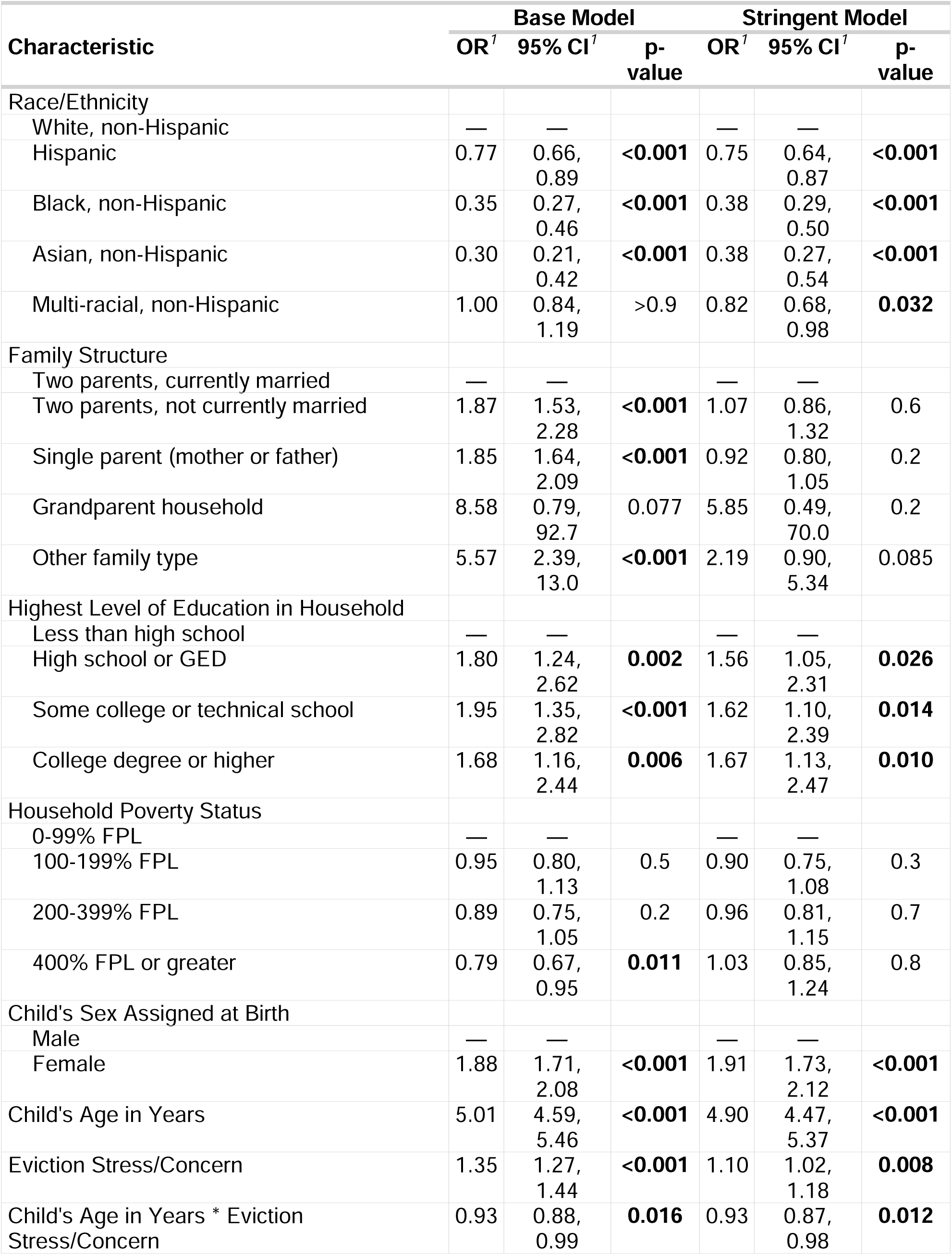
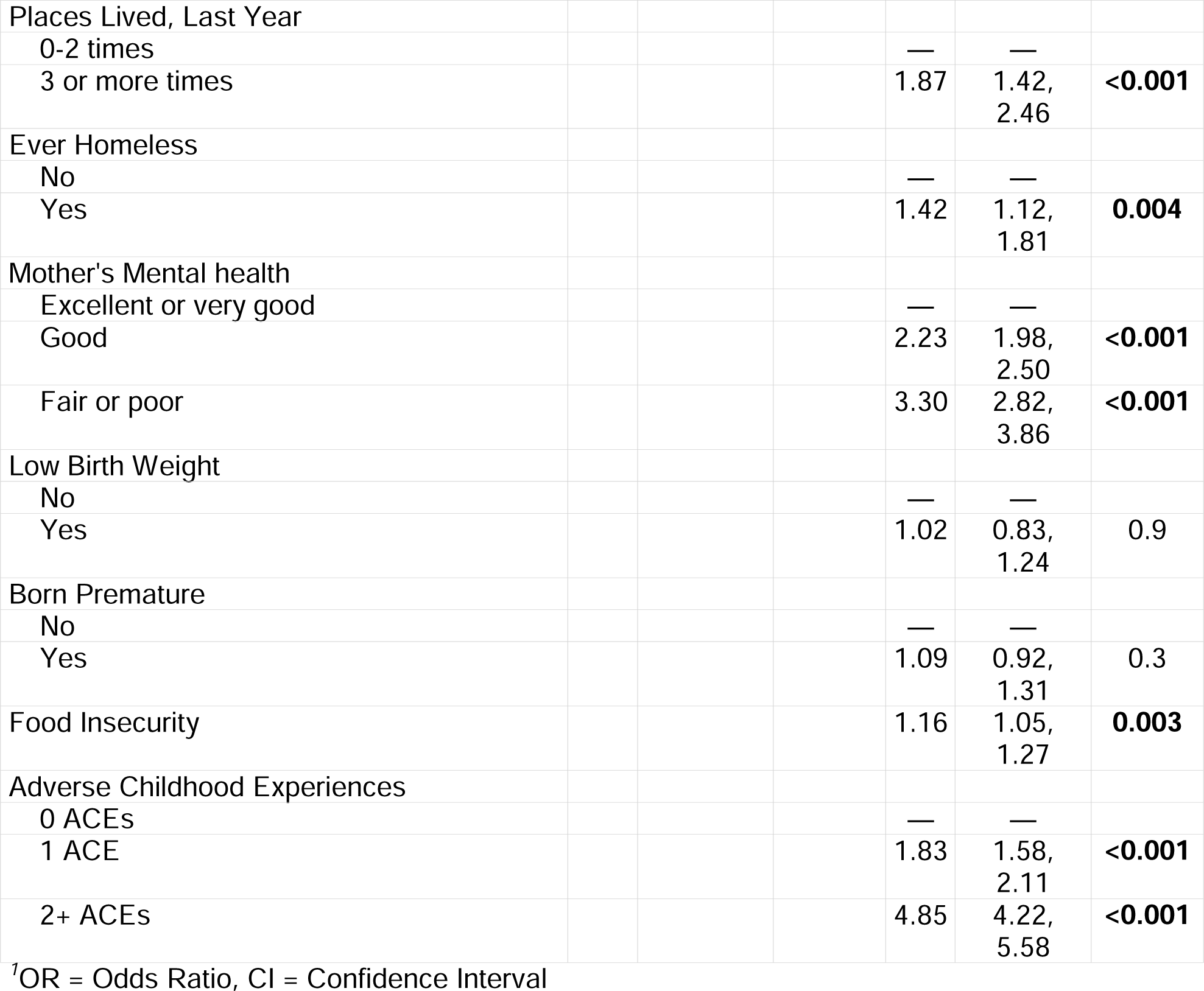
Odds ratios from generalized linear mixed models examining associations between caregiver eviction stress and child depression. The left side of the table shows adjustment using a base set of covariates, while the right side has a more stringent set of model adjustments/covariates.

Like depression, the base adjusted model found anxiety was related to many sociodemographic factors (as noted on the left side of Table 3). Connected to our primary hypotheses, greater reported stress about eviction or loss of housing was significantly associated with higher odds of reported anxiety, with an adjusted OR of 1.26 [CI=1.22-1.31], z=12.792, p<.001. The interaction of stress about eviction or loss of housing X age was significant (z= −2.030, p=0.042). Again, the slope for stress about eviction or loss of housing was continually significant, but varied, at different levels of age (at lower levels of age −1 SD, CI=0.21-0.33, z=8.46, p<.001; at higher levels of age +1 SD CI=0.16-0.23, z=11.71 p<.001). More stringent model adjustment attenuated relations between anxiety and sociodemographic factors (as noted on the right side of Table 3). Diverging slightly from results with depression, in stringently adjusted models, stress about eviction or loss of housing was not significantly associated with higher odds of anxiety, with an adjusted OR of 1.04 [CI=1.00-1.08], z= 1.879, p=0.06. The interaction of stress about eviction or loss of housing X age was not significant (z= −1.881, p=0.06).

**Table 3.**
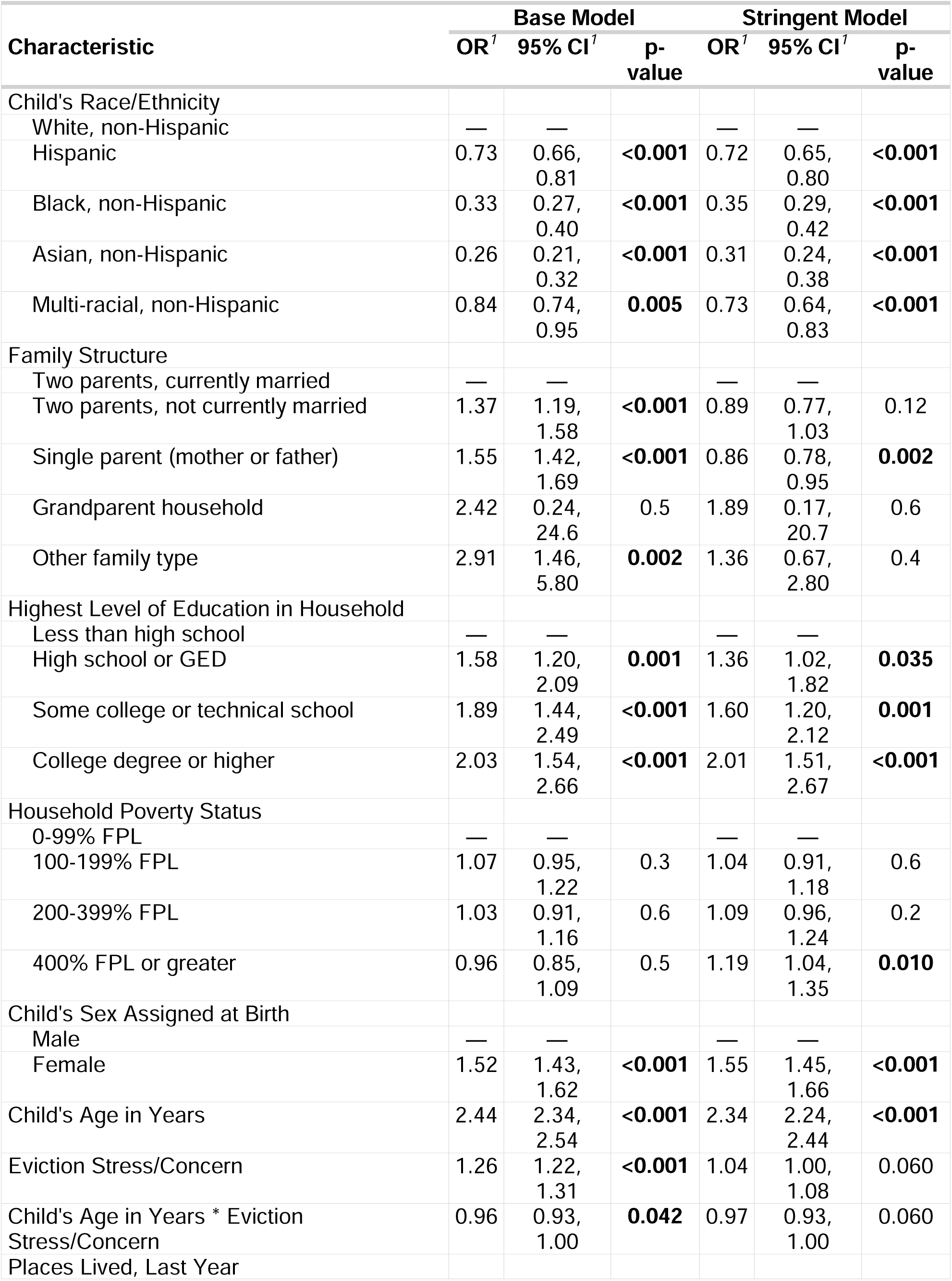
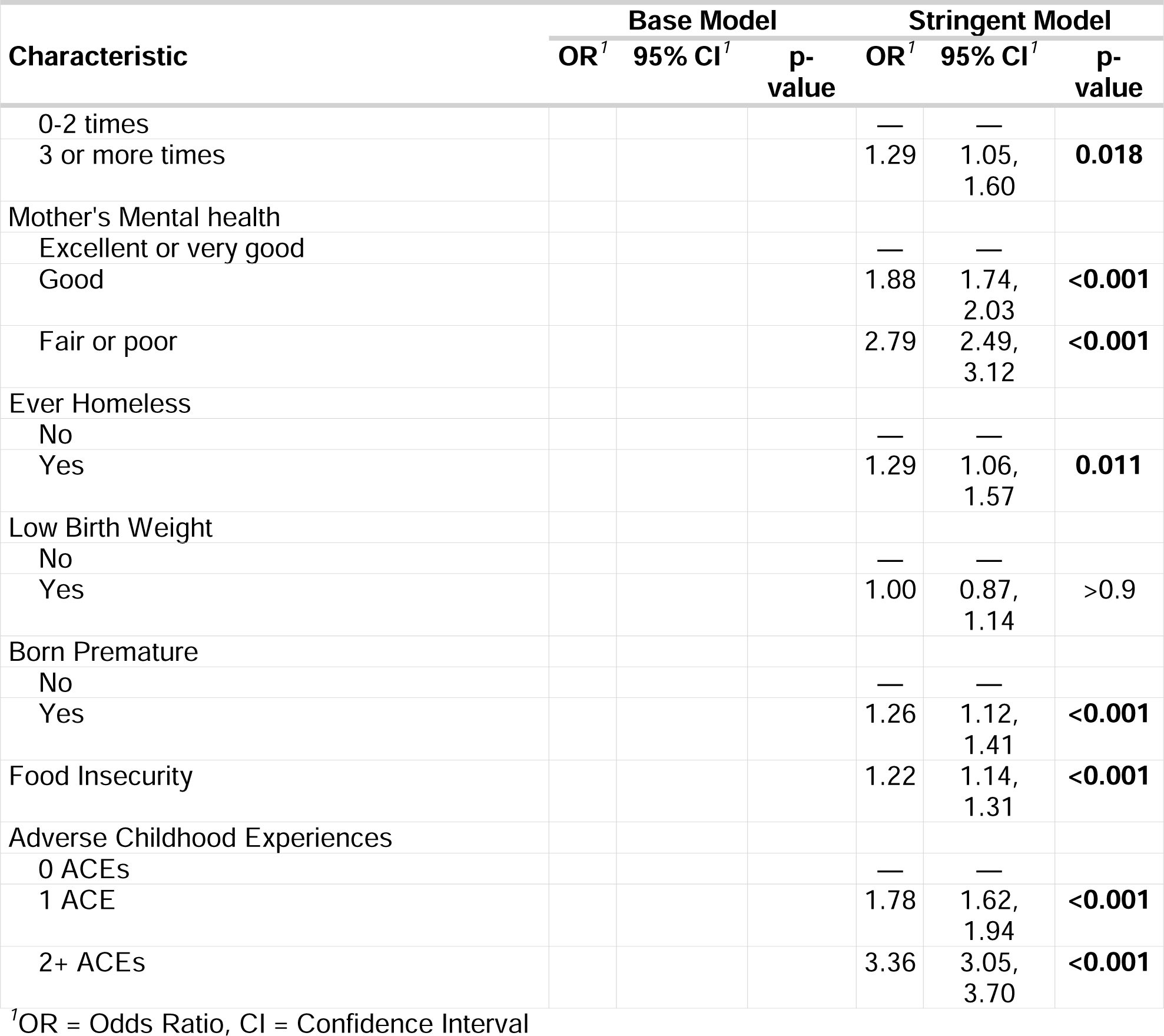
Odds ratios from generalized linear mixed models examining associations between caregiver eviction stress and child anxiety. The left side of the table shows adjustment using a base set of covariates, while the right side has a more stringent set of model adjustments/covariates.

### Models Examining the Presence of ADHD or Behavioral Problems

Focusing on ADHD, we see increased incidence of this disorder was related to family structure, level of caregiver education, poverty status, and sex assigned at birth (see Table 4, left side). Stress about eviction or loss of housing was related to greater rates of ADHD, OR=1.19 [CI=1.15-1.23], z=9.519, p < 0.001. The interaction of stress about eviction or loss of housing X age was not significant (z=-1.670, p=0.09). When adjusting for a more stringent set of covariates, there were connections between ADHD and different covariates (e.g., having low birthweight, childhood adversity, see Table 4, right side). Of important note, in these models, ADHD was not related to stress about eviction or loss of housing (OR=1.03, CI=0.99-1.07, z=1.441, p=0.15) or the interaction of stress about eviction or loss of housing X age (z=-1.486, p=0.14).

**Table 4.**
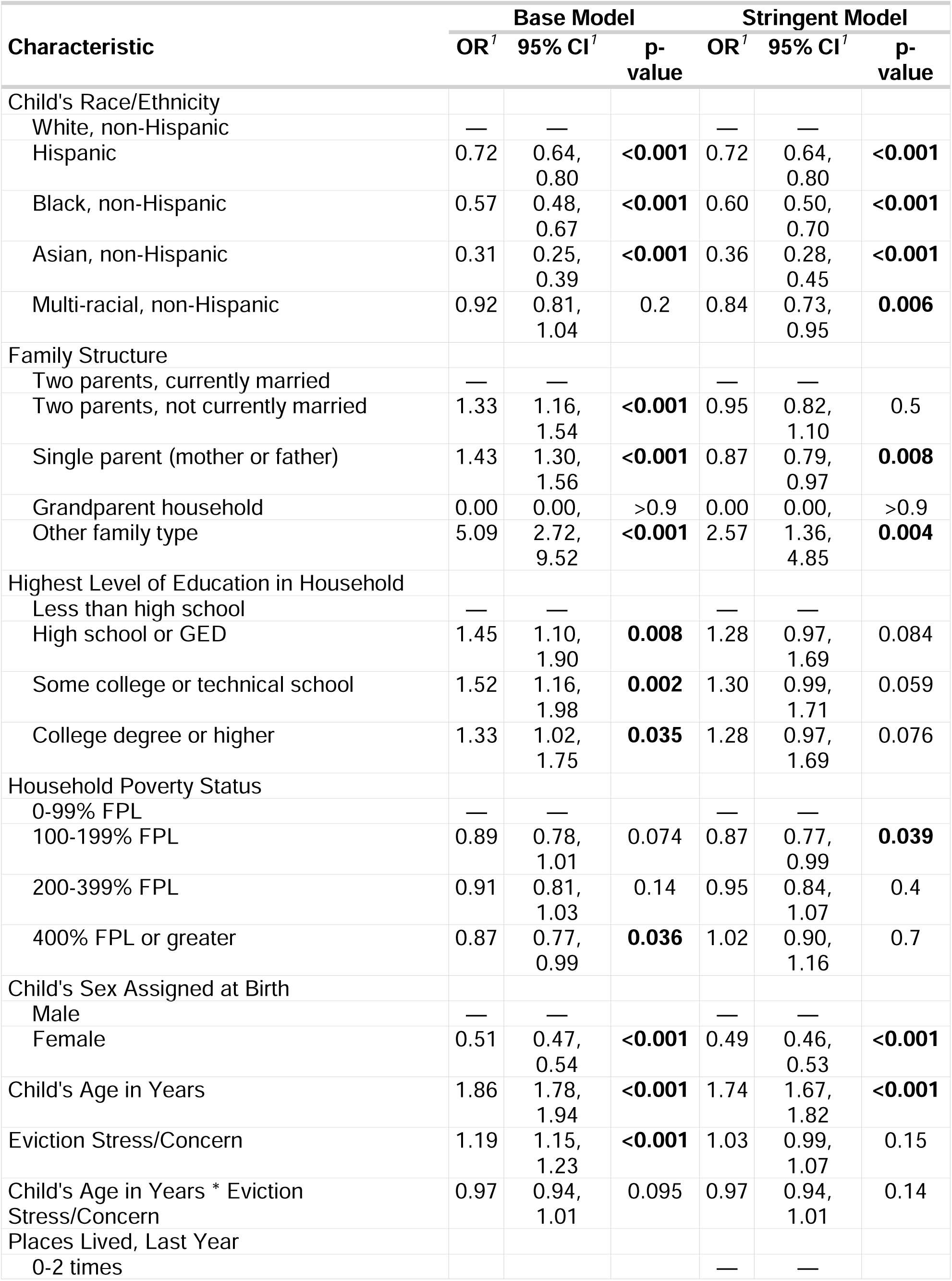
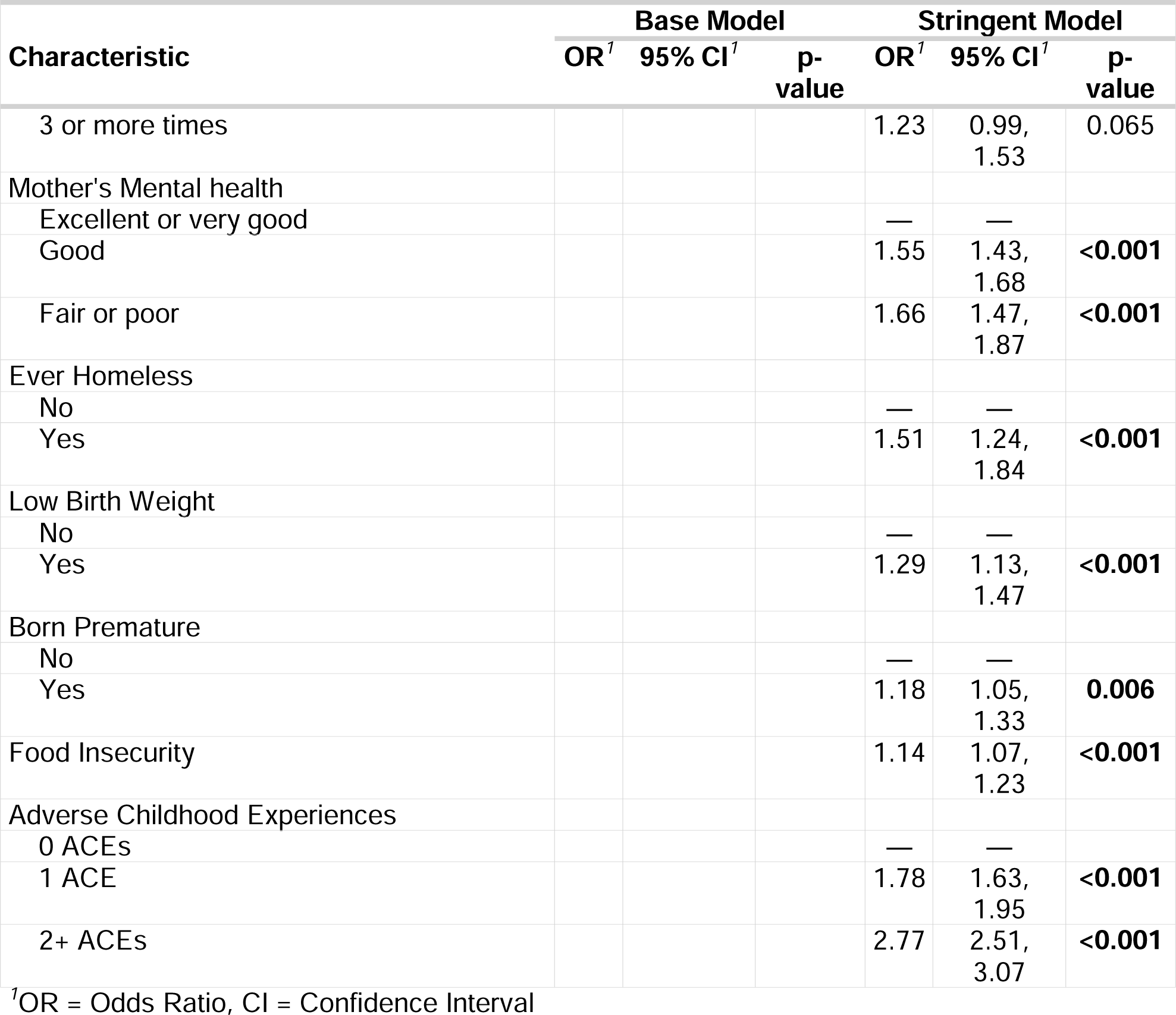
Odds ratios from generalized linear mixed models examining associations between caregiver eviction stress and child ADHD. The left side of the table shows adjustment using a base set of covariates, while the right side has a more stringent set of model adjustments/covariates.

Finally, regarding behavioral/conduct problems, in our base adjusted models, we see increased incidence of this disorder was related to race/ethnicity, family structure, and some indices of socioeconomic status (as noted on the right side of Table 5). Stress about eviction or loss of housing was related to greater rates of behavioral/conduct problems, OR=1.22 [CI=1.18-1.26], z=11.151, p < 0.001. The interaction of stress about eviction or loss of housing X age was not significant (z=-0.175, p=0.86). Stringent adjustment did not change most of these associations, but there were significant connections between behavioral/conduct problems and different covariates (e.g., low birth weight, premature birth, food insecurity, childhood adversity, as shown on the right side of Table 5). Again, of important note and in contrast to results for internalizing issues, behavioral/conduct problems were not related to stress about eviction or loss of housing (OR=1.00, CI: 0.96-1.03, z=0.122, p=0.90) or the interaction of stress about eviction or loss of housing X age (z=0.158, p=0.87).

**Table 5.**
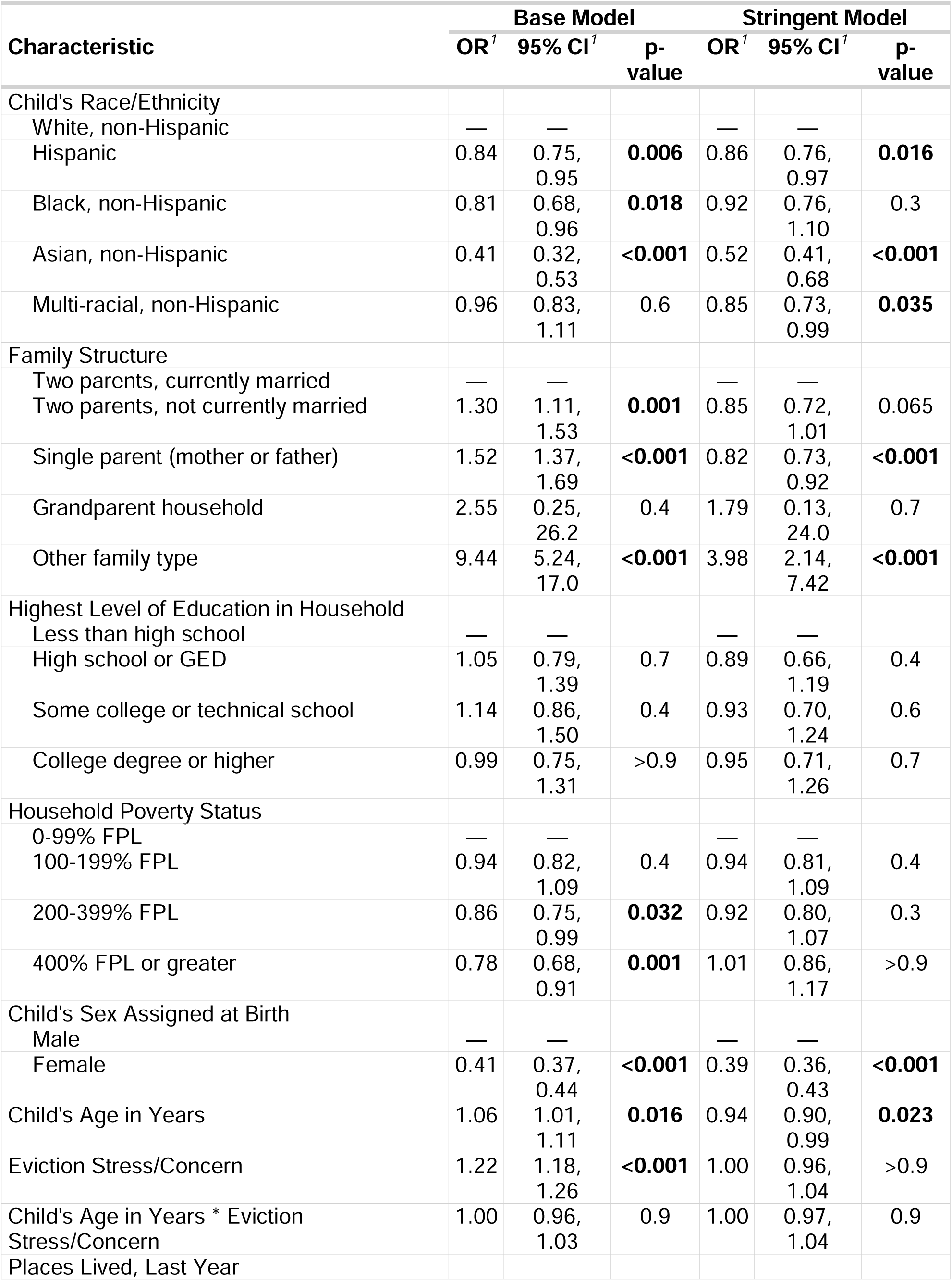
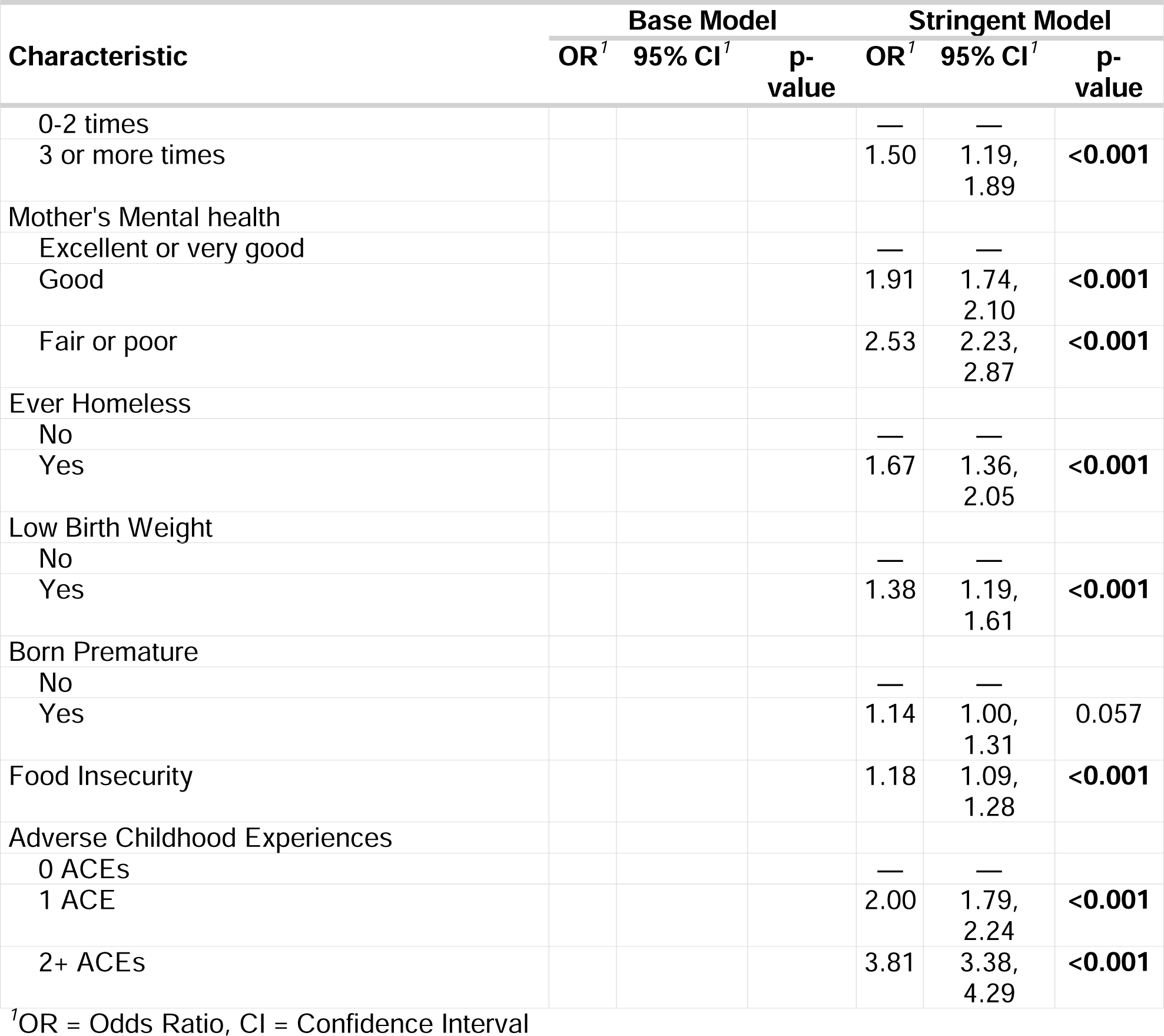
Odds ratios from generalized linear mixed models examining associations between caregiver eviction stress and child behavioral/conduct problems. The left side of the table shows adjustment using a base set of covariates, while the right side has a more stringent set of model adjustments/covariates.

Please see *Supplemental Materials* for statistical models: probing eviction X race interactions, re-running models after utilizing multiple data imputation techniques for missing data and investigating relations when removing younger (> 4 years of age) participants. This latter analysis was motivated by recent American Academy of Pediatrics guidelines noting that certain mental health diagnoses are uncommon or not appropriate for children younger than 4 years of age^42^. Our *Supplemental Materials* contain R-Markdown for all analysis in this main document.

## DISCUSSION

This study explored the association between caregivers’ stress about eviction or loss of housing and children’s mental health, namely depression, anxiety, ADHD, and behavioral/conduct problems. We probed whether these mental health outcomes were related to the main effect of stress about eviction or loss of housing, as well as the interaction of stress about eviction or loss of housing and age. We saw the strongest relations between stress about eviction or loss of housing and two forms of internalizing psychopathology, depression and anxiety. Stress about eviction or loss of housing was related to a 10-35% increase in incidence of these types of disorders. Relations between stress about eviction or loss of housing, ADHD, and behavioral/conduct problems were less robust, with models adjusting for a base set of covariates finding relations; however, these associations were not significant when adjusting estimates using a more stringent set of potential confounders. We saw some notable relations between mental health and the interaction of stress about eviction or loss of housing and age; specifically, relations between stress about eviction or loss of housing and depression and anxiety were strongest in younger samples, often becoming non-significant in older participants. Examined collectively, these findings provide valuable information to researchers interested in the role of eviction in child development, and in particular how this may influence mental health.

These results fit with past findings that eviction, foreclosures and housing loss may significantly impact development. Our findings connect to work noting increased depression in adolescents experiencing household and residential changes. We, however, did not see consistent relations between stress about eviction or loss of housing and externalizing problems. This diverges from past work noting that housing instability and experiencing more than three moves in childhood was associated with aggression and other disruptive behaviors^25^. This may be in part driven by our statistical modeling choices and the use of a stringent set of covariates. In both sets of models, we see that stress about eviction or loss of housing is related to depression; however, associations between stress about eviction or loss of housing and externalizing were only present in models adjusted for a base set of covariates and not in more stringently adjusted models. Externalizing problems were more associated with a child’s perinatal risk (i.e., low birth weight; premature birth), but not all projects have controlled for these variables. The impact of eviction, foreclosures and housing loss may be through more indirect developmental pathways, as it may contribute to perinatal risk, and this is then associated with externalizing challenges. As such, future research is needed to comprehensively understand relations between stress about eviction or loss of housing and externalizing problems.

Critical for future work, our results are unable to parse potential differential impacts of eviction versus housing foreclosure. Study participants were asked about multiple types of housing loss in one prompt (i.e., eviction, foreclosure, or having a house condemned). Few projects, to date, have thoroughly investigated whether one type of housing loss may exert greater impacts on child and family well-being. Some scholars have suggested that foreclosure may be stressful for a more prolonged duration compared to eviction^43^, while other researchers have lumped them under a similar category of “disruptive events”^44^. Work by Diamond and colleagues^45^ found renters who suffered eviction, due to landlords’ foreclosures, had fewer adverse effects, compared to those facing their own foreclosures; in this work, foreclosure on one’s own home was related to later housing instability, reduced homeownership, financial distress, moves to lower-quality neighborhoods, and elevated divorce rates. However, further comparative research, with direct comparison between eviction and foreclosure, is needed to fully understand the impacts of these different types of housing loss.

Reflecting on our work, this cohort was nationally representative and had a good deal of diversity in age, family structure, and socioeconomic status. Related to age, many past studies have only examined a very confined age range (e.g., middle childhood), but our work had participants from 3-17 years of age. With family structure, some past publications have used cohorts oversampling unmarried parents in large U.S. cities^46^. While appropriate, one must use statistical weights to have a truly nationally representative sample and make estimates generalizable to all families. Finally, connected to socioeconomic status, it is important to note that while eviction or housing loss is concentrated in families at or below the federal poverty line^7^, stress related to these experiences still occurs for those not considered “poor” (as detailed in our Supplement). Examined collectively, this dataset allows for a more inclusive assessment of how the stresses associated with eviction or loss of housing are linked to child well-being.

### Limitations

This study has several limitations. First, the work is cross-sectional in nature. Moving forward, longitudinal studies are needed to richly isolate the developmental impacts of eviction. This could be particularly informative related to the age X stress about eviction or loss of housing interactions that we described here. We find that at the highest levels of age in the cohort, stress about eviction or loss of housing was often no longer related to the presence of different disorders. This result, though very interesting, could be indexing conflicting, developmental phenomena. One possibility is that stress related to housing loss occurring early in life have outsized influences on development. Gaps in critical developmental skills may emerge early in childhood due to experiences before 5 years of age^47^. Alternatively, adolescents spend increasingly more time outside the home^48,49^ and the stress about loss of housing may have less “opportunity” to affect youth’s mental health. Prospectively following participants and seeing how families deal with stress about eviction or loss of housing will be important to fully understand potential interactions between housing loss and development. Second, and also connected to age, certain diagnoses are uncommon or not appropriate to diagnose in particularly young children^42^. While we find some significant age X stress about eviction or loss of housing interactions, we must be mindful that the incidence of many of these disorders is very low in the younger age range of our sample. We ran sensitivity analyses for child participants who were older than four years of age (see our *Supplemental Materials*) and find some similar age X stress about eviction or loss of housing interactions. Third, we were not able to incorporate rich geographical information into our statistical models. The NSCH only provided information about which states a family was residing in and we used this as a random factor in our statistical models. Other information could be incorporated (e.g., data from Princeton University’s Eviction Lab) to refine estimates and understand neighborhood-related impacts of eviction. Given variable state policies in eviction and housing protections, geographic variations could be important to understand. Finally, we used caregiver self-reports of mental health challenges; our work would have been strengthened by getting independent measures or clinical interviews of psychopathology. Use of multiple informants (e.g. teachers, clinicians) would have provided a comprehensive picture of the child’s functioning across different contexts (i.e., home; school^50^). This could be advantageous to help guide treatment and intervention strategies.

## Conclusions

Here, we find that stress about eviction or loss of housing is associated with an increased incidence of certain mental health problems in childhood. Given the profound long-term economic and social impacts of these problems^51^, it will be critical for communities to think about lessening housing precarity and better supporting families facing eviction, foreclosure or other housing loss. This could take many forms including rental assistance, legal aid for tenants, eviction diversion programs, and expanded social safety nets^52,53^. With millions facing evictions or foreclosures annually, those in public health and public policy must continue to push for expansion of these programs to reduce inequities and this key social determinant of child health.

## Supporting information

Supplemental Materials/Analyses

## Conflicts of Interest and Financial Disclosures

None to report

## Funding/Support and Role of Funder/Sponsor

The work was supported by internal funds provided to Dr. Hanson by the University of Pittsburgh; that university played no role in any element of the work

## Author Contributions

Dr Hanson had full access to all of the data in the study and takes responsibility for the integrity of the data and the accuracy of the data analysis.

## Data Sharing Statement

All data is publicly available from the Census website: https://www.census.gov/programs-surveys/nsch/data/datasets.html

## Disclaimer

The views expressed in this article are those of the authors and do not necessarily reflect the official policies of the US Department of Health and Human Services (HHS) or the Health Resources and Services Administration (HRSA), nor does mention of HHS or HRSA imply endorsement by the US government.

## Data Availability

Data is available from the Health Resources and Services Administration (HRSA) of the U.S. Department of Health and Human Services (HHS).

https://www.childhealthdata.org/learn-about-the-nsch/NSCH

## Notes

### Competing Interest Statement

The authors have declared no competing interest.

### Funding Statement

This work was supported by internal funding to Dr. Hanson.

### Author Declarations

This study utilized existing de-identified data from the publicly available 2022 National Survey of Children's Health. An ethics review and approval was not required as only de-identified secondary data were analyzed.

### Summary of Updates

-Updated main manuscript text and analyses -Significant expansion of supplemental materials

