## Supplemental Materials/Analyses for "Stress about Eviction or Loss of Housing and Child Mental Health"

Supplemental Materials for “*Stress about Eviction or Loss of Housing and Child Mental Health*

Jamie L Hanson, University of Pittsburgh

### Graphical Probing of Interaction Effects from Main Manuscript

To probe the nature of the significant interactions observed in the regression models (in the main manuscript), we visually examined the conditional effects using marginal effect plots. These graphical analyses allowed us to more deeply understand how the association between stress about eviction/housing loss and mental health issues varied across levels of age. Of note, we are only presenting these graphs when both base and stringently adjusted models found significant (p<.05) associations.

For depression and for stringently adjusted models, as noted in the main manuscript, the interaction of stress about eviction or loss of housing X age was significant (z=-2.505, p=0.012). For these models, the slope for stress about eviction or loss of housing was significant in younger participants, but not for older participants (-1SD age CI=0.05-0.29, z=2.76, p=0.006; at higher levels of age +1 SD CI= -0.03-0.06 z=0.76, p=0.445, as shown in Figure S01).

Figure S01.

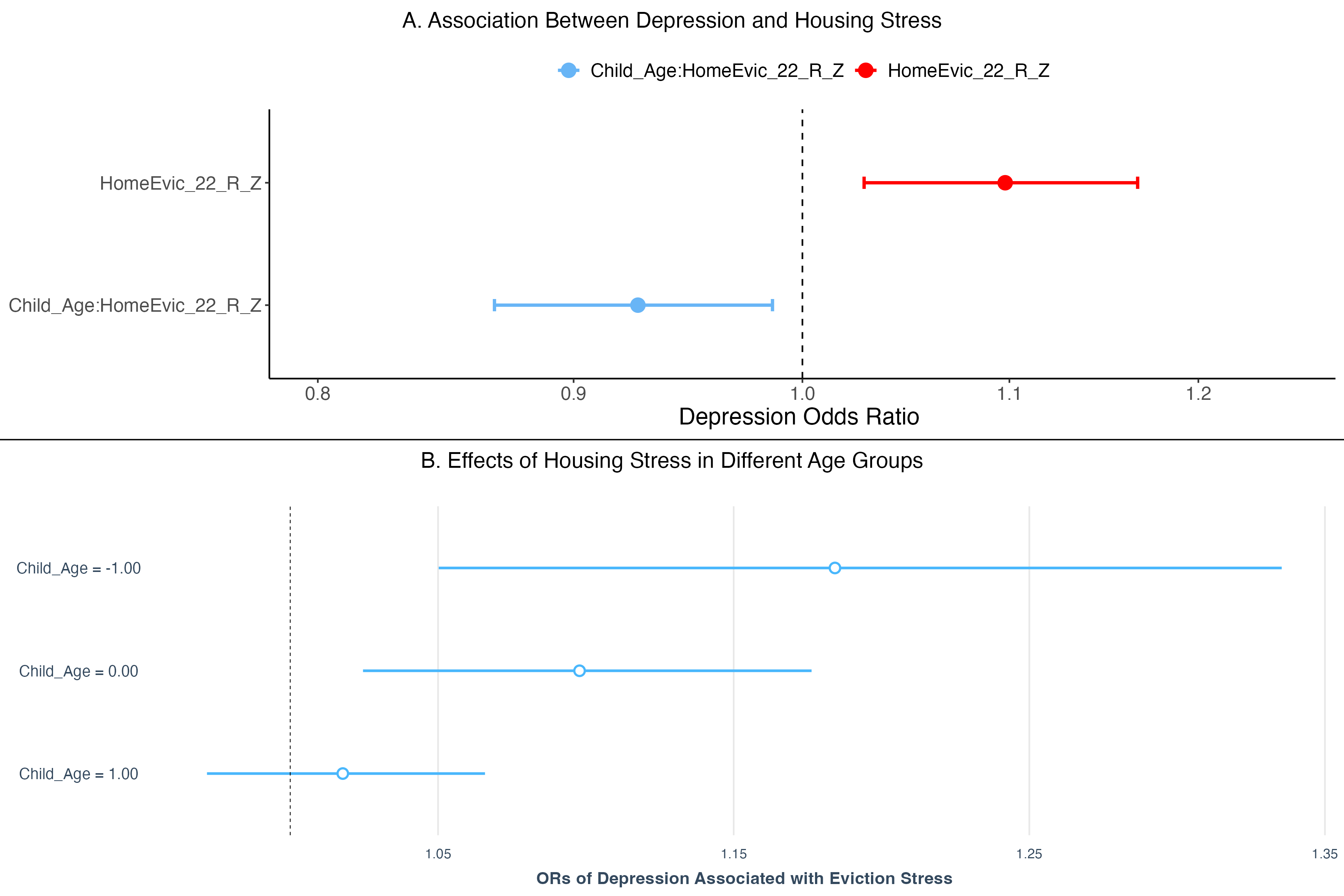

Caption: The top of this graph shows relations between eviction or housing loss related stress and the overall effects from our statistical models (mirroring results reported in the main manuscript); the bottom of the graph shows the odds ratios of depression associated with eviction or housing loss related stress for different child age groups (-1.00 Standard Deviation [SD], Mean, +1.00 SD). For both the younger (-1 SD) and average age participants, eviction stress is related to increases in depression; this is, however, not seen for older participants (+1.00 SD, near the bottom of that figure).

For the other mental health issues, as noted in the main manuscript, the interactions of stress about eviction or loss of housing X age were not significant. We therefore did not graphically unpack these associations.

### Relations Between Eviction Stress and Alternative Controls for Maternal Health

As a further test of robustness, we used a different index of maternal health, specifically maternal physical health. These models also included race/ethnicity, family structure, adult education, poverty level, sex, number of places lived, history of homelessness, low birth weight, prematurity, food insecurity, and adverse childhood experiences, age, stress about eviction or housing loss, and the interaction between age and stress about eviction or housing loss. State was included as a random factor in all models.

Paralleling results from the main manuscript, we found that stress about eviction or housing loss was related to depression (OR=1.12, CI=1.04-1.20, p=0.002) and anxiety (OR=1.06, CI=1.02-1.10, p=0.006). The interaction between age and stress about eviction or housing loss was related to depression (OR=0.93, CI=0.88, 0.99, p=0.025), but not anxiety (OR=0.97, CI=0.93, 1.00, p=0.083). Regarding ADHD and Behavioral Problems, stress about eviction or housing loss was not related to ADHD (OR=1.04, CI=1.00-1.08, p=0.075) or Behavioral Problems (OR=1.02, CI=0.98-1.06, p=0.4). The interaction of age and stress about eviction or housing loss was similarly not related to ADHD (OR=0.97, CI=0.94-1.01, p=0.2) or Behavioral Problems (OR=1.00, CI=0.97-1.04, p=0.8).

### Probing Associations Between Eviction Stress and Child Mental Health in Children Older than Four Years of Age

Given that some forms of poor mental health are uncommon or not appropriate to diagnose in particularly young children (e.g., ADHD, bipolar disorder)^1^, we re-ran all of the models from the main manuscript only including data from child participants who were older than four years of age. Of note, this meant our analytic sample dropped to N= 30432.

For depression, in both the base and stringently adjusted models, stress about eviction or housing loss showed significant associations (base: OR=1.35, CI=1.27-1.45, p<.001; stringent: OR=1.10, CI=1.03-1.19, p=.008), with a significant interaction with age in both cases (base: OR=0.93, CI=0.88-0.99, p=.017; stringent: OR=0.92, CI=0.87-0.98, p=.012), suggesting the impact of stress about eviction or housing loss varies by age. The outputs of these models, including all covariates, are depicted in Table S01. With anxiety, in base the both and stringently adjusted models, there were direct associations between stress about eviction or housing loss and anxiety disorders (base: OR=1.27, CI=1.22-1.32, p<.001; stringent: OR=1.05, CI=1.00-1.10, p=.031). There was however no significant eviction stress by age interactions in either adjusted model (base: OR=0.96, CI=0.92-1.0, p=.062; stringent: OR=0.96, CI=0.92-1.00, p=.077; as shown in Table S02).

| Table S01. | **Base Model** | | | **Stringent Model** | | |
| --- | --- | --- | --- | --- | --- | --- |
| **Characteristic** | **OR***^1^* | **95% CI***^1^* | **p-value** | **OR***^1^* | **95% CI***^1^* | **p-value** |
| Child's Race/Ethnicity |  |  |  |  |  |  |
| White, non-Hispanic | — | — |  | — | — |  |
| Hispanic | 0.76 | 0.66, 0.89 | **<0.001** | 0.75 | 0.64, 0.87 | **<0.001** |
| Black, non-Hispanic | 0.35 | 0.27, 0.46 | **<0.001** | 0.38 | 0.29, 0.50 | **<0.001** |
| Asian, non-Hispanic | 0.30 | 0.21, 0.42 | **<0.001** | 0.38 | 0.27, 0.54 | **<0.001** |
| Multi-racial, non-Hispanic | 1.00 | 0.84, 1.20 | >0.9 | 0.82 | 0.68, 0.98 | **0.034** |
| Family Structure |  |  |  |  |  |  |
| Two parents, currently married | — | — |  | — | — |  |
| Two parents, not currently married | 1.88 | 1.54, 2.29 | **<0.001** | 1.07 | 0.87, 1.32 | 0.5 |
| Single parent (mother or father) | 1.84 | 1.63, 2.08 | **<0.001** | 0.91 | 0.80, 1.04 | 0.2 |
| Grandparent household | 9.41 | 0.80, 111 | 0.075 | 5.79 | 0.47, 71.3 | 0.2 |
| Other family type | 5.57 | 2.39, 13.0 | **<0.001** | 2.27 | 0.92, 5.59 | 0.074 |
| Highest Level of Education in Household |  |  |  |  |  |  |
| Less than high school | — | — |  | — | — |  |
| High school or GED | 1.79 | 1.23, 2.60 | **0.002** | 1.55 | 1.05, 2.29 | **0.029** |
| Some college or technical school | 1.93 | 1.34, 2.79 | **<0.001** | 1.61 | 1.09, 2.37 | **0.016** |
| College degree or higher | 1.67 | 1.15, 2.42 | **0.007** | 1.66 | 1.13, 2.46 | **0.010** |
| Household Poverty Status |  |  |  |  |  |  |
| 0-99% FPL | — | — |  | — | — |  |
| 100-199% FPL | 0.95 | 0.80, 1.13 | 0.6 | 0.90 | 0.75, 1.08 | 0.3 |
| 200-399% FPL | 0.90 | 0.76, 1.06 | 0.2 | 0.97 | 0.81, 1.15 | 0.7 |
| 400% FPL or greater | 0.80 | 0.67, 0.96 | **0.014** | 1.03 | 0.86, 1.25 | 0.7 |
| Child's Sex Assigned at Birth |  |  |  |  |  |  |
| Male | — | — |  | — | — |  |
| Female | 1.88 | 1.70, 2.08 | **<0.001** | 1.91 | 1.72, 2.12 | **<0.001** |
| Child's Age in Years | 4.54 | 4.15, 4.98 | **<0.001** | 4.49 | 4.08, 4.95 | **<0.001** |
| Eviction Stress/Concern | 1.35 | 1.27, 1.45 | **<0.001** | 1.10 | 1.03, 1.19 | **0.008** |
| Child's Age in Years * Eviction Stress/Concern | 0.93 | 0.88, 0.99 | **0.017** | 0.92 | 0.87, 0.98 | **0.012** |
| Places Lived, Last Year |  |  |  |  |  |  |
| 0-2 times |  |  |  | — | — |  |
| 3 or more times |  |  |  | 1.86 | 1.41, 2.44 | **<0.001** |
| Ever Homeless |  |  |  |  |  |  |
| No |  |  |  | — | — |  |
| Yes |  |  |  | 1.42 | 1.12, 1.80 | **0.004** |
| Mother's Mental health |  |  |  |  |  |  |
| Excellent or very good |  |  |  | — | — |  |
| Good |  |  |  | 2.23 | 1.98, 2.50 | **<0.001** |
| Fair or poor |  |  |  | 3.28 | 2.80, 3.84 | **<0.001** |
| Low Birth Weight |  |  |  |  |  |  |
| No |  |  |  | — | — |  |
| Yes |  |  |  | 1.02 | 0.83, 1.24 | 0.9 |
| Born Premature |  |  |  |  |  |  |
| No |  |  |  | — | — |  |
| Yes |  |  |  | 1.10 | 0.92, 1.31 | 0.3 |
| Food Insecurity |  |  |  | 1.16 | 1.06, 1.28 | **0.002** |
| Adverse Childhood Experiences |  |  |  |  |  |  |
| 0 ACEs |  |  |  | — | — |  |
| 1 ACE |  |  |  | 1.83 | 1.58, 2.11 | **<0.001** |
| 2+ ACEs |  |  |  | 4.81 | 4.18, 5.54 | **<0.001** |
| *^1^*OR = Odds Ratio, CI = Confidence Interval | | | | | | |

*Caption: Odds ratios from generalized linear mixed models examining associations between caregiver eviction stress and child depression. Of note, this is for child participants > 4 years of age. The left side of the table shows adjustment using a base set of covariates, while the right side has a more stringent set of model adjustments.*

Table S02.

| Table S02. | **Base Model** | | | **Stringent Model** | | |
| --- | --- | --- | --- | --- | --- | --- |
| **Characteristic** | **OR***^1^* | **95% CI***^1^* | **p-value** | **OR***^1^* | **95% CI***^1^* | **p-value** |
| Child's Race/Ethnicity |  |  |  |  |  |  |
| White, non-Hispanic | — | — |  | — | — |  |
| Hispanic | 0.72 | 0.65, 0.80 | **<0.001** | 0.71 | 0.64, 0.79 | **<0.001** |
| Black, non-Hispanic | 0.32 | 0.27, 0.39 | **<0.001** | 0.34 | 0.28, 0.41 | **<0.001** |
| Asian, non-Hispanic | 0.25 | 0.20, 0.31 | **<0.001** | 0.30 | 0.24, 0.37 | **<0.001** |
| Multi-racial, non-Hispanic | 0.82 | 0.72, 0.93 | **0.002** | 0.71 | 0.62, 0.81 | **<0.001** |
| Family Structure |  |  |  |  |  |  |
| Two parents, currently married | — | — |  | — | — |  |
| Two parents, not currently married | 1.39 | 1.20, 1.60 | **<0.001** | 0.89 | 0.77, 1.04 | 0.15 |
| Single parent (mother or father) | 1.55 | 1.42, 1.69 | **<0.001** | 0.87 | 0.79, 0.95 | **0.004** |
| Grandparent household | 2.97 | 0.26, 33.9 | 0.4 | 2.01 | 0.17, 23.5 | 0.6 |
| Other family type | 3.12 | 1.54, 6.31 | **0.002** | 1.57 | 0.75, 3.30 | 0.2 |
| Highest Level of Education in Household |  |  |  |  |  |  |
| Less than high school | — | — |  | — | — |  |
| High school or GED | 1.61 | 1.21, 2.14 | **<0.001** | 1.40 | 1.04, 1.88 | **0.026** |
| Some college or technical school | 1.92 | 1.46, 2.54 | **<0.001** | 1.62 | 1.21, 2.17 | **0.001** |
| College degree or higher | 2.09 | 1.58, 2.76 | **<0.001** | 2.07 | 1.55, 2.77 | **<0.001** |
| Household Poverty Status |  |  |  |  |  |  |
| 0-99% FPL | — | — |  | — | — |  |
| 100-199% FPL | 1.08 | 0.95, 1.23 | 0.2 | 1.04 | 0.91, 1.19 | 0.6 |
| 200-399% FPL | 1.05 | 0.93, 1.19 | 0.4 | 1.11 | 0.98, 1.26 | 0.11 |
| 400% FPL or greater | 1.0 | 0.88, 1.13 | >0.9 | 1.21 | 1.06, 1.38 | **0.005** |
| Child's Sex Assigned at Birth |  |  |  |  |  |  |
| Male | — | — |  | — | — |  |
| Female | 1.55 | 1.45, 1.65 | **<0.001** | 1.57 | 1.47, 1.68 | **<0.001** |
| Child's Age in Years | 2.11 | 2.01, 2.21 | **<0.001** | 2.03 | 1.93, 2.13 | **<0.001** |
| Eviction Stress/Concern | 1.27 | 1.22, 1.32 | **<0.001** | 1.05 | 1.00, 1.10 | **0.031** |
| Child's Age in Years * Eviction Stress/Concern | 0.96 | 0.92, 1.00 | 0.062 | 0.96 | 0.92, 1.00 | 0.077 |
| Places Lived, Last Year |  |  |  |  |  |  |
| 0-2 times |  |  |  | — | — |  |
| 3 or more times |  |  |  | 1.30 | 1.05, 1.61 | **0.017** |
| Mother's Mental health |  |  |  |  |  |  |
| Excellent or very good |  |  |  | — | — |  |
| Good |  |  |  | 1.91 | 1.77, 2.07 | **<0.001** |
| Fair or poor |  |  |  | 2.75 | 2.45, 3.08 | **<0.001** |
| Ever Homeless |  |  |  |  |  |  |
| No |  |  |  | — | — |  |
| Yes |  |  |  | 1.25 | 1.03, 1.53 | **0.027** |
| Low Birth Weight |  |  |  |  |  |  |
| No |  |  |  | — | — |  |
| Yes |  |  |  | 1.00 | 0.88, 1.15 | >0.9 |
| Born Premature |  |  |  |  |  |  |
| No |  |  |  | — | — |  |
| Yes |  |  |  | 1.26 | 1.12, 1.42 | **<0.001** |
| Food Insecurity |  |  |  | 1.20 | 1.12, 1.29 | **<0.001** |
| Adverse Childhood Experiences |  |  |  |  |  |  |
| 0 ACEs |  |  |  | — | — |  |
| 1 ACE |  |  |  | 1.74 | 1.59, 1.90 | **<0.001** |
| 2+ ACEs |  |  |  | 3.33 | 3.02, 3.67 | **<0.001** |
| *^1^*OR = Odds Ratio, CI = Confidence Interval | | | | | | |

Caption: Odds ratios from generalized linear mixed models examining associations between caregiver eviction stress and child anxiety. *Of note, this is for child participants > 4 years of age. The left side of the table shows adjustment using a base set of covariates, while the right side has a more stringent set of model adjustments.*

For ADHD, stress about eviction or housing loss was significantly associated with ADHD in both the base and stringently adjusted models (base: OR=1.20, CI=1.16-1.25, p<.001; stringent: OR=1.05, CI=1.01-1.10, p=.025). There was also a significant interaction between stress about eviction or housing loss and child's age in the base model (OR=0.96, CI=0.92-1.00, p=.044), but not the stringently adjusted model (OR=0.96, CI=0.92-1.00, p=.055). These results are shown in Table S03. With behavioral problems, there was a main effect of stress about eviction or housing loss (OR=1.23, CI=1.18-1.28, p<.001), but this became non-significant in more stringently adjusted models (OR=1.02, CI=0.98-1.07, p=.4). In both the base and stringently adjusted models, interaction between stress about eviction or housing loss and child's age was not significant (base: OR=0.99, CI=0.95-104, p=.7; stringent: OR=0.99, CI=0.95-1.04, p=.8). These results are shown in Table S04.

| Table S03. | **Base Model** | | | **Stringent Model** | | |
| --- | --- | --- | --- | --- | --- | --- |
| **Characteristic** | **OR***^1^* | **95% CI***^1^* | **p-value** | **OR***^1^* | **95% CI***^1^* | **p-value** |
| Child's Race/Ethnicity |  |  |  |  |  |  |
| White, non-Hispanic | — | — |  | — | — |  |
| Hispanic | 0.72 | 0.64, 0.80 | **<0.001** | 0.72 | 0.64, 0.80 | **<0.001** |
| Black, non-Hispanic | 0.56 | 0.47, 0.66 | **<0.001** | 0.59 | 0.50, 0.70 | **<0.001** |
| Asian, non-Hispanic | 0.30 | 0.24, 0.38 | **<0.001** | 0.34 | 0.27, 0.44 | **<0.001** |
| Multi-racial, non-Hispanic | 0.92 | 0.81, 1.05 | 0.2 | 0.84 | 0.74, 0.96 | **0.010** |
| Family Structure |  |  |  |  |  |  |
| Two parents, currently married | — | — |  | — | — |  |
| Two parents, not currently married | 1.36 | 1.17, 1.57 | **<0.001** | 0.96 | 0.82, 1.12 | 0.6 |
| Single parent (mother or father) | 1.42 | 1.30, 1.56 | **<0.001** | 0.87 | 0.79, 0.97 | **0.008** |
| Grandparent household | 0.00 | 0.00, | >0.9 | 0.00 | 0.00, | >0.9 |
| Other family type | 4.66 | 2.37, 9.16 | **<0.001** | 2.53 | 1.26, 5.06 | **0.009** |
| Highest Level of Education in Household |  |  |  |  |  |  |
| Less than high school | — | — |  | — | — |  |
| High school or GED | 1.40 | 1.06, 1.84 | **0.017** | 1.23 | 0.93, 1.63 | 0.14 |
| Some college or technical school | 1.47 | 1.13, 1.93 | **0.005** | 1.26 | 0.96, 1.66 | 0.10 |
| College degree or higher | 1.31 | 1.00, 1.71 | 0.051 | 1.25 | 0.95, 1.64 | 0.12 |
| Household Poverty Status |  |  |  |  |  |  |
| 0-99% FPL | — | — |  | — | — |  |
| 100-199% FPL | 0.90 | 0.79, 1.02 | 0.10 | 0.88 | 0.77, 1.00 | **0.050** |
| 200-399% FPL | 0.93 | 0.82, 1.05 | 0.2 | 0.96 | 0.84, 1.08 | 0.5 |
| 400% FPL or greater | 0.90 | 0.79, 1.03 | 0.11 | 1.04 | 0.91, 1.19 | 0.5 |
| Child's Sex Assigned at Birth |  |  |  |  |  |  |
| Male | — | — |  | — | — |  |
| Female | 0.51 | 0.48, 0.55 | **<0.001** | 0.49 | 0.46, 0.53 | **<0.001** |
| Child's Age in Years | 1.48 | 1.41, 1.55 | **<0.001** | 1.38 | 1.32, 1.46 | **<0.001** |
| Eviction Stress/Concern | 1.20 | 1.16, 1.25 | **<0.001** | 1.05 | 1.01, 1.10 | **0.025** |
| Child's Age in Years * Eviction Stress/Concern | 0.96 | 0.92, 1.00 | **0.044** | 0.96 | 0.92, 1.00 | 0.055 |
| Places Lived, Last Year |  |  |  |  |  |  |
| 0-2 times |  |  |  | — | — |  |
| 3 or more times |  |  |  | 1.20 | 0.96, 1.50 | 0.11 |
| Mother's Mental health |  |  |  |  |  |  |
| Excellent or very good |  |  |  | — | — |  |
| Good |  |  |  | 1.56 | 1.44, 1.69 | **<0.001** |
| Fair or poor |  |  |  | 1.63 | 1.44, 1.84 | **<0.001** |
| Ever Homeless |  |  |  |  |  |  |
| No |  |  |  | — | — |  |
| Yes |  |  |  | 1.49 | 1.22, 1.81 | **<0.001** |
| Low Birth Weight |  |  |  |  |  |  |
| No |  |  |  | — | — |  |
| Yes |  |  |  | 1.28 | 1.12, 1.46 | **<0.001** |
| Born Premature |  |  |  |  |  |  |
| No |  |  |  | — | — |  |
| Yes |  |  |  | 1.17 | 1.04, 1.32 | **0.012** |
| Food Insecurity |  |  |  | 1.13 | 1.05, 1.21 | **0.001** |
| Adverse Childhood Experiences |  |  |  |  |  |  |
| 0 ACEs |  |  |  | — | — |  |
| 1 ACE |  |  |  | 1.77 | 1.62, 1.95 | **<0.001** |
| 2+ ACEs |  |  |  | 2.73 | 2.46, 3.02 | **<0.001** |
| *^1^*OR = Odds Ratio, CI = Confidence Interval | | | | | | |

*Caption: Odds ratios from generalized linear mixed models examining associations between caregiver eviction stress and child ADHD. Of note, this is for child participants > 4 years of age. The left side of the table shows adjustment using a base set of covariates, while the right side has a more stringent set of model adjustments.*

| Table S04. | **Base Model** | | | **Stringent Model** | | |
| --- | --- | --- | --- | --- | --- | --- |
| **Characteristic** | **OR***^1^* | **95% CI***^1^* | **p-value** | **OR***^1^* | **95% CI***^1^* | **p-value** |
| Child's Race/Ethnicity |  |  |  |  |  |  |
| White, non-Hispanic | — | — |  | — | — |  |
| Hispanic | 0.85 | 0.74, 0.96 | **0.010** | 0.86 | 0.75, 0.98 | **0.023** |
| Black, non-Hispanic | 0.81 | 0.67, 0.97 | **0.025** | 0.91 | 0.75, 1.09 | 0.3 |
| Asian, non-Hispanic | 0.39 | 0.30, 0.51 | **<0.001** | 0.50 | 0.38, 0.65 | **<0.001** |
| Multi-racial, non-Hispanic | 0.96 | 0.82, 1.12 | 0.6 | 0.86 | 0.73, 1.00 | 0.056 |
| Family Structure |  |  |  |  |  |  |
| Two parents, currently married | — | — |  | — | — |  |
| Two parents, not currently married | 1.38 | 1.17, 1.64 | **<0.001** | 0.89 | 0.74, 1.06 | 0.2 |
| Single parent (mother or father) | 1.51 | 1.35, 1.68 | **<0.001** | 0.80 | 0.71, 0.91 | **<0.001** |
| Grandparent household | 4.62 | 0.38, 56.6 | 0.2 | 2.35 | 0.13, 43.2 | 0.6 |
| Other family type | 11.1 | 5.77, 21.2 | **<0.001** | 5.28 | 2.65, 10.5 | **<0.001** |
| Highest Level of Education in Household |  |  |  |  |  |  |
| Less than high school | — | — |  | — | — |  |
| High school or GED | 1.03 | 0.77, 1.39 | 0.8 | 0.87 | 0.64, 1.18 | 0.4 |
| Some college or technical school | 1.13 | 0.84, 1.50 | 0.4 | 0.91 | 0.67, 1.22 | 0.5 |
| College degree or higher | 1.00 | 0.75, 1.34 | >0.9 | 0.94 | 0.70, 1.27 | 0.7 |
| Household Poverty Status |  |  |  |  |  |  |
| 0-99% FPL | — | — |  | — | — |  |
| 100-199% FPL | 0.95 | 0.82, 1.10 | 0.5 | 0.94 | 0.81, 1.10 | 0.4 |
| 200-399% FPL | 0.85 | 0.74, 0.98 | **0.029** | 0.90 | 0.78, 1.05 | 0.2 |
| 400% FPL or greater | 0.78 | 0.67, 0.91 | **0.002** | 0.98 | 0.84, 1.15 | 0.8 |
| Child's Sex Assigned at Birth |  |  |  |  |  |  |
| Male | — | — |  | — | — |  |
| Female | 0.42 | 0.38, 0.45 | **<0.001** | 0.39 | 0.36, 0.43 | **<0.001** |
| Child's Age in Years | 0.84 | 0.80, 0.89 | **<0.001** | 0.75 | 0.71, 0.80 | **<0.001** |
| Eviction Stress/Concern | 1.23 | 1.18, 1.28 | **<0.001** | 1.02 | 0.98, 1.07 | 0.4 |
| Child's Age in Years * Eviction Stress/Concern | 0.99 | 0.95, 1.04 | 0.7 | 0.99 | 0.95, 1.04 | 0.8 |
| Places Lived, Last Year |  |  |  |  |  |  |
| 0-2 times |  |  |  | — | — |  |
| 3 or more times |  |  |  | 1.57 | 1.24, 1.98 | **<0.001** |
| Mother's Mental health |  |  |  |  |  |  |
| Excellent or very good |  |  |  | — | — |  |
| Good |  |  |  | 1.95 | 1.77, 2.15 | **<0.001** |
| Fair or poor |  |  |  | 2.55 | 2.23, 2.91 | **<0.001** |
| Ever Homeless |  |  |  |  |  |  |
| No |  |  |  | — | — |  |
| Yes |  |  |  | 1.68 | 1.36, 2.08 | **<0.001** |
| Low Birth Weight |  |  |  |  |  |  |
| No |  |  |  | — | — |  |
| Yes |  |  |  | 1.38 | 1.18, 1.62 | **<0.001** |
| Born Premature |  |  |  |  |  |  |
| No |  |  |  | — | — |  |
| Yes |  |  |  | 1.18 | 1.02, 1.36 | **0.026** |
| Food Insecurity |  |  |  | 1.14 | 1.05, 1.24 | **0.002** |
| Adverse Childhood Experiences |  |  |  |  |  |  |
| 0 ACEs |  |  |  | — | — |  |
| 1 ACE |  |  |  | 1.93 | 1.71, 2.17 | **<0.001** |
| 2+ ACEs |  |  |  | 3.75 | 3.31, 4.24 | **<0.001** |
| *^1^*OR = Odds Ratio, CI = Confidence Interval | | | | | | |

*Caption: Odds ratios from generalized linear mixed models examining associations between caregiver eviction stress and child behavioral/conduct problems. Of note, this is for child participants > 4 years of age. The left side of the table shows adjustment using a base set of covariates, while the right side has a more stringent set of model adjustments.*

### Relations Between Eviction Stress and Demographic Characteristics

We were interested in understanding potential relations between stress about eviction or housing loss and two important demographic characteristics, namely socioeconomic status and race/ethnicity^2^. To examine relations in levels of reported eviction stress about eviction or housing loss across socioeconomic status, we constructed a one-way between-subjects Analysis of Variance (ANOVA) with eviction stress entered as the dependent variable, and a demographic characteristic (socioeconomic status category; race/ethnicity) entered as the independent variable. We first examined F-statistics and p-values from this ANOVA and then used post-hoc comparisons using Tukey's Honest Significant Difference (HSD) tests; this was to understand which groups different on stress about eviction or housing loss.

For this socioeconomic status variable, caregivers were asked about their income and then families were categorized into one of four groups: 0-99% Federal Poverty Line (FPL), 100-199% FPL, 200-399% FPL, and 400% FPL or greater. This was in relation to the U.S. Census Bureau definition of the FPL. Of note, the original survey responses from the National Survey of Children's Health had ~19% of this data missing, but it was imputed using multiple regression methods. The ANOVA comparing levels of reported eviction stress across these categories was highly statistically significant, F(*3, 52711*) = 1538, p < .001. This indicates significant differences exist in eviction stress among the four socioeconomic groups. Post-hoc comparisons using Tukey's HSD revealed households in the 0-99% FPL group reported significantly higher eviction stress than all other groups: the 100-199% FPL group, the 200-399% FPL group, and the 400% FPL or greater group (p<.001 for all of these post-hoc pairwise comparisons). Additionally, the 200-399% FPL and 400% FPL or greater groups both reported significantly lower eviction stress than the 100-199% FPL group. Finally, eviction stress was also significantly lower in the 400% FPL or greater group compared to the 200-399% FPL group. In summary, lower socioeconomic status households (0-99% FPL) face the greatest levels of eviction stress/concern, with this stress decreasing in higher socioeconomic status groups. These differences are shown in Figure S02.

Figure S02.

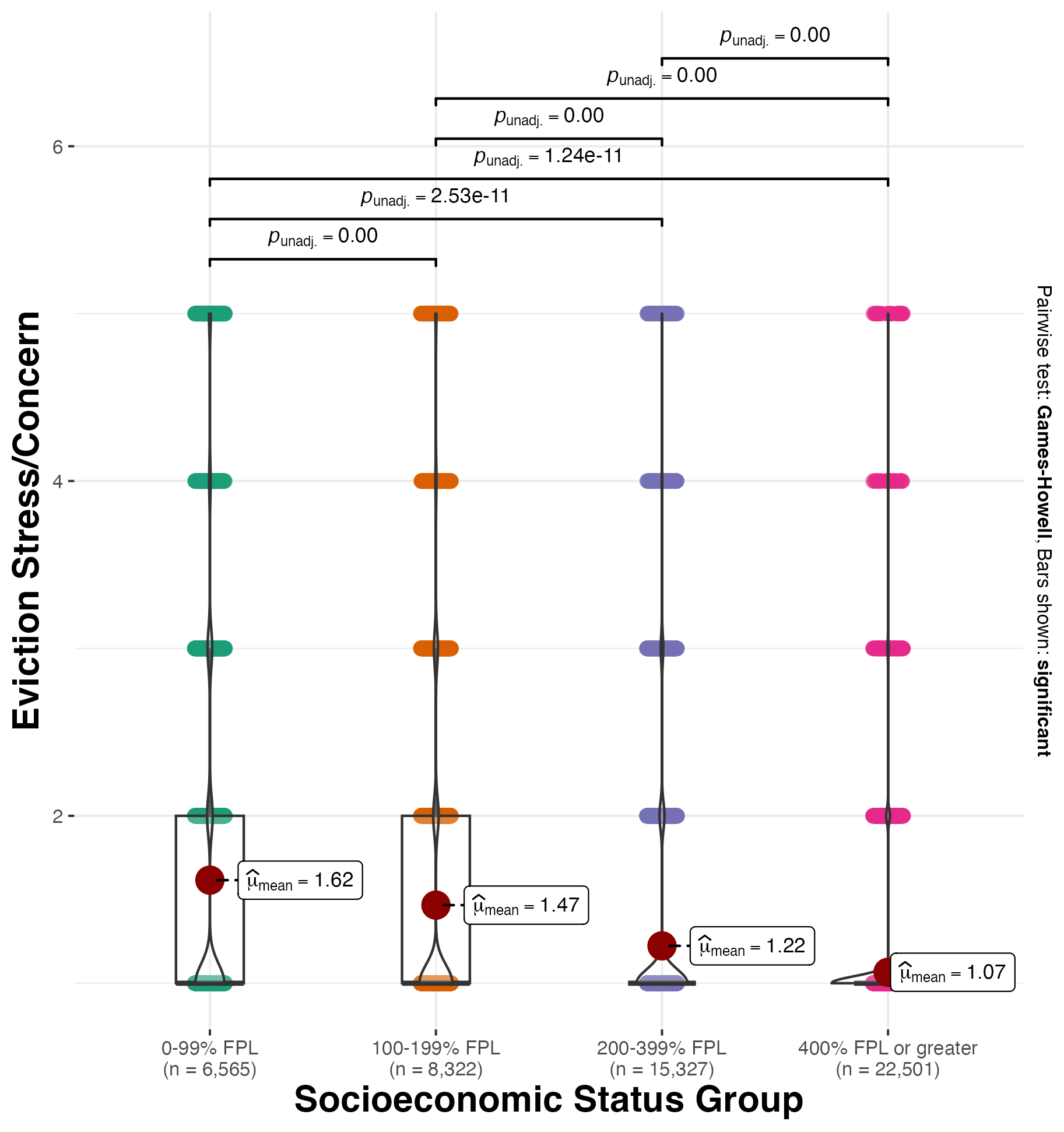

*Caption: Mean levels of reported caregiver eviction stress across categories of family socioeconomic status.*

Turning to potential differences in stress about eviction or housing loss across categories of racial/ethnic background, we compared these different racial/ethnic groups: White (non-Hispanic), Black (non-Hispanic), Asian (non-Hispanic), Multi-racial (non-Hispanic), and Hispanic. These were based on questions to caregivers about their child’s ethnicity (Hispanic, Latino, or Spanish origin; or not) and an open-ended question about the child's race. Children whose parents reported Hispanic/Latino ethnicity were characterized as Hispanic, regardless of their reported race. Non-Hispanic children were classified by their response to the race question, with some race categories combined into an "Other non-Hispanic" group due to small sample sizes. The ANOVA comparing levels of reported eviction stress across categories of the race/ethnicity variable was highly statistically significant, F(*4, 52710*) = 363.2, p < .001. The post-hoc Tukey HSD test found significantly higher eviction stress reported among Black, non-Hispanic individuals compared to all other groups (p<.001 for all of these post-hoc pairwise comparisons). Hispanic individuals also reported significantly higher stress than White, non-Hispanic individuals. Asian, non-Hispanic individuals reported significantly lower stress than Black or Hispanic groups. Multi-racial, non-Hispanic individuals were significantly higher than Asian individuals but lower than Black individuals. These differences are shown in Figure S03.

Figure S03.

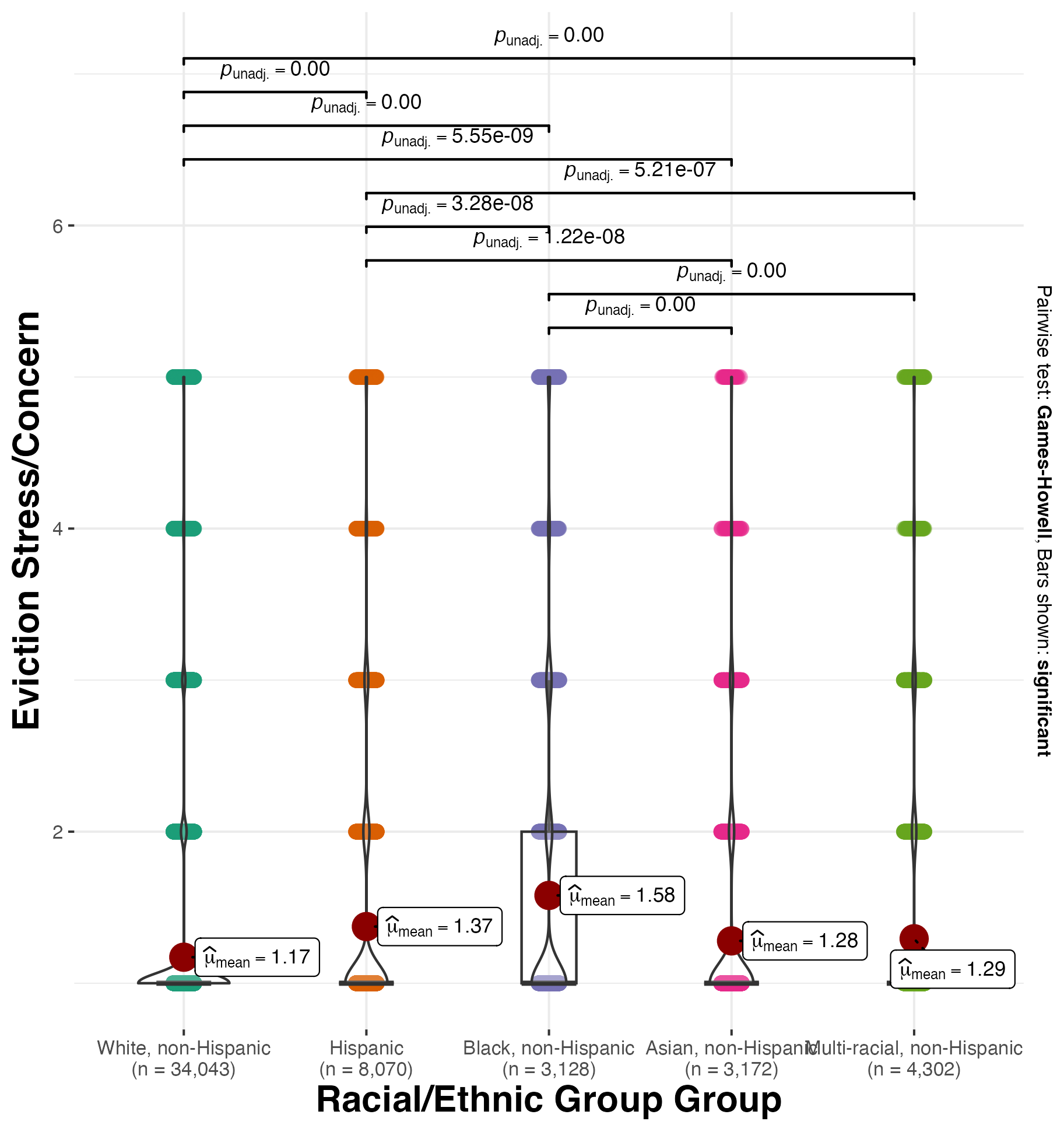

*Caption: Mean levels of reported caregiver eviction stress across categories of child race/ethnicity.*

### Probing Eviction Stress X Race Interactions

Given that race is strongly associated with eviction and Black communities face disproportionately higher rates of eviction compared to other groups^24^, we were interested in examining the interaction of stress about eviction or housing loss and race/ethnicity in predicting different forms of psychopathology. To investigate this idea, we constructed statistical models similar to those in the main manuscript (with model adjustment using a base and a stringent set of covariates) for our four indicators of mental health issues– depression, anxiety, ADHD, and behavioral/conduct problems. Like procedures in the main manuscript, we sought to plot the data and test differences for the simple slopes of the association between stress about eviction or housing loss for different racial/ethnic groups in relation to any reported problem. We used a base model and a more stringent set of covariates for model adjustment, and examined binary indicators of depression, anxiety, ADHD, and behavioral/conduct problems separately with generalized linear mixed models with a logistic link function and state as a random factor. Of note, below, we report the main effects of eviction and race, and the interaction between eviction X races. With the interactions, white participants were used as the reference group.

For depression, in base adjusted models, stress about eviction or housing loss (as a main effect) was significantly related to higher incidences of this mental health issue (base model odds ratio [OR]=1.32, CI=1.25-1.39, z=10.827, p<.001). In the base model, the interaction of stress about eviction or housing loss X race was significant for Hispanic participants (z=-2.563, p=0.01). Unpacking this interaction, eviction stress had a lower slope for Hispanic participants. In relation to depression, the slope of eviction stress for white participants was β=0.28, CI=0.23-0.33, while the slope of eviction stress for Hispanic participants was β=0.15, CI=0.06-0.23. This interaction was not significant for any other racial/ethnic groups (all p’s>0.08). In stringently adjusted models, all interactions between eviction stress X race were non-significant (all p’s>0.09). Effects are noted in Table S05 and graphically depicted in Figure S05.

| Table S05. | **Base Model** | | | **Stringent Model** | | |
| --- | --- | --- | --- | --- | --- | --- |
| **Characteristic** | **OR***^1^* | **95% CI***^1^* | **p-value** | **OR***^1^* | **95% CI***^1^* | **p-value** |
| Eviction Stress/Concern | 1.32 | 1.25, 1.39 | **<0.001** | 1.04 | 0.98, 1.10 | 0.2 |
| Child's Race/Ethnicity |  |  |  |  |  |  |
| White, non-Hispanic | — | — |  | — | — |  |
| Hispanic | 0.81 | 0.69, 0.94 | **0.006** | 0.78 | 0.66, 0.91 | **0.002** |
| Black, non-Hispanic | 0.34 | 0.25, 0.46 | **<0.001** | 0.35 | 0.25, 0.47 | **<0.001** |
| Asian, non-Hispanic | 0.31 | 0.22, 0.44 | **<0.001** | 0.39 | 0.27, 0.55 | **<0.001** |
| Multi-racial, non-Hispanic | 1.01 | 0.84, 1.22 | 0.9 | 0.81 | 0.67, 0.99 | **0.036** |
| Child's Race/Ethnicity * Eviction Stress/Concern |  |  |  |  |  |  |
| Hispanic * Eviction Stress/Concern | 0.88 | 0.79, 0.97 | **0.010** | 0.93 | 0.84, 1.03 | 0.2 |
| Black, non-Hispanic * Eviction Stress/Concern | 0.99 | 0.87, 1.13 | 0.9 | 1.07 | 0.93, 1.23 | 0.3 |
| Asian, non-Hispanic * Eviction Stress/Concern | 0.75 | 0.54, 1.04 | 0.086 | 0.87 | 0.62, 1.22 | 0.4 |
| Multi-racial, non-Hispanic * Eviction Stress/Concern | 0.93 | 0.82, 1.05 | 0.3 | 0.99 | 0.87, 1.12 | 0.8 |
| *^1^*OR = Odds Ratio, CI = Confidence Interval | | | | | | |

*Caption: Adjusted odds ratios from models examining associations between caregiver eviction stress and child depression, testing the interaction between eviction stress and race/ethnicity. The left side of the table shows adjustment using a base set of covariates, while the right side has a more stringent set of model adjustments.*

Figure S05.

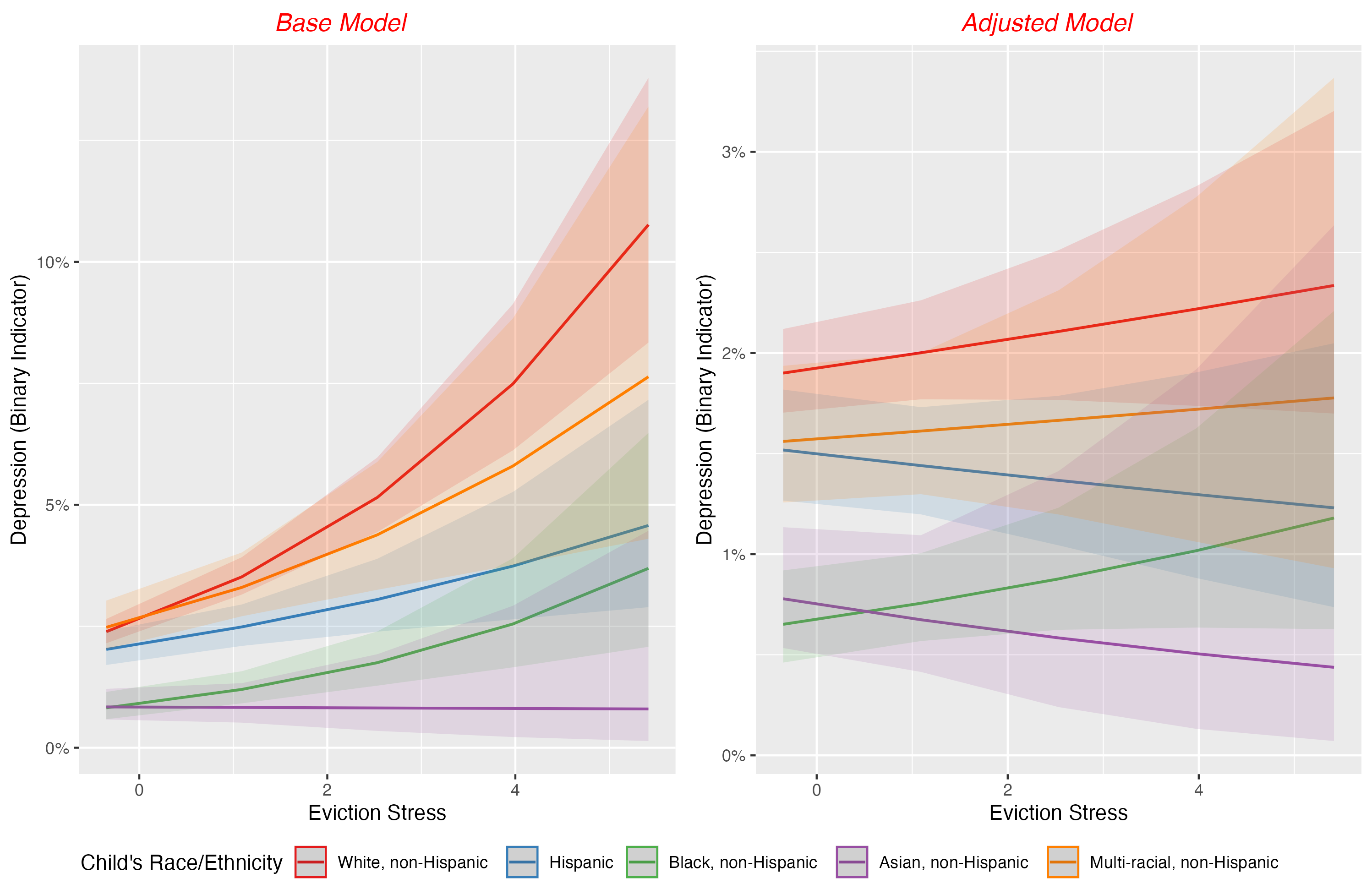

*Caption: Predicted probabilities of a child having depression from models testing the interaction between eviction stress and race/ethnicity.*

With anxiety, in base adjusted models, stress about eviction or housing loss (as a main effect) was significantly related to higher incidences of this mental health issue (base model OR=1.29, CI=1.24-1.34, z=13.094, p<.001). In the base model, the interaction of eviction stress X race was significant for Hispanic participants (z=-3.091, p=0.002) and Black participants (z=-1.984, p=0.047). Unpacking this interaction, stress about eviction or housing loss had a lower slope for Hispanic participants. In relation to anxiety, the slope of eviction stress for white participants was β=0.25, CI=0.21-0.29, while the slope of eviction stress for Hispanic participants was β=0.14, CI=0.07-0.20 and Black participants was β=0.15, CI=0.05-0.24. This interaction was not significant for any other racial/ethnic groups (all p’s>0.056). In stringently adjusted models, all interactions between eviction stress X race were non-significant (all p’s>0.11). Effects are noted in Table S06 and graphically depicted in Figure S06.

| Table S06. | **Base Model** | | | **Stringent Model** | | |
| --- | --- | --- | --- | --- | --- | --- |
| **Characteristic** | **OR***^1^* | **95% CI***^1^* | **p-value** | **OR***^1^* | **95% CI***^1^* | **p-value** |
| Eviction Stress/Concern | 1.29 | 1.24, 1.34 | **<0.001** | 1.03 | 0.99, 1.08 | 0.12 |
| Child's Race/Ethnicity |  |  |  |  |  |  |
| White, non-Hispanic | — | — |  | — | — |  |
| Hispanic | 0.75 | 0.67, 0.83 | **<0.001** | 0.73 | 0.66, 0.81 | **<0.001** |
| Black, non-Hispanic | 0.35 | 0.28, 0.42 | **<0.001** | 0.35 | 0.28, 0.43 | **<0.001** |
| Asian, non-Hispanic | 0.26 | 0.21, 0.32 | **<0.001** | 0.30 | 0.24, 0.38 | **<0.001** |
| Multi-racial, non-Hispanic | 0.84 | 0.74, 0.95 | **0.007** | 0.73 | 0.64, 0.83 | **<0.001** |
| Child's Race/Ethnicity * Eviction Stress/Concern |  |  |  |  |  |  |
| Hispanic * Eviction Stress/Concern | 0.89 | 0.83, 0.96 | **0.002** | 0.94 | 0.87, 1.01 | 0.11 |
| Black, non-Hispanic * Eviction Stress/Concern | 0.90 | 0.81, 1.00 | **0.047** | 0.98 | 0.88, 1.09 | 0.7 |
| Asian, non-Hispanic * Eviction Stress/Concern | 0.92 | 0.77, 1.10 | 0.4 | 1.05 | 0.87, 1.27 | 0.6 |
| Multi-racial, non-Hispanic * Eviction Stress/Concern | 0.94 | 0.85, 1.04 | 0.2 | 0.98 | 0.89, 1.08 | 0.7 |
| *^1^*OR = Odds Ratio, CI = Confidence Interval | | | | | | |

*Caption: Adjusted odds ratios from models examining associations between caregiver eviction stress and child anxiety, testing the interaction between eviction stress and race/ethnicity. The left side of the table shows adjustment using a base set of covariates, while the right side has a more stringent set of model adjustments.*

Figure S06.

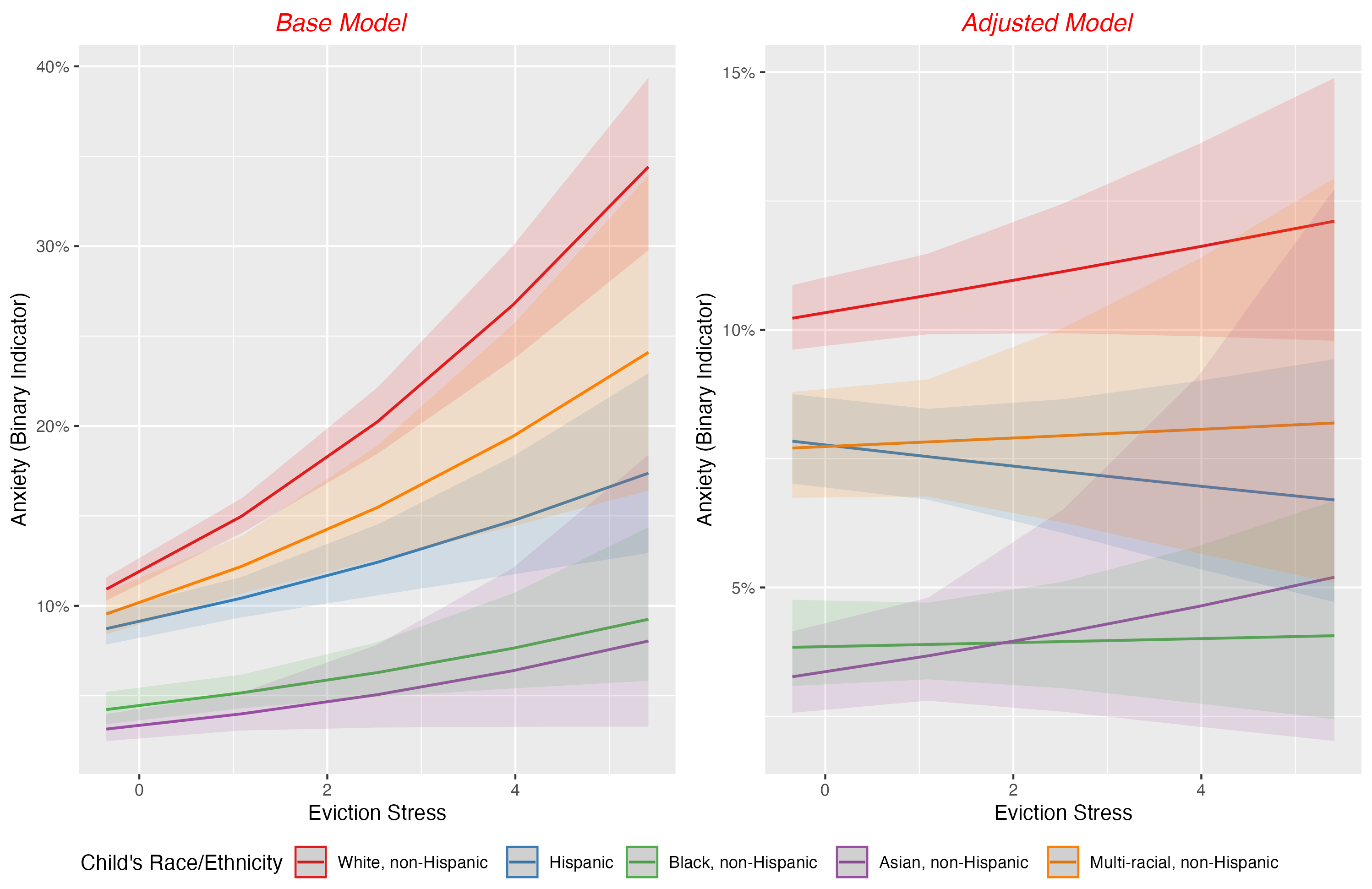

*Caption: Predicted probabilities of a child having anxiety from models testing the interaction between eviction stress and race/ethnicity.*

Related to ADHD, in base and stringently adjusted models, stress about eviction or housing loss (as a main effect) was significantly related to higher incidences of this mental health issue (base model odds ratio [OR]=1.25, CI=1.20-1.30, z=11.281, p<.001; stringent model OR=1.07, CI=1.02-1.1, z=2.927, p=0.03). In the base model, the interaction of stress about eviction or housing loss X race was significant for Hispanic (z=-4.546, p<0.001), Black (z=-2.878, p=0.003), and multi-racial (z=-2.541, p=0.01) participants. Unpacking this interaction, eviction stress had a lower slope for these participants. In relation to ADHD, the slope of eviction stress for white participants was β=0.22, CI=0.18-0.26, while the slope of eviction stress for Hispanic participants was β=0.03, CI=-0.04-0.11, Black participants was β=0.08, CI=-0.01-0.17, and multi-racial participants was β=0.09, CI=0.00-0.19. In stringently adjusted models, the interaction of eviction stress X race was significant for Hispanic (z=3.766, p<0.001) and multi-racial (z=-1.962, p=0.049) participants. In this model, the slope of eviction stress for white participants was β=0.06, CI=0.02-0.11, while the slope of eviction stress for Hispanic participants was β=-0.10, CI=-0.18- -0.02 and for multi-racial participants was β=-0.04, CI=-0.14-0.06. This interaction was not significant for any other racial/ethnic groups (all p’s>0.09). Effects are noted in Table S07 and graphically depicted in Figure S07.

| Table S07. | **Base Model** | | | **Stringent Model** | | |
| --- | --- | --- | --- | --- | --- | --- |
| **Characteristic** | **OR***^1^* | **95% CI***^1^* | **p-value** | **OR***^1^* | **95% CI***^1^* | **p-value** |
| Eviction Stress/Concern | 1.25 | 1.20, 1.30 | **<0.001** | 1.07 | 1.02, 1.11 | **0.003** |
| Child's Race/Ethnicity |  |  |  |  |  |  |
| White, non-Hispanic | — | — |  | — | — |  |
| Hispanic | 0.75 | 0.67, 0.84 | **<0.001** | 0.75 | 0.67, 0.84 | **<0.001** |
| Black, non-Hispanic | 0.60 | 0.50, 0.71 | **<0.001** | 0.61 | 0.51, 0.73 | **<0.001** |
| Asian, non-Hispanic | 0.31 | 0.25, 0.39 | **<0.001** | 0.36 | 0.28, 0.45 | **<0.001** |
| Multi-racial, non-Hispanic | 0.93 | 0.82, 1.06 | 0.3 | 0.85 | 0.74, 0.96 | **0.012** |
| Child's Race/Ethnicity * Eviction Stress/Concern |  |  |  |  |  |  |
| Hispanic * Eviction Stress/Concern | 0.83 | 0.76, 0.90 | **<0.001** | 0.85 | 0.78, 0.92 | **<0.001** |
| Black, non-Hispanic * Eviction Stress/Concern | 0.87 | 0.78, 0.95 | **0.004** | 0.92 | 0.83, 1.02 | 0.10 |
| Asian, non-Hispanic * Eviction Stress/Concern | 0.92 | 0.76, 1.11 | 0.4 | 0.99 | 0.82, 1.20 | >0.9 |
| Multi-racial, non-Hispanic * Eviction Stress/Concern | 0.88 | 0.79, 0.97 | **0.011** | 0.90 | 0.81, 1.00 | **0.050** |
| *^1^*OR = Odds Ratio, CI = Confidence Interval | | | | | | |

*Caption: Adjusted odds ratios from models examining associations between caregiver eviction stress and child ADHD, testing the interaction between eviction stress and race/ethnicity. The left side of the table shows adjustment using a base set of covariates, while the right side has a more stringent set of model adjustments.*

Figure S07.

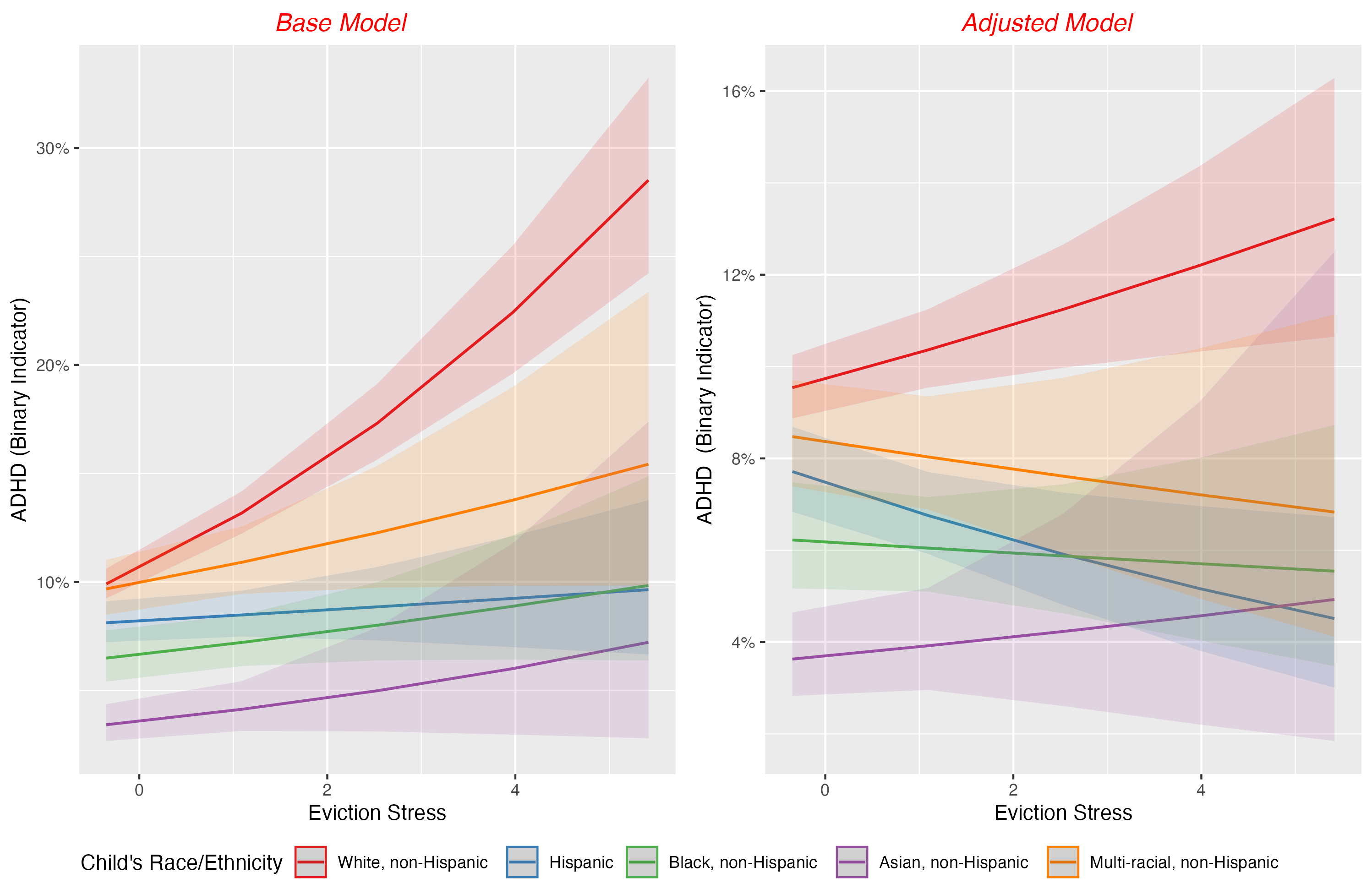

*Caption: Predicted probabilities of a child having ADHD from models testing the interaction between eviction stress and race/ethnicity.*

Finally, regarding behavioral/conduct problems, in base adjusted models, eviction stress (as a main effect) was significantly related to higher incidences of this mental health issue (base model odds ratio [OR]=1.30, CI=1.24-1.35, z=12.19, p<.001). In the base model, the interaction of eviction stress X race was significant for Hispanic (z=-3.602, p<0.001) and Black (z=3.080, p=0.002) participants. Unpacking this interaction, eviction stress had a lower slope for these participants. In relation to ADHD, the slope of eviction stress for white participants was β=0.26, CI=0.22-0.3, while the slope of eviction stress for Hispanic participants was β=0.11, CI=-0.03-0.18 and for Black participants was β=0.10, CI=-0.01-0.19. In stringently adjusted models, the interaction of eviction stress X race was significant for Hispanic participants (z=-2.509, p=0.012). In this model, the slope of eviction stress for white participants was β=0.04, CI=0.00-0.09, while the slope of eviction stress for Hispanic participants was β=-0.07, CI=-0.15- -0.01. This interaction was not significant for any other racial/ethnic groups (all p’s>0.11). Effects are noted in Table S08 and graphically depicted in Figure S08.

| Table S08. | **Base Model** | | | **Stringent Model** | | |
| --- | --- | --- | --- | --- | --- | --- |
| **Characteristic** | **OR***^1^* | **95% CI***^1^* | **p-value** | **OR***^1^* | **95% CI***^1^* | **p-value** |
| Eviction Stress/Concern | 1.30 | 1.24, 1.35 | **<0.001** | 1.04 | 1.00, 1.09 | 0.074 |
| Child's Race/Ethnicity |  |  |  |  |  |  |
| White, non-Hispanic | — | — |  | — | — |  |
| Hispanic | 0.89 | 0.78, 1.00 | 0.057 | 0.89 | 0.79, 1.01 | 0.075 |
| Black, non-Hispanic | 0.89 | 0.73, 1.07 | 0.2 | 0.96 | 0.79, 1.16 | 0.6 |
| Asian, non-Hispanic | 0.42 | 0.33, 0.55 | **<0.001** | 0.53 | 0.41, 0.68 | **<0.001** |
| Multi-racial, non-Hispanic | 0.97 | 0.84, 1.13 | 0.7 | 0.86 | 0.74, 1.00 | 0.056 |
| Child's Race/Ethnicity * Eviction Stress/Concern |  |  |  |  |  |  |
| Hispanic * Eviction Stress/Concern | 0.86 | 0.79, 0.93 | **<0.001** | 0.89 | 0.82, 0.98 | **0.012** |
| Black, non-Hispanic * Eviction Stress/Concern | 0.85 | 0.77, 0.94 | **0.002** | 0.92 | 0.82, 1.02 | 0.11 |
| Asian, non-Hispanic * Eviction Stress/Concern | 0.87 | 0.71, 1.07 | 0.2 | 0.96 | 0.78, 1.19 | 0.7 |
| Multi-racial, non-Hispanic * Eviction Stress/Concern | 0.91 | 0.82, 1.01 | 0.090 | 0.94 | 0.84, 1.05 | 0.3 |
| *^1^*OR = Odds Ratio, CI = Confidence Interval | | | | | | |

*Caption: Adjusted odds ratios from models examining associations between caregiver eviction stress and child behavioral/conduct problems, testing the interaction between eviction stress and race/ethnicity. The left side of the table shows adjustment using a base set of covariates, while the right side has a more stringent set of model adjustments.*

Figure S08.

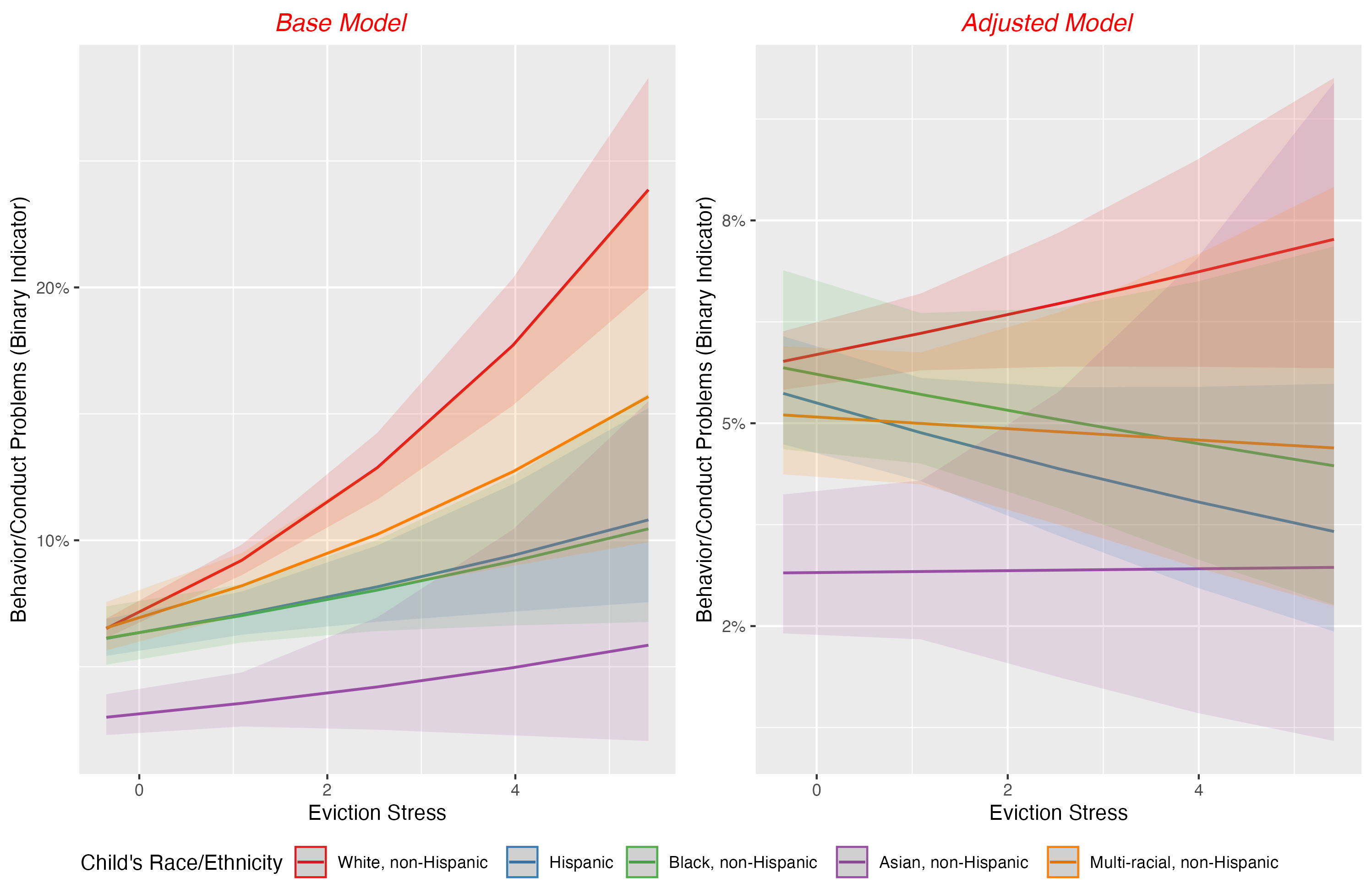

Caption: Predicted probabilities of a child having behavioral/conduct problems from models testing the interaction between eviction stress and race/ethnicity.

### Using Data Imputation for Missing Independent Variables

Across our models, we had a large degree of missingness (~32%). Over 8,600 participants did not have our dependent variables of interest. An additional, approximately 8,000 participants were missing independent variables (i.e., our large set of sociodemographic covariates). To address missing data, we employed three different imputation methods for comparison: predictive mean matching (PMM), k-nearest neighbors (KNN), and random forest.

First, using the *mice* package^3^ in R, we performed PMM with 50 imputations and 25 iterations. PMM maintains the observed distribution of values by imputing real values from similar cases. Second, we applied KNN imputation (using the KNNimp function from the *multiUS* R library^4^), which fills missing values using data from the k-most similar complete cases. Third, we implemented random forest imputation using the *missForest* package^5^. This approach uses an ensemble of decision trees to predict missing values based on observed data patterns and was set to a maximum of 10 iterations.

For all three approaches, we imputed values for twenty (independent) variables including demographic characteristics (race, family structure, adult education, poverty level), risk factors (places lived, maternal physical and mental health, adverse childhood experiences), and child characteristics (sex, homelessness history, birth weight, prematurity, age, stress about eviction or housing loss, state of residence, food security). Our dependent variables, mental health outcomes: depression, anxiety, ADHD, and behavioral problems, were included during the data imputation process but then replaced (with *‘NA’*); this was done to preserve their authentic distributions of dependent variables. Our final sample with partially imputed data was N=45214.

Across all three imputation approaches, depressive disorders showed consistent associations with stress about eviction or housing loss and its interaction with age (as shown in Table S09). Using predictive mean matching (PMM), stress about eviction or housing loss was associated with increased odds of depression (OR=1.13, CI=1.07-1.20, p<.001), with a significant age interaction (OR=0.92, CI=0.88-0.97, p<.001). Similar patterns emerged using K-nearest neighbor imputation (eviction: OR=1.10, CI=1.03-1.16, p=.002; age interaction: OR=0.93, CI=0.88-0.97, p=.003) and Random Forest imputation (eviction: OR=1.09, CI=1.03-1.16, p=.004; age interaction: OR=0.93, CI=0.88-0.97, p=.003).

Table S09.

| Associations with Depressive Disorders | | | | |
| --- | --- | --- | --- | --- |
| Characteristics | Odds Ratio | 95% CI | p-value | Imputation |
| Eviction Stress/Concern | 1.1292633 | 1.07-1.20 | p<.001 | PMM |
| Age * Eviction Stress/Concern | 0.9204410 | 0.88-0.97 | p<.001 | PMM |
| Eviction Stress/Concern | 1.0964009 | 1.03-1.16 | 0.002 | K-nearest neighbor |
| Age * Eviction Stress/Concern | 0.9259070 | 0.88-0.97 | 0.003 | K-nearest neighbor |
| Eviction Stress/Concern | 1.0893620 | 1.03-1.16 | 0.004 | Random Forest |
| Age * Eviction Stress/Concern | 0.9259516 | 0.88-0.97 | 0.003 | Random Forest |

For anxiety disorders, PMM imputation revealed significant associations with stress about eviction or housing loss (OR=1.06, CI=1.02-1.10, p=.002) and its interaction with age (OR=0.96, CI=0.93-1.00, p=.028). K-nearest neighbor imputation showed similar patterns, though with slightly smaller effect sizes for eviction stress (OR=1.04, CI=1.00-1.08, p=.037) and age interaction (OR=0.95, CI=0.92-0.99, p=.005). Using Random Forest imputation, the association with stress about eviction or housing loss was not significant (OR=1.03, CI=1.00-1.07, p=.087), while the age interaction remained significant (OR=0.96, CI=0.93-0.99, p=.006). These results are shown In Table S10.

Table S10.

| Associations with Anxiety Disorders | | | | | | |
| --- | --- | --- | --- | --- | --- | --- |
| Characteristics | Odds Ratio | | 95% CI | p-value | | Imputation |
| Eviction Stress/Concern | 1.0582015 | 1.02-1.10 | | 0.002 | PMM | |
| Age * Eviction Stress/Concern | 0.9640277 | 0.93-1.00 | | 0.028 | PMM | |
| Eviction Stress/Concern | 1.0383911 | 1.00-1.08 | | 0.037 | K-nearest neighbor | |
| Age * Eviction Stress/Concern | 0.9542941 | 0.92-0.99 | | 0.005 | K-nearest neighbor | |
| Eviction Stress/Concern | 1.0315071 | 1.00-1.07 | | 0.087 | Random Forest | |
| Age * Eviction Stress/Concern | 0.9559828 | 0.93-0.99 | | 0.006 | Random Forest | |

With ADHD, PMM imputation showed modest but significant associations with both stress about eviction or housing loss (OR=1.03, CI=1.00-1.07, p=.041) and the age interaction (OR=0.97, CI=0.94-1.00, p=.042). Results from K-nearest neighbor imputation revealed a marginally significant association with eviction stress (OR=1.03, CI=1.00-1.07, p=.067) and a significant age interaction (OR=0.96, CI=0.93-1.00, p=.025). Similarly, Random Forest imputation showed a marginally significant association with eviction stress (OR=1.03, CI=1.00-1.07, p=.080) and a significant age interaction (OR=0.96, CI=0.93-1.00, p=.026). These results are show In Table S11.

Table S11.

| Associations with ADHD | | | | |
| --- | --- | --- | --- | --- |
| Characteristics | Odds Ratio | 95% CI | p-value | Imputation |
| Eviction Stress/Concern | 1.0347702 | 1.00-1.07 | 0.041 | PMM |
| Age * Eviction Stress/Concern | 0.9702400 | 0.94-1.00 | 0.042 | PMM |
| Eviction Stress/Concern | 1.0326364 | 1.00-1.07 | 0.067 | K-nearest neighbor |
| Age * Eviction Stress/Concern | 0.9643756 | 0.93-1.00 | 0.025 | K-nearest neighbor |
| Eviction Stress/Concern | 1.0311107 | 1.00-1.07 | 0.080 | Random Forest |
| Age * Eviction Stress/Concern | 0.9646819 | 0.93-1.00 | 0.026 | Random Forest |

Regarding behavioral problems, no statistical models using different types of imputed data revealed significant associations (as shown in Table S12). PMM imputation showed no significant relationships with either stress about eviction or housing loss (OR=1.01, CI=0.98-1.05, p=.433) or the age interaction (OR=1.00, CI=0.97-1.04, p=.799). Similarly, null results were found using K-nearest neighbor imputation (eviction: OR=1.00, CI=0.96-1.03, p=.876; age interaction: OR=1.00, CI=0.96-1.03, p=.772) and Random Forest imputation (eviction: OR=0.99, CI=0.96-1.03, p=.712; age interaction: OR=1.00, CI=0.96-1.03, p=.821).

Table S12.

| Associations with Behavioral Problems | | | | |
| --- | --- | --- | --- | --- |
| Characteristics | Odds Ratio | 95% CI | p-value | Imputation |
| Eviction Stress/Concern | 1.0138212 | 0.98-1.05 | 0.433 | PMM |
| Age * Eviction Stress/Concern | 1.0042064 | 0.97-1.04 | 0.799 | PMM |
| Eviction Stress/Concern | 0.9972104 | 0.96-1.03 | 0.876 | K-nearest neighbor |
| Age * Eviction Stress/Concern | 0.9950447 | 0.96-1.03 | 0.772 | K-nearest neighbor |
| Eviction Stress/Concern | 0.9934204 | 0.96-1.03 | 0.712 | Random Forest |
| Age * Eviction Stress/Concern | 0.9961493 | 0.96-1.03 | 0.821 | Random Forest |

### R Markdown (Analytic Code) Related to Analyses in the Main Manuscript

**# Load Required Libraries**

pacman::p_load(haven, tidyverse, sjPlot, ggstatsplot, interactions, ggstats, oddsratio, epiDisplay, Hmisc, gtsummary, easystats)

**\# Load Data File (SPSS Format)**

NSCH_Topical_DRC_CAHMI<-read_sav("2022 NSCH_Topical_DRC_CAHMI.sav")

**\# Recode Different Variables and Create Factors**

\# Recode and create factor variables for race/ethnicity
NSCH_Topical_DRC_CAHMI$raceASIA_22<-as.numeric(car::recode(NSCH_Topical_DRC_CAHMI$raceASIA_22,"2=1;1=2;3=3;4=4;5=5"))
NSCH_Topical_DRC_CAHMI$raceASIA_22_F<-factor(NSCH_Topical_DRC_CAHMI$raceASIA_22)

\# Recode medical diagnosis variable (0=No, 1=Yes, NA for missing)
NSCH_Topical_DRC_CAHMI$MEDB10ScrQ5_22_R<-as.numeric(car::recode(NSCH_Topical_DRC_CAHMI$MEDB10ScrQ5_22,"1=1;2=0;95=NA"))

\# Recode and standardize home eviction stress variable
NSCH_Topical_DRC_CAHMI$HomeEvic_22_R<-labelled::remove_labels(NSCH_Topical_DRC_CAHMI$HomeEvic_22)
NSCH_Topical_DRC_CAHMI$HomeEvic_22_R<-as.numeric(car::recode(NSCH_Topical_DRC_CAHMI$HomeEvic_22,"1=5;2=4;3=3;4=2;5=1"))
NSCH_Topical_DRC_CAHMI$HomeEvic_22_R_Z<-as.numeric(scale(NSCH_Topical_DRC_CAHMI$HomeEvic_22_R,scale=TRUE))

\# Recode sex variable (0=Male, 1=Female)
NSCH_Topical_DRC_CAHMI$sex_22_R<-as.numeric(car::recode(NSCH_Topical_DRC_CAHMI$sex_22,"1=0;2=1;99=NA"))
NSCH_Topical_DRC_CAHMI$sex_22_R<-labelled::remove_labels(NSCH_Topical_DRC_CAHMI$sex_22_R)

\# Recode homelessness variable (0=No, 1=Yes)
NSCH_Topical_DRC_CAHMI$EverHomeless_22_R<-as.numeric(car::recode(NSCH_Topical_DRC_CAHMI$EverHomeless_22,"1=1;2=0;3=NA;99=NA"))
NSCH_Topical_DRC_CAHMI$EverHomeless_22_R_F<-factor(NSCH_Topical_DRC_CAHMI$EverHomeless_22_R)

\# Recode low birth weight variable (0=No, 1=Yes)
NSCH_Topical_DRC_CAHMI$LowBWght_22_R<-as.numeric(car::recode(NSCH_Topical_DRC_CAHMI$LowBWght_22,"1=1;2=0;99=NA"))
NSCH_Topical_DRC_CAHMI$LowBWght_22_R_F<-factor(NSCH_Topical_DRC_CAHMI$LowBWght_22_R)

\# Recode places lived variable (0=0-2 times, 1=3 or more times)
NSCH_Topical_DRC_CAHMI$PlacesLived_22_R<-as.numeric(car::recode(NSCH_Topical_DRC_CAHMI$PlacesLived_22,"1=0;2=1;99=NA"))
NSCH_Topical_DRC_CAHMI$PlacesLived_22_R_F<-factor(NSCH_Topical_DRC_CAHMI$PlacesLived_22_R)

\# Recode premature birth variable (0=No, 1=Yes)
NSCH_Topical_DRC_CAHMI$BornPre_22_R<-as.numeric(car::recode(NSCH_Topical_DRC_CAHMI$BornPre_22,"1=1;2=0;99=NA"))
NSCH_Topical_DRC_CAHMI$BornPre_22_R_F<-factor(NSCH_Topical_DRC_CAHMI$BornPre_22_R)

\# Recode food insecurity variable
NSCH_Topical_DRC_CAHMI$FoodSit_22_R<-as.numeric(car::recode(NSCH_Topical_DRC_CAHMI$FoodSit_22,"99=NA"))
NSCH_Topical_DRC_CAHMI$FoodSit_22_R<-labelled::remove_labels(NSCH_Topical_DRC_CAHMI$FoodSit_22_R)

\# Create factor variables for family structure, adult education, and poverty level
NSCH_Topical_DRC_CAHMI$famstruct5_22_F<-factor(NSCH_Topical_DRC_CAHMI$famstruct5_22)
NSCH_Topical_DRC_CAHMI$AdultEduc_22_F<-factor(NSCH_Topical_DRC_CAHMI$AdultEduc_22)
NSCH_Topical_DRC_CAHMI$povlev4_22_F<-factor(NSCH_Topical_DRC_CAHMI$povlev4_22)

\# Clean and standardize child's age variable
NSCH_Topical_DRC_CAHMI$SC_AGE_YEARS<-labelled::remove_labels(NSCH_Topical_DRC_CAHMI$SC_AGE_YEARS); NSCH_Topical_DRC_CAHMI$SC_AGE_YEARS_Z<-as.numeric(scale(NSCH_Topical_DRC_CAHMI$SC_AGE_YEARS,scale=TRUE))

\# Recode ACEs (Adverse Childhood Experiences) variable
NSCH_Topical_DRC_CAHMI$ACE2more11_22_R<-as.numeric(car::recode(NSCH_Topical_DRC_CAHMI$ACE2more11_22,"99=NA"))
NSCH_Topical_DRC_CAHMI$ACE2more11_22_R_F<-as.factor(NSCH_Topical_DRC_CAHMI$ACE2more11_22_R)

\# Recode mother's physical and mental health variables
NSCH_Topical_DRC_CAHMI$MothPhyH_22_R<-as.numeric(car::recode(NSCH_Topical_DRC_CAHMI$MothPhyH_22,"95=NA;99=NA"))
NSCH_Topical_DRC_CAHMI$MothPhyH_22_R_F<-as.factor(NSCH_Topical_DRC_CAHMI$MothPhyH_22_R)
NSCH_Topical_DRC_CAHMI$MotherMH_22_R<-as.numeric(car::recode(NSCH_Topical_DRC_CAHMI$MotherMH_22,"95=NA;99=NA"))
NSCH_Topical_DRC_CAHMI$MotherMH_22_R_F<-as.factor(NSCH_Topical_DRC_CAHMI$MotherMH_22_R)

**\# Fixing Data Labels (Mainly to Make Tables Better Later)**

\# Create labeled factor variables with descriptive levels
\# Sex
NSCH_Topical_DRC_CAHMI$sex_22_R_L <- factor(NSCH_Topical_DRC_CAHMI$sex_22_R,
levels = c(0,1),
labels = c("Male", "Female"))
label(NSCH_Topical_DRC_CAHMI$sex_22_R_L)<-"Child's Sex Assigned at Birth"

\# Age labels
label(NSCH_Topical_DRC_CAHMI$SC_AGE_YEARS_Z)<-"Child's Age in Years"
label(NSCH_Topical_DRC_CAHMI$SC_AGE_YEARS)<-"Child's Age in Years"

\# Medical diagnosis label
NSCH_Topical_DRC_CAHMI$MEDB10ScrQ5_22_R_L <- factor(NSCH_Topical_DRC_CAHMI$MEDB10ScrQ5_22_R,
levels = c(0,1),
labels = c("No", "Yes"))
label(NSCH_Topical_DRC_CAHMI$MEDB10ScrQ5_22_R_L)<- "Any Diagnosis"

\# Family Structure label
NSCH_Topical_DRC_CAHMI$famstruct5_22_F_L <- factor(NSCH_Topical_DRC_CAHMI$famstruct5_22_F,
levels = c(1,2,3,4,5),
labels = c("Two parents, currently married","Two parents, not currently married","Single parent (mother or father)","Grandparent household","Other family type"))
label(NSCH_Topical_DRC_CAHMI$famstruct5_22_F_L)<- "Family Structure"

\# Adult Education label
NSCH_Topical_DRC_CAHMI$AdultEduc_22_F_L <- factor(NSCH_Topical_DRC_CAHMI$AdultEduc_22_F,
levels = c(1,2,3,4),
labels = c("Less than high school","High school or GED","Some college or technical school","College degree or higher"))
label(NSCH_Topical_DRC_CAHMI$AdultEduc_22_F_L)<- "Highest Level of Education in Household"

\# Race label
NSCH_Topical_DRC_CAHMI$raceASIA_22_F_L <- factor(NSCH_Topical_DRC_CAHMI$raceASIA_22_F,
levels = c(1,2,3,4,5),
labels = c("White, non-Hispanic","Hispanic","Black, non-Hispanic","Asian, non-Hispanic","Multi-racial, non-Hispanic"))
label(NSCH_Topical_DRC_CAHMI$raceASIA_22_F_L)<- "Child's Race/Ethnicity"

\# Poverty/SES in household label
NSCH_Topical_DRC_CAHMI$povlev4_22_F_L <- factor(NSCH_Topical_DRC_CAHMI$povlev4_22_F,
levels = c(1,2,3,4),
labels = c("0-99% FPL","100-199% FPL","200-399% FPL","400% FPL or greater"))
label(NSCH_Topical_DRC_CAHMI$povlev4_22_F_L)<- "Household Poverty Status"

\# Eviction Stress label
label(NSCH_Topical_DRC_CAHMI$HomeEvic_22_R_Z)<- "Eviction Stress/Concern"

\# Placed lived label
NSCH_Topical_DRC_CAHMI$PlacesLived_22_R_F<-labelled::remove_labels(NSCH_Topical_DRC_CAHMI$PlacesLived_22_R_F)
NSCH_Topical_DRC_CAHMI$PlacesLived_22_R_F_L <- factor(NSCH_Topical_DRC_CAHMI$PlacesLived_22_R_F,
levels = c(0,1),
labels = c("0-2 times","3 or more times"))
label(NSCH_Topical_DRC_CAHMI$PlacesLived_22_R_F_L)<- "Places Lived, Last Year"

\# Mother's physical health label
NSCH_Topical_DRC_CAHMI$MothPhyH_22_R_F<-labelled::remove_labels(NSCH_Topical_DRC_CAHMI$MothPhyH_22_R_F)
NSCH_Topical_DRC_CAHMI$MothPhyH_22_R_F_L <- factor(NSCH_Topical_DRC_CAHMI$MothPhyH_22_R_F,
levels = c(1,2,3),
labels = c("Excellent or very good", "Good","Fair or poor"))
label(NSCH_Topical_DRC_CAHMI$MothPhyH_22_R_F_L)<- "Mother's Physical health"

\# Mother's mental health label
NSCH_Topical_DRC_CAHMI$MotherMH_22_R_F<-labelled::remove_labels(NSCH_Topical_DRC_CAHMI$MotherMH_22_R_F)
NSCH_Topical_DRC_CAHMI$MotherMH_22_R_F_L <- factor(NSCH_Topical_DRC_CAHMI$MotherMH_22_R_F,
levels = c(1,2,3),
labels = c("Excellent or very good", "Good","Fair or poor"))
label(NSCH_Topical_DRC_CAHMI$MotherMH_22_R_F_L)<- "Mother's Mental health"

\# Homelessness label
NSCH_Topical_DRC_CAHMI$EverHomeless_22_R_F<-labelled::remove_labels(NSCH_Topical_DRC_CAHMI$EverHomeless_22_R_F)
NSCH_Topical_DRC_CAHMI$EverHomeless_22_R_F_L <- factor(NSCH_Topical_DRC_CAHMI$EverHomeless_22_R_F,
levels = c(0,1),
labels = c("No", "Yes"))
label(NSCH_Topical_DRC_CAHMI$EverHomeless_22_R_F_L)<- "Ever Homeless"

\# Low Birth Weight label
NSCH_Topical_DRC_CAHMI$LowBWght_22_R_F_L <- factor(NSCH_Topical_DRC_CAHMI$LowBWght_22_R_F,
levels = c(0,1),
labels = c("No", "Yes"))
label(NSCH_Topical_DRC_CAHMI$LowBWght_22_R_F_L)<- "Low Birth Weight"

\# Premature Birth label
NSCH_Topical_DRC_CAHMI$BornPre_22_R_F_L <- factor(NSCH_Topical_DRC_CAHMI$BornPre_22_R_F,
levels = c(0,1),
labels = c("No", "Yes"))
label(NSCH_Topical_DRC_CAHMI$BornPre_22_R_F_L)<- "Born Premature"

\# Food Insecurity label
NSCH_Topical_DRC_CAHMI$FoodSit_22_R<-labelled::remove_labels(NSCH_Topical_DRC_CAHMI$FoodSit_22_R)
label(NSCH_Topical_DRC_CAHMI$FoodSit_22_R)<- "Food Insecurity"

\# ACEs label
NSCH_Topical_DRC_CAHMI$ACE2more11_22_R_F<-labelled::remove_labels(NSCH_Topical_DRC_CAHMI$ACE2more11_22_R_F)
NSCH_Topical_DRC_CAHMI$ACE2more11_22_R_F_L <- factor(NSCH_Topical_DRC_CAHMI$ACE2more11_22_R_F,
levels = c(1,2,3),
labels = c("0 ACEs","1 ACE","2+ ACEs"))
label(NSCH_Topical_DRC_CAHMI$ACE2more11_22_R_F_L)<- "Adverse Childhood Experiences"

\# Parent/Caregiver Sex label
NSCH_Topical_DRC_CAHMI$A1_SEX_R<-as.numeric(car::recode(NSCH_Topical_DRC_CAHMI$A1_SEX,"1=0;2=1;99=NA"))
NSCH_Topical_DRC_CAHMI$A1_SEX_R<-labelled::remove_labels(NSCH_Topical_DRC_CAHMI$A1_SEX_R)
NSCH_Topical_DRC_CAHMI$A1_SEX_R_L <- factor(NSCH_Topical_DRC_CAHMI$A1_SEX_R,
 levels = c(0,1),
 labels = c("Male", "Female"))
label(NSCH_Topical_DRC_CAHMI$A1_SEX_R_L)<-"Parent's Sex Assigned at Birth"

\# Parent/Caregiver Age label
NSCH_Topical_DRC_CAHMI$A1_AGE_R<-as.numeric(car::recode(NSCH_Topical_DRC_CAHMI$A1_AGE,"999=NA"))
NSCH_Topical_DRC_CAHMI$A1_AGE_R<-labelled::remove_labels(NSCH_Topical_DRC_CAHMI$A1_AGE_R)
label(NSCH_Topical_DRC_CAHMI$A1_AGE_R)<-"Parent's Age in Years"

**\# Recoding Dependent Variables (Binary Indicators of Psychopathology)**

\# Depression Diagnosis label (0=Never; 0=Previously; 1=Current)
NSCH_Topical_DRC_CAHMI$depress_22_R_Alt<-car::recode(NSCH_Topical_DRC_CAHMI$depress_22,"1=0;2=0;3=1;95=NA;99=NA")
NSCH_Topical_DRC_CAHMI$depress_22_R_Alt<-labelled::remove_labels(NSCH_Topical_DRC_CAHMI$depress_22_R_Alt)

\# Anxiety Diagnosis label (0=Never; 0=Previously; 1=Current)
NSCH_Topical_DRC_CAHMI$anxiety_22_R_Alt<-car::recode(NSCH_Topical_DRC_CAHMI$anxiety_22,"1=0;2=0;3=1;95=NA;99=NA")
NSCH_Topical_DRC_CAHMI$anxiety_22_R_Alt<-labelled::remove_labels(NSCH_Topical_DRC_CAHMI$anxiety_22_R_Alt)

\# Behavioral Problems Diagnosis label (0=Never; 0=Previously; 1=Current)
NSCH_Topical_DRC_CAHMI$behavior_22_R_Alt<-car::recode(NSCH_Topical_DRC_CAHMI$behavior_22,"1=0;2=0;3=1;95=NA;99=NA")
NSCH_Topical_DRC_CAHMI$behavior_22_R_Alt<-labelled::remove_labels(NSCH_Topical_DRC_CAHMI$behavior_22_R_Alt)

\# ADHD Diagnosis label (0=Never; 0=Previously; 1=Current)
NSCH_Topical_DRC_CAHMI$ADHD_22_R_Alt<-car::recode(NSCH_Topical_DRC_CAHMI$ADHD_22,"1=0;2=0;3=1;95=NA;99=NA")
NSCH_Topical_DRC_CAHMI$ADHD_22_R_Alt<-labelled::remove_labels(NSCH_Topical_DRC_CAHMI$ADHD_22_R_Alt)

\# Autism Diagnosis label (0=Never; 0=Previously; 1=Current)
NSCH_Topical_DRC_CAHMI$autism_22_R_Alt<-car::recode(NSCH_Topical_DRC_CAHMI$autism_22,"1=0;2=0;3=1;95=NA;99=NA")
NSCH_Topical_DRC_CAHMI$autism_22_R_Alt<-labelled::remove_labels(NSCH_Topical_DRC_CAHMI$autism_22_R_Alt)

\# Developmental Delay Diagnosis label (0=Never; 0=Previously; 1=Current)
NSCH_Topical_DRC_CAHMI$DevDelay_22_R_Alt<-car::recode(NSCH_Topical_DRC_CAHMI$DevDelay_22,"1=0;2=0;3=1;95=NA;99=NA")
NSCH_Topical_DRC_CAHMI$DevDelay_22_R_Alt<-labelled::remove_labels(NSCH_Topical_DRC_CAHMI$DevDelay_22_R_Alt)

\# Intellectual Disability Diagnosis label (0=Never; 0=Previously; 1=Current)
NSCH_Topical_DRC_CAHMI$IntDisab_22_R_Alt<-car::recode(NSCH_Topical_DRC_CAHMI$IntDisab_22,"1=0;2=0;3=1;95=NA;99=NA")
NSCH_Topical_DRC_CAHMI$IntDisab_22_R_Alt<-labelled::remove_labels(NSCH_Topical_DRC_CAHMI$IntDisab_22_R_Alt)

\# Tourette's Disorder Diagnosis label (0=Never; 0=Previously; 1=Current)
NSCH_Topical_DRC_CAHMI$tourette_22_R_Alt<-car::recode(NSCH_Topical_DRC_CAHMI$tourette_22,"1=0;2=0;3=1;95=NA;99=NA")
NSCH_Topical_DRC_CAHMI$tourette_22_R_Alt<-labelled::remove_labels(NSCH_Topical_DRC_CAHMI$tourette_22_R_Alt)

\# Speech Disorder Diagnosis label (0=Never; 0=Previously; 1=Current)
NSCH_Topical_DRC_CAHMI$speech_22_R_Alt<-car::recode(NSCH_Topical_DRC_CAHMI$speech_22,"1=0;2=0;3=1;95=NA;99=NA")
NSCH_Topical_DRC_CAHMI$speech_22_R_Alt<-labelled::remove_labels(NSCH_Topical_DRC_CAHMI$speech_22_R_Alt)

\# Learning Disorder Diagnosis label (0=Never; 0=Previously; 1=Current)
NSCH_Topical_DRC_CAHMI$learning_22_R_Alt<-car::recode(NSCH_Topical_DRC_CAHMI$learning_22,"1=0;2=0;3=1;95=NA;99=NA")
NSCH_Topical_DRC_CAHMI$learning_22_R_Alt<-labelled::remove_labels(NSCH_Topical_DRC_CAHMI$learning_22_R_Alt)

**\# Create Analytic Sample, Complete Cases Only**

nrow(NSCH_Topical_DRC_CAHMI)

\## [1] 54103

NSCH_Topical_DRC_CAHMI_subset<-subset(NSCH_Topical_DRC_CAHMI,NSCH_Topical_DRC_CAHMI$MEDB10ScrQ5_22_R_L!='NA' & NSCH_Topical_DRC_CAHMI$raceASIA_22_F_L!='NA' & NSCH_Topical_DRC_CAHMI$famstruct5_22_F_L!='NA' & NSCH_Topical_DRC_CAHMI$AdultEduc_22_F_L!='NA' & NSCH_Topical_DRC_CAHMI$povlev4_22_F_L!='NA' & NSCH_Topical_DRC_CAHMI$sex_22_R_L!='NA' & NSCH_Topical_DRC_CAHMI$PlacesLived_22_R_F_L!='NA' & NSCH_Topical_DRC_CAHMI$MothPhyH_22_R_F_L!='NA' & NSCH_Topical_DRC_CAHMI$MotherMH_22_R_F_L!='NA' & NSCH_Topical_DRC_CAHMI$EverHomeless_22_R_F_L!='NA' & NSCH_Topical_DRC_CAHMI$LowBWght_22_R_F_L!='NA' & NSCH_Topical_DRC_CAHMI$BornPre_22_R_F_L!='NA' & NSCH_Topical_DRC_CAHMI$FoodSit_22_R!='NA' & NSCH_Topical_DRC_CAHMI$ACE2more11_22_R_F_L!='NA' & NSCH_Topical_DRC_CAHMI$SC_AGE_YEARS_Z!='NA' & NSCH_Topical_DRC_CAHMI$HomeEvic_22_R_Z!='NA' & NSCH_Topical_DRC_CAHMI$SC_AGE_YEARS_Z!='NA' & NSCH_Topical_DRC_CAHMI$depress_22_R_Alt!='NA' & NSCH_Topical_DRC_CAHMI$anxiety_22_R_Alt!='NA' & NSCH_Topical_DRC_CAHMI$ADHD_22_R_Alt!='NA' & NSCH_Topical_DRC_CAHMI$behavior_22_R_Alt!='NA')

nrow(NSCH_Topical_DRC_CAHMI_subset)

\## [1] 36638

**\# Create Summary Table (Table 01)**

\# Create summary tables

\# Define variables for full sample table
library(gtsummary)
vars <- c("A1_SEX_R_L","A1_AGE_R","sex_22_R_L", "SC_AGE_YEARS", "raceASIA_22_F_L", "povlev4_22_F_L", "famstruct5_22_F_L", "AdultEduc_22_F_L")
NSCH_Topical_DRC_CAHMI_vars_full <- NSCH_Topical_DRC_CAHMI[vars]
race_levels <- c("Asian, non-Hispanic", "Black, non-Hispanic", "Hispanic", "Multi-racial, non-Hispanic", "White, non-Hispanic")
NSCH_Topical_DRC_CAHMI_vars_full$raceASIA_22_F_L <- factor(NSCH_Topical_DRC_CAHMI_vars_full$raceASIA_22_F_L, levels = race_levels)

\# Create tables for full sample
full_tbl <- NSCH_Topical_DRC_CAHMI_vars_full %>%
 tbl_summary(
 statistic = all_continuous() ~ "{mean} ({sd})",
 digits = all_continuous() ~ 2,
 include = all_of(vars),
 label = list(
 raceASIA_22_F_L ~ "Child's Race/Ethnicity†"
 ),
 missing_text = "Missing" # Add this line to include missing value information
 ) %>%
 modify_footnote(
 update = list(
 stat_0 = "n (%); Mean (SD); † Multi-racial, non-Hispanic includes: American Indian or Alaska Native Non-Hispanic, Native Hawaiian and Other Pacific Islander Non-Hispanic, and other Multi-Race Non-Hispanic Groups"))

```
## Warning: The `update` argument of `modify_footnote()` is deprecated as of gtsummary
## 2.0.0.
## ℹ Use `modify_footnote(...)` input instead. Dynamic dots allow for syntax like
##   `modify_footnote(!!!list(...))`.
## ℹ The deprecated feature was likely used in the gtsummary package.
##   Please report the issue at <https://github.com/ddsjoberg/gtsummary/issues>.
## This warning is displayed once every 8 hours.
## Call `lifecycle::last_lifecycle_warnings()` to see where this warning was
## generated.
```

\# Define variables for analytic sample table
NSCH_Topical_DRC_CAHMI_vars_with_data<-NSCH_Topical_DRC_CAHMI_subset[vars]
NSCH_Topical_DRC_CAHMI_vars_with_data$raceASIA_22_F_L <- factor(NSCH_Topical_DRC_CAHMI_vars_with_data$raceASIA_22_F_L, levels = race_levels)

\# Create tables for analytic sample
analytic_tbl <- NSCH_Topical_DRC_CAHMI_vars_with_data %>%
 tbl_summary(
 statistic = all_continuous() ~ "{mean} ({sd})",
 digits = all_continuous() ~ 2,
 include = all_of(vars),
 label = list(
 raceASIA_22_F_L ~ "Child's Race/Ethnicity†"
 ),
 missing_text = "Missing" # Add this line to include missing value information
 ) %>%
 modify_footnote(
 update = list(
 stat_0 = "n (%); Mean (SD); † Multi-racial, non-Hispanic includes: American Indian or Alaska Native Non-Hispanic, Native Hawaiian and Other Pacific Islander Non-Hispanic, and other Multi-Race Non-Hispanic Groups"))

\# Merge tables and save
combined_table01<-tbl_merge(
 tbls = list(full_tbl, analytic_tbl),
 tab_spanner = c("**Full Sample**", "**Analytic Sample**")
)

combined_table01

|  | Full Sample | Analytic Sample |
| --- | --- | --- |
| Characteristic | N = 54,103*^1^* | N = 36,638*^1^* |
| Parent's Sex Assigned at Birth |  |  |
| Male | 16,478 (31%) | 10,345 (28%) |
| Female | 35,959 (69%) | 26,273 (72%) |
| Missing | 1,666 | 20 |
| Parent's Age in Years | 42.08 (9.50) | 42.38 (8.20) |
| Missing | 1,692 | 74 |
| Child's Sex Assigned at Birth |  |  |
| Male | 27,911 (52%) | 18,901 (52%) |
| Female | 26,192 (48%) | 17,737 (48%) |
| Child's Age in Years | 8.60 (5.31) | 9.91 (4.62) |
| Child's Race/Ethnicity† |  |  |
| Asian, non-Hispanic | 3,307 (6.1%) | 2,042 (5.6%) |
| Black, non-Hispanic | 3,292 (6.1%) | 1,884 (5.1%) |
| Hispanic | 8,370 (15%) | 5,298 (14%) |
| Multi-racial, non-Hispanic | 4,401 (8.1%) | 2,921 (8.0%) |
| White, non-Hispanic | 34,733 (64%) | 24,493 (67%) |
| Household Poverty Status |  |  |
| 0-99% FPL | 6,867 (13%) | 4,044 (11%) |
| 100-199% FPL | 8,629 (16%) | 5,447 (15%) |
| 200-399% FPL | 15,674 (29%) | 10,735 (29%) |
| 400% FPL or greater | 22,933 (42%) | 16,412 (45%) |
| Family Structure |  |  |
| Two parents, currently married | 37,133 (71%) | 28,293 (77%) |
| Two parents, not currently married | 3,084 (5.9%) | 2,101 (5.7%) |
| Single parent (mother or father) | 10,441 (20%) | 6,189 (17%) |
| Grandparent household | 1,412 (2.7%) | 4 (<0.1%) |
| Other family type | 483 (0.9%) | 51 (0.1%) |
| Missing | 1,550 |  |
| Highest Level of Education in Household |  |  |
| Less than high school | 1,433 (2.6%) | 746 (2.0%) |
| High school or GED | 7,046 (13%) | 4,080 (11%) |
| Some college or technical school | 11,394 (21%) | 7,549 (21%) |
| College degree or higher | 34,230 (63%) | 24,263 (66%) |
| *^1^*n (%); Mean (SD); † Multi-racial, non-Hispanic includes: American Indian or Alaska Native Non-Hispanic, Native Hawaiian and Other Pacific Islander Non-Hispanic, and other Multi-Race Non-Hispanic Groups | | |

\# Output table as Word doc
combined_table01 %>%
 gtsummary::as_gt() %>%
 gt::gtsave("Table1.docx")

**\# Analyses Related to Depression**

\# Load Library for Linear Mixed Effect Modeling
library(lme4)

```
## Loading required package: Matrix
##
## Attaching package: 'Matrix'
## The following objects are masked from 'package:tidyr':
##
##     expand, pack, unpack
```

\# Changing Name of Age Variable (for Ease Later in Figures, etc.)
NSCH_Topical_DRC_CAHMI_subset$Child_Age<-NSCH_Topical_DRC_CAHMI_subset$SC_AGE_YEARS_Z

\# Depression Base Model
Depression_Base_Adjusted_Age_Interact<-glmer(depress_22_R_Alt ~ raceASIA_22_F_L + famstruct5_22_F_L + AdultEduc_22_F_L + povlev4_22_F_L + sex_22_R_L + Child_Age * HomeEvic_22_R_Z + (1|FIPSST), family=binomial, data=NSCH_Topical_DRC_CAHMI_subset, control = glmerControl(optimizer = "bobyqa",optCtrl=list(maxfun=2e5)),nAGQ = 0)
summary(Depression_Base_Adjusted_Age_Interact)

```
## Generalized linear mixed model fit by maximum likelihood (Adaptive
##   Gauss-Hermite Quadrature, nAGQ = 0) [glmerMod]
##  Family: binomial  ( logit )
## Formula:
## depress_22_R_Alt ~ raceASIA_22_F_L + famstruct5_22_F_L + AdultEduc_22_F_L +
##     povlev4_22_F_L + sex_22_R_L + Child_Age * HomeEvic_22_R_Z +
##     (1 | FIPSST)
##    Data: NSCH_Topical_DRC_CAHMI_subset
## Control: glmerControl(optimizer = "bobyqa", optCtrl = list(maxfun = 2e+05))
##
##      AIC      BIC   logLik deviance df.resid
##  12245.0  12415.1  -6102.5  12205.0    36618
##
## Scaled residuals:
##     Min      1Q  Median      3Q     Max
## -1.4730 -0.2563 -0.1294 -0.0678 17.6459
##
## Random effects:
##  Groups Name        Variance Std.Dev.
##  FIPSST (Intercept) 0.03162  0.1778
## Number of obs: 36638, groups:  FIPSST, 51
##
## Fixed effects:
##                                                      Estimate Std. Error
## (Intercept)                                         -4.877513   0.202577
## raceASIA_22_F_LHispanic                             -0.265824   0.076239
## raceASIA_22_F_LBlack, non-Hispanic                  -1.046509   0.134748
## raceASIA_22_F_LAsian, non-Hispanic                  -1.213854   0.172924
## raceASIA_22_F_LMulti-racial, non-Hispanic           -0.001226   0.090933
## famstruct5_22_F_LTwo parents, not currently married  0.624971   0.101446
## famstruct5_22_F_LSingle parent (mother or father)    0.616550   0.061460
## famstruct5_22_F_LGrandparent household               2.149475   1.214254
## famstruct5_22_F_LOther family type                   1.718244   0.431858
## AdultEduc_22_F_LHigh school or GED                   0.590079   0.191015
## AdultEduc_22_F_LSome college or technical school     0.668515   0.188245
## AdultEduc_22_F_LCollege degree or higher             0.521238   0.189044
## povlev4_22_F_L100-199% FPL                          -0.054632   0.088821
## povlev4_22_F_L200-399% FPL                          -0.115847   0.085308
## povlev4_22_F_L400% FPL or greater                   -0.229612   0.090064
## sex_22_R_LFemale                                     0.633027   0.050607
## Child_Age                                            1.610984   0.044282
## HomeEvic_22_R_Z                                      0.299224   0.032009
## Child_Age:HomeEvic_22_R_Z                           -0.068379   0.028315
##                                                     z value Pr(>|z|)
## (Intercept)                                         -24.077  < 2e-16 ***
## raceASIA_22_F_LHispanic                              -3.487 0.000489 ***
## raceASIA_22_F_LBlack, non-Hispanic                   -7.766 8.07e-15 ***
## raceASIA_22_F_LAsian, non-Hispanic                   -7.020 2.23e-12 ***
## raceASIA_22_F_LMulti-racial, non-Hispanic            -0.013 0.989246
## famstruct5_22_F_LTwo parents, not currently married   6.161 7.25e-10 ***
## famstruct5_22_F_LSingle parent (mother or father)    10.032  < 2e-16 ***
## famstruct5_22_F_LGrandparent household                1.770 0.076693 .
## famstruct5_22_F_LOther family type                    3.979 6.93e-05 ***
## AdultEduc_22_F_LHigh school or GED                    3.089 0.002007 **
## AdultEduc_22_F_LSome college or technical school      3.551 0.000383 ***
## AdultEduc_22_F_LCollege degree or higher              2.757 0.005829 **
## povlev4_22_F_L100-199% FPL                           -0.615 0.538503
## povlev4_22_F_L200-399% FPL                           -1.358 0.174470
## povlev4_22_F_L400% FPL or greater                    -2.549 0.010790 *
## sex_22_R_LFemale                                     12.509  < 2e-16 ***
## Child_Age                                            36.380  < 2e-16 ***
## HomeEvic_22_R_Z                                       9.348  < 2e-16 ***
## Child_Age:HomeEvic_22_R_Z                            -2.415 0.015737 *
## ---
## Signif. codes:  0 '***' 0.001 '**' 0.01 '*' 0.05 '.' 0.1 ' ' 1
##
## Correlation matrix not shown by default, as p = 19 > 12.
## Use print(x, correlation=TRUE)  or
##     vcov(x)        if you need it
```

\# Simple Slope Work
modelbased::estimate_slopes(Depression_Base_Adjusted_Age_Interact, trend = "HomeEvic_22_R_Z", by ="Child_Age",nuisance=c("raceASIA_22_F_L","famstruct5_22_F_L","AdultEduc_22_F_L","povlev4_22_F_L","sex_22_R_L"),length = 100)

```
## Estimated Marginal Effects
##
## Child_Age | Coefficient |   SE |       95% CI |     z |  df |      p
## --------------------------------------------------------------------
## -1.06     |        0.37 | 0.06 | [0.26, 0.49] |  6.36 | Inf | < .001
## -1.03     |        0.37 | 0.06 | [0.26, 0.48] |  6.41 | Inf | < .001
## -1.00     |        0.37 | 0.06 | [0.26, 0.48] |  6.45 | Inf | < .001
## -0.98     |        0.37 | 0.06 | [0.26, 0.48] |  6.50 | Inf | < .001
## -0.95     |        0.36 | 0.06 | [0.26, 0.47] |  6.55 | Inf | < .001
## -0.92     |        0.36 | 0.05 | [0.25, 0.47] |  6.60 | Inf | < .001
## -0.90     |        0.36 | 0.05 | [0.25, 0.47] |  6.65 | Inf | < .001
## -0.87     |        0.36 | 0.05 | [0.25, 0.46] |  6.71 | Inf | < .001
## -0.84     |        0.36 | 0.05 | [0.25, 0.46] |  6.76 | Inf | < .001
## -0.82     |        0.36 | 0.05 | [0.25, 0.46] |  6.82 | Inf | < .001
## -0.79     |        0.35 | 0.05 | [0.25, 0.45] |  6.87 | Inf | < .001
## -0.76     |        0.35 | 0.05 | [0.25, 0.45] |  6.93 | Inf | < .001
## -0.74     |        0.35 | 0.05 | [0.25, 0.45] |  6.99 | Inf | < .001
## -0.71     |        0.35 | 0.05 | [0.25, 0.44] |  7.05 | Inf | < .001
## -0.68     |        0.35 | 0.05 | [0.25, 0.44] |  7.12 | Inf | < .001
## -0.66     |        0.34 | 0.05 | [0.25, 0.44] |  7.18 | Inf | < .001
## -0.63     |        0.34 | 0.05 | [0.25, 0.43] |  7.24 | Inf | < .001
## -0.60     |        0.34 | 0.05 | [0.25, 0.43] |  7.31 | Inf | < .001
## -0.58     |        0.34 | 0.05 | [0.25, 0.43] |  7.38 | Inf | < .001
## -0.55     |        0.34 | 0.05 | [0.25, 0.43] |  7.45 | Inf | < .001
## -0.52     |        0.33 | 0.04 | [0.25, 0.42] |  7.52 | Inf | < .001
## -0.50     |        0.33 | 0.04 | [0.25, 0.42] |  7.60 | Inf | < .001
## -0.47     |        0.33 | 0.04 | [0.25, 0.42] |  7.67 | Inf | < .001
## -0.44     |        0.33 | 0.04 | [0.25, 0.41] |  7.75 | Inf | < .001
## -0.42     |        0.33 | 0.04 | [0.25, 0.41] |  7.83 | Inf | < .001
## -0.39     |        0.33 | 0.04 | [0.25, 0.41] |  7.91 | Inf | < .001
## -0.36     |        0.32 | 0.04 | [0.24, 0.40] |  7.99 | Inf | < .001
## -0.34     |        0.32 | 0.04 | [0.24, 0.40] |  8.08 | Inf | < .001
## -0.31     |        0.32 | 0.04 | [0.24, 0.40] |  8.17 | Inf | < .001
## -0.28     |        0.32 | 0.04 | [0.24, 0.39] |  8.26 | Inf | < .001
## -0.26     |        0.32 | 0.04 | [0.24, 0.39] |  8.35 | Inf | < .001
## -0.23     |        0.31 | 0.04 | [0.24, 0.39] |  8.44 | Inf | < .001
## -0.20     |        0.31 | 0.04 | [0.24, 0.39] |  8.54 | Inf | < .001
## -0.18     |        0.31 | 0.04 | [0.24, 0.38] |  8.64 | Inf | < .001
## -0.15     |        0.31 | 0.04 | [0.24, 0.38] |  8.74 | Inf | < .001
## -0.12     |        0.31 | 0.03 | [0.24, 0.38] |  8.84 | Inf | < .001
## -0.10     |        0.31 | 0.03 | [0.24, 0.37] |  8.95 | Inf | < .001
## -0.07     |        0.30 | 0.03 | [0.24, 0.37] |  9.05 | Inf | < .001
## -0.04     |        0.30 | 0.03 | [0.24, 0.37] |  9.16 | Inf | < .001
## -0.02     |        0.30 | 0.03 | [0.24, 0.36] |  9.28 | Inf | < .001
## 9.73e-03  |        0.30 | 0.03 | [0.24, 0.36] |  9.39 | Inf | < .001
## 0.04      |        0.30 | 0.03 | [0.24, 0.36] |  9.51 | Inf | < .001
## 0.06      |        0.29 | 0.03 | [0.23, 0.35] |  9.63 | Inf | < .001
## 0.09      |        0.29 | 0.03 | [0.23, 0.35] |  9.75 | Inf | < .001
## 0.12      |        0.29 | 0.03 | [0.23, 0.35] |  9.87 | Inf | < .001
## 0.14      |        0.29 | 0.03 | [0.23, 0.35] |  9.99 | Inf | < .001
## 0.17      |        0.29 | 0.03 | [0.23, 0.34] | 10.12 | Inf | < .001
## 0.20      |        0.29 | 0.03 | [0.23, 0.34] | 10.25 | Inf | < .001
## 0.22      |        0.28 | 0.03 | [0.23, 0.34] | 10.37 | Inf | < .001
## 0.25      |        0.28 | 0.03 | [0.23, 0.33] | 10.50 | Inf | < .001
## 0.28      |        0.28 | 0.03 | [0.23, 0.33] | 10.63 | Inf | < .001
## 0.30      |        0.28 | 0.03 | [0.23, 0.33] | 10.76 | Inf | < .001
## 0.33      |        0.28 | 0.03 | [0.23, 0.33] | 10.89 | Inf | < .001
## 0.36      |        0.27 | 0.02 | [0.23, 0.32] | 11.01 | Inf | < .001
## 0.38      |        0.27 | 0.02 | [0.22, 0.32] | 11.14 | Inf | < .001
## 0.41      |        0.27 | 0.02 | [0.22, 0.32] | 11.26 | Inf | < .001
## 0.44      |        0.27 | 0.02 | [0.22, 0.32] | 11.38 | Inf | < .001
## 0.46      |        0.27 | 0.02 | [0.22, 0.31] | 11.49 | Inf | < .001
## 0.49      |        0.27 | 0.02 | [0.22, 0.31] | 11.60 | Inf | < .001
## 0.52      |        0.26 | 0.02 | [0.22, 0.31] | 11.70 | Inf | < .001
## 0.54      |        0.26 | 0.02 | [0.22, 0.31] | 11.80 | Inf | < .001
## 0.57      |        0.26 | 0.02 | [0.22, 0.30] | 11.88 | Inf | < .001
## 0.60      |        0.26 | 0.02 | [0.22, 0.30] | 11.96 | Inf | < .001
## 0.62      |        0.26 | 0.02 | [0.21, 0.30] | 12.03 | Inf | < .001
## 0.65      |        0.25 | 0.02 | [0.21, 0.30] | 12.08 | Inf | < .001
## 0.68      |        0.25 | 0.02 | [0.21, 0.29] | 12.12 | Inf | < .001
## 0.70      |        0.25 | 0.02 | [0.21, 0.29] | 12.15 | Inf | < .001
## 0.73      |        0.25 | 0.02 | [0.21, 0.29] | 12.16 | Inf | < .001
## 0.76      |        0.25 | 0.02 | [0.21, 0.29] | 12.16 | Inf | < .001
## 0.78      |        0.25 | 0.02 | [0.21, 0.29] | 12.14 | Inf | < .001
## 0.81      |        0.24 | 0.02 | [0.20, 0.28] | 12.10 | Inf | < .001
## 0.84      |        0.24 | 0.02 | [0.20, 0.28] | 12.05 | Inf | < .001
## 0.86      |        0.24 | 0.02 | [0.20, 0.28] | 11.98 | Inf | < .001
## 0.89      |        0.24 | 0.02 | [0.20, 0.28] | 11.89 | Inf | < .001
## 0.92      |        0.24 | 0.02 | [0.20, 0.28] | 11.79 | Inf | < .001
## 0.94      |        0.23 | 0.02 | [0.20, 0.27] | 11.67 | Inf | < .001
## 0.97      |        0.23 | 0.02 | [0.19, 0.27] | 11.53 | Inf | < .001
## 1.00      |        0.23 | 0.02 | [0.19, 0.27] | 11.38 | Inf | < .001
## 1.02      |        0.23 | 0.02 | [0.19, 0.27] | 11.22 | Inf | < .001
## 1.05      |        0.23 | 0.02 | [0.19, 0.27] | 11.04 | Inf | < .001
## 1.08      |        0.23 | 0.02 | [0.18, 0.27] | 10.86 | Inf | < .001
## 1.10      |        0.22 | 0.02 | [0.18, 0.27] | 10.66 | Inf | < .001
## 1.13      |        0.22 | 0.02 | [0.18, 0.26] | 10.46 | Inf | < .001
## 1.16      |        0.22 | 0.02 | [0.18, 0.26] | 10.25 | Inf | < .001
## 1.18      |        0.22 | 0.02 | [0.18, 0.26] | 10.03 | Inf | < .001
## 1.21      |        0.22 | 0.02 | [0.17, 0.26] |  9.81 | Inf | < .001
## 1.24      |        0.21 | 0.02 | [0.17, 0.26] |  9.58 | Inf | < .001
## 1.26      |        0.21 | 0.02 | [0.17, 0.26] |  9.36 | Inf | < .001
## 1.29      |        0.21 | 0.02 | [0.17, 0.26] |  9.13 | Inf | < .001
## 1.32      |        0.21 | 0.02 | [0.16, 0.26] |  8.90 | Inf | < .001
## 1.34      |        0.21 | 0.02 | [0.16, 0.25] |  8.68 | Inf | < .001
## 1.37      |        0.21 | 0.02 | [0.16, 0.25] |  8.45 | Inf | < .001
## 1.40      |        0.20 | 0.02 | [0.16, 0.25] |  8.23 | Inf | < .001
## 1.42      |        0.20 | 0.03 | [0.15, 0.25] |  8.01 | Inf | < .001
## 1.45      |        0.20 | 0.03 | [0.15, 0.25] |  7.79 | Inf | < .001
## 1.48      |        0.20 | 0.03 | [0.15, 0.25] |  7.58 | Inf | < .001
## 1.50      |        0.20 | 0.03 | [0.14, 0.25] |  7.37 | Inf | < .001
## 1.53      |        0.19 | 0.03 | [0.14, 0.25] |  7.17 | Inf | < .001
## 1.56      |        0.19 | 0.03 | [0.14, 0.25] |  6.97 | Inf | < .001
## 1.58      |        0.19 | 0.03 | [0.14, 0.25] |  6.78 | Inf | < .001
## Marginal effects estimated for HomeEvic_22_R_Z
```

\# Depression Strict Model
Depression_Stringent_Adjusted_Age_Interact<- glmer(depress_22_R_Alt ~ raceASIA_22_F_L + famstruct5_22_F_L + AdultEduc_22_F_L + povlev4_22_F_L + sex_22_R_L + PlacesLived_22_R_F_L + EverHomeless_22_R_F_L + MotherMH_22_R_F_L + LowBWght_22_R_F_L + BornPre_22_R_F_L + FoodSit_22_R + ACE2more11_22_R_F_L + Child_Age * HomeEvic_22_R_Z + (1|FIPSST), family=binomial, data=NSCH_Topical_DRC_CAHMI_subset, control = glmerControl(optimizer = "bobyqa",optCtrl=list(maxfun=2e5)),nAGQ = 0)
summary(Depression_Stringent_Adjusted_Age_Interact)

```
## Generalized linear mixed model fit by maximum likelihood (Adaptive
##   Gauss-Hermite Quadrature, nAGQ = 0) [glmerMod]
##  Family: binomial  ( logit )
## Formula:
## depress_22_R_Alt ~ raceASIA_22_F_L + famstruct5_22_F_L + AdultEduc_22_F_L +
##     povlev4_22_F_L + sex_22_R_L + PlacesLived_22_R_F_L + EverHomeless_22_R_F_L +
##     MotherMH_22_R_F_L + LowBWght_22_R_F_L + BornPre_22_R_F_L +
##     FoodSit_22_R + ACE2more11_22_R_F_L + Child_Age * HomeEvic_22_R_Z +
##     (1 | FIPSST)
##    Data: NSCH_Topical_DRC_CAHMI_subset
## Control: glmerControl(optimizer = "bobyqa", optCtrl = list(maxfun = 2e+05))
##
##      AIC      BIC   logLik deviance df.resid
##  11107.0  11353.8  -5524.5  11049.0    36609
##
## Scaled residuals:
##     Min      1Q  Median      3Q     Max
## -1.7305 -0.2162 -0.1079 -0.0532 17.4918
##
## Random effects:
##  Groups Name        Variance Std.Dev.
##  FIPSST (Intercept) 0.02094  0.1447
## Number of obs: 36638, groups:  FIPSST, 51
##
## Fixed effects:
##                                                     Estimate Std. Error z value
## (Intercept)                                         -6.01643    0.22956 -26.208
## raceASIA_22_F_LHispanic                             -0.28702    0.07829  -3.666
## raceASIA_22_F_LBlack, non-Hispanic                  -0.96346    0.13809  -6.977
## raceASIA_22_F_LAsian, non-Hispanic                  -0.96912    0.17625  -5.498
## raceASIA_22_F_LMulti-racial, non-Hispanic           -0.20280    0.09442  -2.148
## famstruct5_22_F_LTwo parents, not currently married  0.06298    0.10772   0.585
## famstruct5_22_F_LSingle parent (mother or father)   -0.08715    0.06761  -1.289
## famstruct5_22_F_LGrandparent household               1.76707    1.26594   1.396
## famstruct5_22_F_LOther family type                   0.78429    0.45464   1.725
## AdultEduc_22_F_LHigh school or GED                   0.44581    0.20057   2.223
## AdultEduc_22_F_LSome college or technical school     0.48535    0.19797   2.452
## AdultEduc_22_F_LCollege degree or higher             0.51524    0.19901   2.589
## povlev4_22_F_L100-199% FPL                          -0.10636    0.09321  -1.141
## povlev4_22_F_L200-399% FPL                          -0.03863    0.08973  -0.430
## povlev4_22_F_L400% FPL or greater                    0.02755    0.09605   0.287
## sex_22_R_LFemale                                     0.64870    0.05267  12.316
## PlacesLived_22_R_F_L3 or more times                  0.62655    0.13942   4.494
## EverHomeless_22_R_F_LYes                             0.35056    0.12255   2.861
## MotherMH_22_R_F_LGood                                0.80027    0.05895  13.575
## MotherMH_22_R_F_LFair or poor                        1.19277    0.08047  14.822
## LowBWght_22_R_F_LYes                                 0.01558    0.10162   0.153
## BornPre_22_R_F_LYes                                  0.08998    0.09068   0.992
## FoodSit_22_R                                         0.14658    0.04854   3.020
## ACE2more11_22_R_F_L1 ACE                             0.60172    0.07465   8.060
## ACE2more11_22_R_F_L2+ ACEs                           1.57950    0.07169  22.033
## Child_Age                                            1.58931    0.04712  33.726
## HomeEvic_22_R_Z                                      0.09337    0.03523   2.650
## Child_Age:HomeEvic_22_R_Z                           -0.07575    0.03023  -2.505
##                                                     Pr(>|z|)
## (Intercept)                                          < 2e-16 ***
## raceASIA_22_F_LHispanic                             0.000246 ***
## raceASIA_22_F_LBlack, non-Hispanic                  3.02e-12 ***
## raceASIA_22_F_LAsian, non-Hispanic                  3.83e-08 ***
## raceASIA_22_F_LMulti-racial, non-Hispanic           0.031723 *
## famstruct5_22_F_LTwo parents, not currently married 0.558785
## famstruct5_22_F_LSingle parent (mother or father)   0.197371
## famstruct5_22_F_LGrandparent household              0.162759
## famstruct5_22_F_LOther family type                  0.084517 .
## AdultEduc_22_F_LHigh school or GED                  0.026233 *
## AdultEduc_22_F_LSome college or technical school    0.014222 *
## AdultEduc_22_F_LCollege degree or higher            0.009625 **
## povlev4_22_F_L100-199% FPL                          0.253880
## povlev4_22_F_L200-399% FPL                          0.666863
## povlev4_22_F_L400% FPL or greater                   0.774198
## sex_22_R_LFemale                                     < 2e-16 ***
## PlacesLived_22_R_F_L3 or more times                 6.99e-06 ***
## EverHomeless_22_R_F_LYes                            0.004229 **
## MotherMH_22_R_F_LGood                                < 2e-16 ***
## MotherMH_22_R_F_LFair or poor                        < 2e-16 ***
## LowBWght_22_R_F_LYes                                0.878170
## BornPre_22_R_F_LYes                                 0.321047
## FoodSit_22_R                                        0.002530 **
## ACE2more11_22_R_F_L1 ACE                            7.60e-16 ***
## ACE2more11_22_R_F_L2+ ACEs                           < 2e-16 ***
## Child_Age                                            < 2e-16 ***
## HomeEvic_22_R_Z                                     0.008041 **
## Child_Age:HomeEvic_22_R_Z                           0.012237 *
## ---
## Signif. codes:  0 '***' 0.001 '**' 0.01 '*' 0.05 '.' 0.1 ' ' 1
##
## Correlation matrix not shown by default, as p = 28 > 12.
## Use print(x, correlation=TRUE)  or
##     vcov(x)        if you need it
```

\#
modelbased::estimate_slopes(Depression_Stringent_Adjusted_Age_Interact, trend = "HomeEvic_22_R_Z", by = "Child_Age",nuisance=c("raceASIA_22_F_L","famstruct5_22_F_L","AdultEduc_22_F_L","povlev4_22_F_L","sex_22_R_L","PlacesLived_22_R_F_L","MotherMH_22_R_F_L","EverHomeless_22_R_F_L","LowBWght_22_R_F_L","BornPre_22_R_F_L","FoodSit_22_R","ACE2more11_22_R_F_L"), length = 100)

```
## Estimated Marginal Effects
##
## Child_Age | Coefficient |   SE |        95% CI |         z |  df |     p
## ------------------------------------------------------------------------
## -1.06     |        0.17 | 0.06 | [ 0.05, 0.30] |      2.76 | Inf | 0.006
## -1.03     |        0.17 | 0.06 | [ 0.05, 0.29] |      2.76 | Inf | 0.006
## -1.00     |        0.17 | 0.06 | [ 0.05, 0.29] |      2.76 | Inf | 0.006
## -0.98     |        0.17 | 0.06 | [ 0.05, 0.29] |      2.76 | Inf | 0.006
## -0.95     |        0.17 | 0.06 | [ 0.05, 0.28] |      2.76 | Inf | 0.006
## -0.92     |        0.16 | 0.06 | [ 0.05, 0.28] |      2.76 | Inf | 0.006
## -0.90     |        0.16 | 0.06 | [ 0.05, 0.28] |      2.76 | Inf | 0.006
## -0.87     |        0.16 | 0.06 | [ 0.05, 0.27] |      2.76 | Inf | 0.006
## -0.84     |        0.16 | 0.06 | [ 0.05, 0.27] |      2.76 | Inf | 0.006
## -0.82     |        0.16 | 0.06 | [ 0.05, 0.27] |      2.76 | Inf | 0.006
## -0.79     |        0.15 | 0.06 | [ 0.04, 0.26] |      2.76 | Inf | 0.006
## -0.76     |        0.15 | 0.05 | [ 0.04, 0.26] |      2.76 | Inf | 0.006
## -0.74     |        0.15 | 0.05 | [ 0.04, 0.25] |      2.76 | Inf | 0.006
## -0.71     |        0.15 | 0.05 | [ 0.04, 0.25] |      2.76 | Inf | 0.006
## -0.68     |        0.15 | 0.05 | [ 0.04, 0.25] |      2.76 | Inf | 0.006
## -0.66     |        0.14 | 0.05 | [ 0.04, 0.24] |      2.76 | Inf | 0.006
## -0.63     |        0.14 | 0.05 | [ 0.04, 0.24] |      2.76 | Inf | 0.006
## -0.60     |        0.14 | 0.05 | [ 0.04, 0.24] |      2.76 | Inf | 0.006
## -0.58     |        0.14 | 0.05 | [ 0.04, 0.23] |      2.76 | Inf | 0.006
## -0.55     |        0.14 | 0.05 | [ 0.04, 0.23] |      2.76 | Inf | 0.006
## -0.52     |        0.13 | 0.05 | [ 0.04, 0.23] |      2.76 | Inf | 0.006
## -0.50     |        0.13 | 0.05 | [ 0.04, 0.22] |      2.75 | Inf | 0.006
## -0.47     |        0.13 | 0.05 | [ 0.04, 0.22] |      2.75 | Inf | 0.006
## -0.44     |        0.13 | 0.05 | [ 0.04, 0.22] |      2.75 | Inf | 0.006
## -0.42     |        0.12 | 0.05 | [ 0.04, 0.21] |      2.75 | Inf | 0.006
## -0.39     |        0.12 | 0.04 | [ 0.04, 0.21] |      2.75 | Inf | 0.006
## -0.36     |        0.12 | 0.04 | [ 0.03, 0.21] |      2.74 | Inf | 0.006
## -0.34     |        0.12 | 0.04 | [ 0.03, 0.20] |      2.74 | Inf | 0.006
## -0.31     |        0.12 | 0.04 | [ 0.03, 0.20] |      2.74 | Inf | 0.006
## -0.28     |        0.11 | 0.04 | [ 0.03, 0.20] |      2.73 | Inf | 0.006
## -0.26     |        0.11 | 0.04 | [ 0.03, 0.19] |      2.73 | Inf | 0.006
## -0.23     |        0.11 | 0.04 | [ 0.03, 0.19] |      2.72 | Inf | 0.007
## -0.20     |        0.11 | 0.04 | [ 0.03, 0.19] |      2.72 | Inf | 0.007
## -0.18     |        0.11 | 0.04 | [ 0.03, 0.18] |      2.71 | Inf | 0.007
## -0.15     |        0.10 | 0.04 | [ 0.03, 0.18] |      2.70 | Inf | 0.007
## -0.12     |        0.10 | 0.04 | [ 0.03, 0.18] |      2.70 | Inf | 0.007
## -0.10     |        0.10 | 0.04 | [ 0.03, 0.17] |      2.69 | Inf | 0.007
## -0.07     |        0.10 | 0.04 | [ 0.03, 0.17] |      2.68 | Inf | 0.007
## -0.04     |        0.10 | 0.04 | [ 0.03, 0.17] |      2.67 | Inf | 0.008
## -0.02     |        0.09 | 0.04 | [ 0.02, 0.16] |      2.66 | Inf | 0.008
## 9.73e-03  |        0.09 | 0.04 | [ 0.02, 0.16] |      2.65 | Inf | 0.008
## 0.04      |        0.09 | 0.03 | [ 0.02, 0.16] |      2.63 | Inf | 0.008
## 0.06      |        0.09 | 0.03 | [ 0.02, 0.15] |      2.62 | Inf | 0.009
## 0.09      |        0.09 | 0.03 | [ 0.02, 0.15] |      2.60 | Inf | 0.009
## 0.12      |        0.08 | 0.03 | [ 0.02, 0.15] |      2.59 | Inf | 0.010
## 0.14      |        0.08 | 0.03 | [ 0.02, 0.15] |      2.57 | Inf | 0.010
## 0.17      |        0.08 | 0.03 | [ 0.02, 0.14] |      2.55 | Inf | 0.011
## 0.20      |        0.08 | 0.03 | [ 0.02, 0.14] |      2.53 | Inf | 0.011
## 0.22      |        0.08 | 0.03 | [ 0.02, 0.14] |      2.51 | Inf | 0.012
## 0.25      |        0.07 | 0.03 | [ 0.02, 0.13] |      2.48 | Inf | 0.013
## 0.28      |        0.07 | 0.03 | [ 0.01, 0.13] |      2.46 | Inf | 0.014
## 0.30      |        0.07 | 0.03 | [ 0.01, 0.13] |      2.43 | Inf | 0.015
## 0.33      |        0.07 | 0.03 | [ 0.01, 0.12] |      2.40 | Inf | 0.016
## 0.36      |        0.07 | 0.03 | [ 0.01, 0.12] |      2.37 | Inf | 0.018
## 0.38      |        0.06 | 0.03 | [ 0.01, 0.12] |      2.33 | Inf | 0.020
## 0.41      |        0.06 | 0.03 | [ 0.01, 0.12] |      2.29 | Inf | 0.022
## 0.44      |        0.06 | 0.03 | [ 0.01, 0.11] |      2.25 | Inf | 0.024
## 0.46      |        0.06 | 0.03 | [ 0.01, 0.11] |      2.21 | Inf | 0.027
## 0.49      |        0.06 | 0.03 | [ 0.01, 0.11] |      2.16 | Inf | 0.030
## 0.52      |        0.05 | 0.03 | [ 0.00, 0.10] |      2.11 | Inf | 0.034
## 0.54      |        0.05 | 0.03 | [ 0.00, 0.10] |      2.06 | Inf | 0.039
## 0.57      |        0.05 | 0.03 | [ 0.00, 0.10] |      2.01 | Inf | 0.045
## 0.60      |        0.05 | 0.02 | [ 0.00, 0.10] |      1.95 | Inf | 0.051
## 0.62      |        0.05 | 0.02 | [ 0.00, 0.09] |      1.89 | Inf | 0.059
## 0.65      |        0.04 | 0.02 | [ 0.00, 0.09] |      1.82 | Inf | 0.068
## 0.68      |        0.04 | 0.02 | [ 0.00, 0.09] |      1.76 | Inf | 0.079
## 0.70      |        0.04 | 0.02 | [-0.01, 0.09] |      1.68 | Inf | 0.092
## 0.73      |        0.04 | 0.02 | [-0.01, 0.08] |      1.61 | Inf | 0.107
## 0.76      |        0.04 | 0.02 | [-0.01, 0.08] |      1.53 | Inf | 0.125
## 0.78      |        0.03 | 0.02 | [-0.01, 0.08] |      1.46 | Inf | 0.145
## 0.81      |        0.03 | 0.02 | [-0.01, 0.08] |      1.37 | Inf | 0.169
## 0.84      |        0.03 | 0.02 | [-0.02, 0.08] |      1.29 | Inf | 0.197
## 0.86      |        0.03 | 0.02 | [-0.02, 0.07] |      1.21 | Inf | 0.228
## 0.89      |        0.03 | 0.02 | [-0.02, 0.07] |      1.12 | Inf | 0.263
## 0.92      |        0.02 | 0.02 | [-0.02, 0.07] |      1.03 | Inf | 0.302
## 0.94      |        0.02 | 0.02 | [-0.02, 0.07] |      0.94 | Inf | 0.346
## 0.97      |        0.02 | 0.02 | [-0.03, 0.07] |      0.85 | Inf | 0.394
## 1.00      |        0.02 | 0.02 | [-0.03, 0.06] |      0.76 | Inf | 0.445
## 1.02      |        0.02 | 0.02 | [-0.03, 0.06] |      0.67 | Inf | 0.501
## 1.05      |        0.01 | 0.02 | [-0.03, 0.06] |      0.58 | Inf | 0.559
## 1.08      |        0.01 | 0.02 | [-0.04, 0.06] |      0.50 | Inf | 0.620
## 1.10      |    9.89e-03 | 0.02 | [-0.04, 0.06] |      0.41 | Inf | 0.683
## 1.13      |    7.87e-03 | 0.02 | [-0.04, 0.06] |      0.32 | Inf | 0.748
## 1.16      |    5.85e-03 | 0.02 | [-0.04, 0.05] |      0.24 | Inf | 0.813
## 1.18      |    3.84e-03 | 0.03 | [-0.05, 0.05] |      0.15 | Inf | 0.878
## 1.21      |    1.82e-03 | 0.03 | [-0.05, 0.05] |      0.07 | Inf | 0.943
## 1.24      |   -2.01e-04 | 0.03 | [-0.05, 0.05] | -7.81e-03 | Inf | 0.994
## 1.26      |   -2.22e-03 | 0.03 | [-0.05, 0.05] |     -0.09 | Inf | 0.932
## 1.29      |   -4.24e-03 | 0.03 | [-0.06, 0.05] |     -0.16 | Inf | 0.873
## 1.32      |   -6.25e-03 | 0.03 | [-0.06, 0.05] |     -0.23 | Inf | 0.815
## 1.34      |   -8.27e-03 | 0.03 | [-0.06, 0.05] |     -0.30 | Inf | 0.761
## 1.37      |       -0.01 | 0.03 | [-0.06, 0.04] |     -0.37 | Inf | 0.710
## 1.40      |       -0.01 | 0.03 | [-0.07, 0.04] |     -0.44 | Inf | 0.661
## 1.42      |       -0.01 | 0.03 | [-0.07, 0.04] |     -0.50 | Inf | 0.616
## 1.45      |       -0.02 | 0.03 | [-0.07, 0.04] |     -0.56 | Inf | 0.573
## 1.48      |       -0.02 | 0.03 | [-0.08, 0.04] |     -0.62 | Inf | 0.534
## 1.50      |       -0.02 | 0.03 | [-0.08, 0.04] |     -0.68 | Inf | 0.497
## 1.53      |       -0.02 | 0.03 | [-0.08, 0.04] |     -0.73 | Inf | 0.463
## 1.56      |       -0.02 | 0.03 | [-0.09, 0.04] |     -0.79 | Inf | 0.432
## 1.58      |       -0.03 | 0.03 | [-0.09, 0.04] |     -0.84 | Inf | 0.403
## Marginal effects estimated for HomeEvic_22_R_Z
```

**\# Table Making Related to Depression (Table 02)**

\# Table with Depression Base Model
MDD_base_table <- tbl_regression(Depression_Base_Adjusted_Age_Interact, exponentiate = TRUE, label = list(raceASIA_22_F_L ~ "Race/Ethnicity")) %>% bold_p()

\# Table with Depression Strict Model
MDD_stringent_table <- tbl_regression(Depression_Stringent_Adjusted_Age_Interact, exponentiate = TRUE,label = list(raceASIA_22_F_L ~ "Race/Ethnicity"))%>% bold_p()

\# Combining Depression Tables
MDD_combined_table<-tbl_merge(
 tbls = list(MDD_base_table, MDD_stringent_table),
 tab_spanner = c("**Base Model**", "**Stringent Model**")
)

MDD_combined_table

|  | Base Model | | | Stringent Model | | |
| --- | --- | --- | --- | --- | --- | --- |
| Characteristic | OR*^1^* | 95% CI*^1^* | p-value | OR*^1^* | 95% CI*^1^* | p-value |
| Race/Ethnicity |  |  |  |  |  |  |
| White, non-Hispanic | — | — |  | — | — |  |
| Hispanic | 0.77 | 0.66, 0.89 | <0.001 | 0.75 | 0.64, 0.87 | <0.001 |
| Black, non-Hispanic | 0.35 | 0.27, 0.46 | <0.001 | 0.38 | 0.29, 0.50 | <0.001 |
| Asian, non-Hispanic | 0.30 | 0.21, 0.42 | <0.001 | 0.38 | 0.27, 0.54 | <0.001 |
| Multi-racial, non-Hispanic | 1.00 | 0.84, 1.19 | >0.9 | 0.82 | 0.68, 0.98 | 0.032 |
| Family Structure |  |  |  |  |  |  |
| Two parents, currently married | — | — |  | — | — |  |
| Two parents, not currently married | 1.87 | 1.53, 2.28 | <0.001 | 1.07 | 0.86, 1.32 | 0.6 |
| Single parent (mother or father) | 1.85 | 1.64, 2.09 | <0.001 | 0.92 | 0.80, 1.05 | 0.2 |
| Grandparent household | 8.58 | 0.79, 92.7 | 0.077 | 5.85 | 0.49, 70.0 | 0.2 |
| Other family type | 5.57 | 2.39, 13.0 | <0.001 | 2.19 | 0.90, 5.34 | 0.085 |
| Highest Level of Education in Household |  |  |  |  |  |  |
| Less than high school | — | — |  | — | — |  |
| High school or GED | 1.80 | 1.24, 2.62 | 0.002 | 1.56 | 1.05, 2.31 | 0.026 |
| Some college or technical school | 1.95 | 1.35, 2.82 | <0.001 | 1.62 | 1.10, 2.39 | 0.014 |
| College degree or higher | 1.68 | 1.16, 2.44 | 0.006 | 1.67 | 1.13, 2.47 | 0.010 |
| Household Poverty Status |  |  |  |  |  |  |
| 0-99% FPL | — | — |  | — | — |  |
| 100-199% FPL | 0.95 | 0.80, 1.13 | 0.5 | 0.90 | 0.75, 1.08 | 0.3 |
| 200-399% FPL | 0.89 | 0.75, 1.05 | 0.2 | 0.96 | 0.81, 1.15 | 0.7 |
| 400% FPL or greater | 0.79 | 0.67, 0.95 | 0.011 | 1.03 | 0.85, 1.24 | 0.8 |
| Child's Sex Assigned at Birth |  |  |  |  |  |  |
| Male | — | — |  | — | — |  |
| Female | 1.88 | 1.71, 2.08 | <0.001 | 1.91 | 1.73, 2.12 | <0.001 |
| Child's Age in Years | 5.01 | 4.59, 5.46 | <0.001 | 4.90 | 4.47, 5.37 | <0.001 |
| Eviction Stress/Concern | 1.35 | 1.27, 1.44 | <0.001 | 1.10 | 1.02, 1.18 | 0.008 |
| Child's Age in Years * Eviction Stress/Concern | 0.93 | 0.88, 0.99 | 0.016 | 0.93 | 0.87, 0.98 | 0.012 |
| Places Lived, Last Year |  |  |  |  |  |  |
| 0-2 times |  |  |  | — | — |  |
| 3 or more times |  |  |  | 1.87 | 1.42, 2.46 | <0.001 |
| Ever Homeless |  |  |  |  |  |  |
| No |  |  |  | — | — |  |
| Yes |  |  |  | 1.42 | 1.12, 1.81 | 0.004 |
| Mother's Mental health |  |  |  |  |  |  |
| Excellent or very good |  |  |  | — | — |  |
| Good |  |  |  | 2.23 | 1.98, 2.50 | <0.001 |
| Fair or poor |  |  |  | 3.30 | 2.82, 3.86 | <0.001 |
| Low Birth Weight |  |  |  |  |  |  |
| No |  |  |  | — | — |  |
| Yes |  |  |  | 1.02 | 0.83, 1.24 | 0.9 |
| Born Premature |  |  |  |  |  |  |
| No |  |  |  | — | — |  |
| Yes |  |  |  | 1.09 | 0.92, 1.31 | 0.3 |
| Food Insecurity |  |  |  | 1.16 | 1.05, 1.27 | 0.003 |
| Adverse Childhood Experiences |  |  |  |  |  |  |
| 0 ACEs |  |  |  | — | — |  |
| 1 ACE |  |  |  | 1.83 | 1.58, 2.11 | <0.001 |
| 2+ ACEs |  |  |  | 4.85 | 4.22, 5.58 | <0.001 |
| *^1^*OR = Odds Ratio, CI = Confidence Interval | | | | | | |

\# Output as Word Doc
MDD_combined_table %>%
 gtsummary::as_gt() %>%
 gt::gtsave("Table2.docx")

**\# Analyses Related to Anxiety**

\# Anxiety Base Model
Anxiety_Base_Adjusted_Age_Interact<-glmer(anxiety_22_R_Alt ~ raceASIA_22_F_L + famstruct5_22_F_L + AdultEduc_22_F_L + povlev4_22_F_L + sex_22_R_L + Child_Age * HomeEvic_22_R_Z + (1|FIPSST), family=binomial, data=NSCH_Topical_DRC_CAHMI_subset, control = glmerControl(optimizer = "bobyqa",optCtrl=list(maxfun=2e5)),nAGQ = 0)
summary(Anxiety_Base_Adjusted_Age_Interact)

```
## Generalized linear mixed model fit by maximum likelihood (Adaptive
##   Gauss-Hermite Quadrature, nAGQ = 0) [glmerMod]
##  Family: binomial  ( logit )
## Formula:
## anxiety_22_R_Alt ~ raceASIA_22_F_L + famstruct5_22_F_L + AdultEduc_22_F_L +
##     povlev4_22_F_L + sex_22_R_L + Child_Age * HomeEvic_22_R_Z +
##     (1 | FIPSST)
##    Data: NSCH_Topical_DRC_CAHMI_subset
## Control: glmerControl(optimizer = "bobyqa", optCtrl = list(maxfun = 2e+05))
##
##      AIC      BIC   logLik deviance df.resid
##  25148.5  25318.7 -12554.3  25108.5    36618
##
## Scaled residuals:
##     Min      1Q  Median      3Q     Max
## -1.5226 -0.4247 -0.2769 -0.1910 14.1892
##
## Random effects:
##  Groups Name        Variance Std.Dev.
##  FIPSST (Intercept) 0.03075  0.1754
## Number of obs: 36638, groups:  FIPSST, 51
##
## Fixed effects:
##                                                     Estimate Std. Error z value
## (Intercept)                                         -3.17989    0.14593 -21.791
## raceASIA_22_F_LHispanic                             -0.31881    0.05232  -6.093
## raceASIA_22_F_LBlack, non-Hispanic                  -1.11181    0.09466 -11.745
## raceASIA_22_F_LAsian, non-Hispanic                  -1.36339    0.11220 -12.152
## raceASIA_22_F_LMulti-racial, non-Hispanic           -0.17497    0.06280  -2.786
## famstruct5_22_F_LTwo parents, not currently married  0.31514    0.07144   4.411
## famstruct5_22_F_LSingle parent (mother or father)    0.43588    0.04384   9.943
## famstruct5_22_F_LGrandparent household               0.88254    1.18356   0.746
## famstruct5_22_F_LOther family type                   1.06919    0.35134   3.043
## AdultEduc_22_F_LHigh school or GED                   0.45999    0.14214   3.236
## AdultEduc_22_F_LSome college or technical school     0.63827    0.13935   4.580
## AdultEduc_22_F_LCollege degree or higher             0.70678    0.13925   5.075
## povlev4_22_F_L100-199% FPL                           0.06965    0.06426   1.084
## povlev4_22_F_L200-399% FPL                           0.02900    0.06105   0.475
## povlev4_22_F_L400% FPL or greater                   -0.03812    0.06309  -0.604
## sex_22_R_LFemale                                     0.41929    0.03292  12.737
## Child_Age                                            0.89121    0.02135  41.738
## HomeEvic_22_R_Z                                      0.23236    0.01816  12.798
## Child_Age:HomeEvic_22_R_Z                           -0.03596    0.01771  -2.030
##                                                     Pr(>|z|)
## (Intercept)                                          < 2e-16 ***
## raceASIA_22_F_LHispanic                             1.11e-09 ***
## raceASIA_22_F_LBlack, non-Hispanic                   < 2e-16 ***
## raceASIA_22_F_LAsian, non-Hispanic                   < 2e-16 ***
## raceASIA_22_F_LMulti-racial, non-Hispanic            0.00534 **
## famstruct5_22_F_LTwo parents, not currently married 1.03e-05 ***
## famstruct5_22_F_LSingle parent (mother or father)    < 2e-16 ***
## famstruct5_22_F_LGrandparent household               0.45587
## famstruct5_22_F_LOther family type                   0.00234 **
## AdultEduc_22_F_LHigh school or GED                   0.00121 **
## AdultEduc_22_F_LSome college or technical school    4.64e-06 ***
## AdultEduc_22_F_LCollege degree or higher            3.87e-07 ***
## povlev4_22_F_L100-199% FPL                           0.27839
## povlev4_22_F_L200-399% FPL                           0.63477
## povlev4_22_F_L400% FPL or greater                    0.54575
## sex_22_R_LFemale                                     < 2e-16 ***
## Child_Age                                            < 2e-16 ***
## HomeEvic_22_R_Z                                      < 2e-16 ***
## Child_Age:HomeEvic_22_R_Z                            0.04237 *
## ---
## Signif. codes:  0 '***' 0.001 '**' 0.01 '*' 0.05 '.' 0.1 ' ' 1
##
## Correlation matrix not shown by default, as p = 19 > 12.
## Use print(x, correlation=TRUE)  or
##     vcov(x)        if you need it
```

\#
modelbased::estimate_slopes(Anxiety_Base_Adjusted_Age_Interact, trend = "HomeEvic_22_R_Z", by ="Child_Age",nuisance=c("raceASIA_22_F_L","famstruct5_22_F_L","AdultEduc_22_F_L","povlev4_22_F_L","sex_22_R_L"),length = 100)

```
## Estimated Marginal Effects
##
## Child_Age | Coefficient |   SE |       95% CI |     z |  df |      p
## --------------------------------------------------------------------
## -1.06     |        0.27 | 0.03 | [0.21, 0.33] |  8.30 | Inf | < .001
## -1.03     |        0.27 | 0.03 | [0.21, 0.33] |  8.38 | Inf | < .001
## -1.00     |        0.27 | 0.03 | [0.21, 0.33] |  8.46 | Inf | < .001
## -0.98     |        0.27 | 0.03 | [0.21, 0.33] |  8.54 | Inf | < .001
## -0.95     |        0.27 | 0.03 | [0.21, 0.33] |  8.63 | Inf | < .001
## -0.92     |        0.27 | 0.03 | [0.21, 0.33] |  8.71 | Inf | < .001
## -0.90     |        0.26 | 0.03 | [0.21, 0.32] |  8.80 | Inf | < .001
## -0.87     |        0.26 | 0.03 | [0.21, 0.32] |  8.89 | Inf | < .001
## -0.84     |        0.26 | 0.03 | [0.21, 0.32] |  8.98 | Inf | < .001
## -0.82     |        0.26 | 0.03 | [0.21, 0.32] |  9.07 | Inf | < .001
## -0.79     |        0.26 | 0.03 | [0.20, 0.32] |  9.16 | Inf | < .001
## -0.76     |        0.26 | 0.03 | [0.20, 0.31] |  9.26 | Inf | < .001
## -0.74     |        0.26 | 0.03 | [0.20, 0.31] |  9.36 | Inf | < .001
## -0.71     |        0.26 | 0.03 | [0.20, 0.31] |  9.46 | Inf | < .001
## -0.68     |        0.26 | 0.03 | [0.20, 0.31] |  9.56 | Inf | < .001
## -0.66     |        0.26 | 0.03 | [0.20, 0.31] |  9.67 | Inf | < .001
## -0.63     |        0.26 | 0.03 | [0.20, 0.31] |  9.77 | Inf | < .001
## -0.60     |        0.25 | 0.03 | [0.20, 0.30] |  9.88 | Inf | < .001
## -0.58     |        0.25 | 0.03 | [0.20, 0.30] |  9.99 | Inf | < .001
## -0.55     |        0.25 | 0.02 | [0.20, 0.30] | 10.10 | Inf | < .001
## -0.52     |        0.25 | 0.02 | [0.20, 0.30] | 10.22 | Inf | < .001
## -0.50     |        0.25 | 0.02 | [0.20, 0.30] | 10.34 | Inf | < .001
## -0.47     |        0.25 | 0.02 | [0.20, 0.30] | 10.46 | Inf | < .001
## -0.44     |        0.25 | 0.02 | [0.20, 0.29] | 10.58 | Inf | < .001
## -0.42     |        0.25 | 0.02 | [0.20, 0.29] | 10.70 | Inf | < .001
## -0.39     |        0.25 | 0.02 | [0.20, 0.29] | 10.83 | Inf | < .001
## -0.36     |        0.25 | 0.02 | [0.20, 0.29] | 10.95 | Inf | < .001
## -0.34     |        0.24 | 0.02 | [0.20, 0.29] | 11.08 | Inf | < .001
## -0.31     |        0.24 | 0.02 | [0.20, 0.29] | 11.21 | Inf | < .001
## -0.28     |        0.24 | 0.02 | [0.20, 0.28] | 11.35 | Inf | < .001
## -0.26     |        0.24 | 0.02 | [0.20, 0.28] | 11.48 | Inf | < .001
## -0.23     |        0.24 | 0.02 | [0.20, 0.28] | 11.62 | Inf | < .001
## -0.20     |        0.24 | 0.02 | [0.20, 0.28] | 11.75 | Inf | < .001
## -0.18     |        0.24 | 0.02 | [0.20, 0.28] | 11.89 | Inf | < .001
## -0.15     |        0.24 | 0.02 | [0.20, 0.28] | 12.03 | Inf | < .001
## -0.12     |        0.24 | 0.02 | [0.20, 0.27] | 12.16 | Inf | < .001
## -0.10     |        0.24 | 0.02 | [0.20, 0.27] | 12.30 | Inf | < .001
## -0.07     |        0.23 | 0.02 | [0.20, 0.27] | 12.44 | Inf | < .001
## -0.04     |        0.23 | 0.02 | [0.20, 0.27] | 12.58 | Inf | < .001
## -0.02     |        0.23 | 0.02 | [0.20, 0.27] | 12.71 | Inf | < .001
## 9.73e-03  |        0.23 | 0.02 | [0.20, 0.27] | 12.85 | Inf | < .001
## 0.04      |        0.23 | 0.02 | [0.20, 0.27] | 12.98 | Inf | < .001
## 0.06      |        0.23 | 0.02 | [0.20, 0.26] | 13.11 | Inf | < .001
## 0.09      |        0.23 | 0.02 | [0.20, 0.26] | 13.23 | Inf | < .001
## 0.12      |        0.23 | 0.02 | [0.19, 0.26] | 13.35 | Inf | < .001
## 0.14      |        0.23 | 0.02 | [0.19, 0.26] | 13.47 | Inf | < .001
## 0.17      |        0.23 | 0.02 | [0.19, 0.26] | 13.58 | Inf | < .001
## 0.20      |        0.23 | 0.02 | [0.19, 0.26] | 13.69 | Inf | < .001
## 0.22      |        0.22 | 0.02 | [0.19, 0.26] | 13.78 | Inf | < .001
## 0.25      |        0.22 | 0.02 | [0.19, 0.25] | 13.87 | Inf | < .001
## 0.28      |        0.22 | 0.02 | [0.19, 0.25] | 13.96 | Inf | < .001
## 0.30      |        0.22 | 0.02 | [0.19, 0.25] | 14.03 | Inf | < .001
## 0.33      |        0.22 | 0.02 | [0.19, 0.25] | 14.09 | Inf | < .001
## 0.36      |        0.22 | 0.02 | [0.19, 0.25] | 14.14 | Inf | < .001
## 0.38      |        0.22 | 0.02 | [0.19, 0.25] | 14.18 | Inf | < .001
## 0.41      |        0.22 | 0.02 | [0.19, 0.25] | 14.21 | Inf | < .001
## 0.44      |        0.22 | 0.02 | [0.19, 0.25] | 14.22 | Inf | < .001
## 0.46      |        0.22 | 0.02 | [0.19, 0.25] | 14.22 | Inf | < .001
## 0.49      |        0.21 | 0.02 | [0.19, 0.24] | 14.21 | Inf | < .001
## 0.52      |        0.21 | 0.02 | [0.18, 0.24] | 14.18 | Inf | < .001
## 0.54      |        0.21 | 0.02 | [0.18, 0.24] | 14.14 | Inf | < .001
## 0.57      |        0.21 | 0.02 | [0.18, 0.24] | 14.09 | Inf | < .001
## 0.60      |        0.21 | 0.02 | [0.18, 0.24] | 14.02 | Inf | < .001
## 0.62      |        0.21 | 0.02 | [0.18, 0.24] | 13.94 | Inf | < .001
## 0.65      |        0.21 | 0.02 | [0.18, 0.24] | 13.85 | Inf | < .001
## 0.68      |        0.21 | 0.02 | [0.18, 0.24] | 13.74 | Inf | < .001
## 0.70      |        0.21 | 0.02 | [0.18, 0.24] | 13.62 | Inf | < .001
## 0.73      |        0.21 | 0.02 | [0.18, 0.24] | 13.49 | Inf | < .001
## 0.76      |        0.21 | 0.02 | [0.18, 0.24] | 13.34 | Inf | < .001
## 0.78      |        0.20 | 0.02 | [0.17, 0.23] | 13.19 | Inf | < .001
## 0.81      |        0.20 | 0.02 | [0.17, 0.23] | 13.03 | Inf | < .001
## 0.84      |        0.20 | 0.02 | [0.17, 0.23] | 12.86 | Inf | < .001
## 0.86      |        0.20 | 0.02 | [0.17, 0.23] | 12.68 | Inf | < .001
## 0.89      |        0.20 | 0.02 | [0.17, 0.23] | 12.50 | Inf | < .001
## 0.92      |        0.20 | 0.02 | [0.17, 0.23] | 12.30 | Inf | < .001
## 0.94      |        0.20 | 0.02 | [0.17, 0.23] | 12.11 | Inf | < .001
## 0.97      |        0.20 | 0.02 | [0.17, 0.23] | 11.91 | Inf | < .001
## 1.00      |        0.20 | 0.02 | [0.16, 0.23] | 11.71 | Inf | < .001
## 1.02      |        0.20 | 0.02 | [0.16, 0.23] | 11.50 | Inf | < .001
## 1.05      |        0.19 | 0.02 | [0.16, 0.23] | 11.30 | Inf | < .001
## 1.08      |        0.19 | 0.02 | [0.16, 0.23] | 11.09 | Inf | < .001
## 1.10      |        0.19 | 0.02 | [0.16, 0.23] | 10.88 | Inf | < .001
## 1.13      |        0.19 | 0.02 | [0.16, 0.23] | 10.68 | Inf | < .001
## 1.16      |        0.19 | 0.02 | [0.16, 0.23] | 10.47 | Inf | < .001
## 1.18      |        0.19 | 0.02 | [0.15, 0.23] | 10.27 | Inf | < .001
## 1.21      |        0.19 | 0.02 | [0.15, 0.23] | 10.06 | Inf | < .001
## 1.24      |        0.19 | 0.02 | [0.15, 0.23] |  9.86 | Inf | < .001
## 1.26      |        0.19 | 0.02 | [0.15, 0.22] |  9.66 | Inf | < .001
## 1.29      |        0.19 | 0.02 | [0.15, 0.22] |  9.47 | Inf | < .001
## 1.32      |        0.19 | 0.02 | [0.15, 0.22] |  9.27 | Inf | < .001
## 1.34      |        0.18 | 0.02 | [0.14, 0.22] |  9.08 | Inf | < .001
## 1.37      |        0.18 | 0.02 | [0.14, 0.22] |  8.89 | Inf | < .001
## 1.40      |        0.18 | 0.02 | [0.14, 0.22] |  8.71 | Inf | < .001
## 1.42      |        0.18 | 0.02 | [0.14, 0.22] |  8.53 | Inf | < .001
## 1.45      |        0.18 | 0.02 | [0.14, 0.22] |  8.35 | Inf | < .001
## 1.48      |        0.18 | 0.02 | [0.14, 0.22] |  8.18 | Inf | < .001
## 1.50      |        0.18 | 0.02 | [0.13, 0.22] |  8.01 | Inf | < .001
## 1.53      |        0.18 | 0.02 | [0.13, 0.22] |  7.84 | Inf | < .001
## 1.56      |        0.18 | 0.02 | [0.13, 0.22] |  7.68 | Inf | < .001
## 1.58      |        0.18 | 0.02 | [0.13, 0.22] |  7.52 | Inf | < .001
## Marginal effects estimated for HomeEvic_22_R_Z
```

\# Anxiety Strict Model
Anxiety_Stringent_Adjusted_Age_Interact<- glmer(anxiety_22_R_Alt ~ raceASIA_22_F_L + famstruct5_22_F_L + AdultEduc_22_F_L + povlev4_22_F_L + sex_22_R_L + PlacesLived_22_R_F_L + MotherMH_22_R_F_L + EverHomeless_22_R_F_L + LowBWght_22_R_F_L + BornPre_22_R_F_L + FoodSit_22_R + ACE2more11_22_R_F_L + Child_Age * HomeEvic_22_R_Z + (1|FIPSST), family=binomial, data=NSCH_Topical_DRC_CAHMI_subset, control = glmerControl(optimizer = "bobyqa",optCtrl=list(maxfun=2e5)),nAGQ = 0)
summary(Anxiety_Stringent_Adjusted_Age_Interact)

```
## Generalized linear mixed model fit by maximum likelihood (Adaptive
##   Gauss-Hermite Quadrature, nAGQ = 0) [glmerMod]
##  Family: binomial  ( logit )
## Formula:
## anxiety_22_R_Alt ~ raceASIA_22_F_L + famstruct5_22_F_L + AdultEduc_22_F_L +
##     povlev4_22_F_L + sex_22_R_L + PlacesLived_22_R_F_L + MotherMH_22_R_F_L +
##     EverHomeless_22_R_F_L + LowBWght_22_R_F_L + BornPre_22_R_F_L +
##     FoodSit_22_R + ACE2more11_22_R_F_L + Child_Age * HomeEvic_22_R_Z +
##     (1 | FIPSST)
##    Data: NSCH_Topical_DRC_CAHMI_subset
## Control: glmerControl(optimizer = "bobyqa", optCtrl = list(maxfun = 2e+05))
##
##      AIC      BIC   logLik deviance df.resid
##  23590.3  23837.1 -11766.2  23532.3    36609
##
## Scaled residuals:
##     Min      1Q  Median      3Q     Max
## -2.2170 -0.3888 -0.2484 -0.1589 14.9136
##
## Random effects:
##  Groups Name        Variance Std.Dev.
##  FIPSST (Intercept) 0.02981  0.1727
## Number of obs: 36638, groups:  FIPSST, 51
##
## Fixed effects:
##                                                     Estimate Std. Error z value
## (Intercept)                                         -4.09667    0.16202 -25.285
## raceASIA_22_F_LHispanic                             -0.33236    0.05374  -6.184
## raceASIA_22_F_LBlack, non-Hispanic                  -1.05677    0.09671 -10.927
## raceASIA_22_F_LAsian, non-Hispanic                  -1.18525    0.11407 -10.390
## raceASIA_22_F_LMulti-racial, non-Hispanic           -0.31780    0.06507  -4.884
## famstruct5_22_F_LTwo parents, not currently married -0.11859    0.07560  -1.569
## famstruct5_22_F_LSingle parent (mother or father)   -0.14976    0.04886  -3.065
## famstruct5_22_F_LGrandparent household               0.63577    1.22064   0.521
## famstruct5_22_F_LOther family type                   0.31060    0.36629   0.848
## AdultEduc_22_F_LHigh school or GED                   0.31099    0.14782   2.104
## AdultEduc_22_F_LSome college or technical school     0.46688    0.14509   3.218
## AdultEduc_22_F_LCollege degree or higher             0.69627    0.14513   4.798
## povlev4_22_F_L100-199% FPL                           0.03722    0.06693   0.556
## povlev4_22_F_L200-399% FPL                           0.08907    0.06365   1.399
## povlev4_22_F_L400% FPL or greater                    0.17144    0.06654   2.576
## sex_22_R_LFemale                                     0.43777    0.03404  12.862
## PlacesLived_22_R_F_L3 or more times                  0.25838    0.10905   2.369
## MotherMH_22_R_F_LGood                                0.63218    0.03867  16.350
## MotherMH_22_R_F_LFair or poor                        1.02535    0.05677  18.062
## EverHomeless_22_R_F_LYes                             0.25553    0.10070   2.537
## LowBWght_22_R_F_LYes                                -0.00348    0.06768  -0.051
## BornPre_22_R_F_LYes                                  0.22924    0.05942   3.858
## FoodSit_22_R                                         0.19956    0.03466   5.758
## ACE2more11_22_R_F_L1 ACE                             0.57415    0.04532  12.668
## ACE2more11_22_R_F_L2+ ACEs                           1.21160    0.04899  24.729
## Child_Age                                            0.84900    0.02253  37.675
## HomeEvic_22_R_Z                                      0.03828    0.02037   1.879
## Child_Age:HomeEvic_22_R_Z                           -0.03531    0.01878  -1.881
##                                                     Pr(>|z|)
## (Intercept)                                          < 2e-16 ***
## raceASIA_22_F_LHispanic                             6.25e-10 ***
## raceASIA_22_F_LBlack, non-Hispanic                   < 2e-16 ***
## raceASIA_22_F_LAsian, non-Hispanic                   < 2e-16 ***
## raceASIA_22_F_LMulti-racial, non-Hispanic           1.04e-06 ***
## famstruct5_22_F_LTwo parents, not currently married 0.116724
## famstruct5_22_F_LSingle parent (mother or father)   0.002176 **
## famstruct5_22_F_LGrandparent household              0.602473
## famstruct5_22_F_LOther family type                  0.396459
## AdultEduc_22_F_LHigh school or GED                  0.035394 *
## AdultEduc_22_F_LSome college or technical school    0.001291 **
## AdultEduc_22_F_LCollege degree or higher            1.60e-06 ***
## povlev4_22_F_L100-199% FPL                          0.578164
## povlev4_22_F_L200-399% FPL                          0.161717
## povlev4_22_F_L400% FPL or greater                   0.009982 **
## sex_22_R_LFemale                                     < 2e-16 ***
## PlacesLived_22_R_F_L3 or more times                 0.017823 *
## MotherMH_22_R_F_LGood                                < 2e-16 ***
## MotherMH_22_R_F_LFair or poor                        < 2e-16 ***
## EverHomeless_22_R_F_LYes                            0.011167 *
## LowBWght_22_R_F_LYes                                0.958994
## BornPre_22_R_F_LYes                                 0.000115 ***
## FoodSit_22_R                                        8.53e-09 ***
## ACE2more11_22_R_F_L1 ACE                             < 2e-16 ***
## ACE2more11_22_R_F_L2+ ACEs                           < 2e-16 ***
## Child_Age                                            < 2e-16 ***
## HomeEvic_22_R_Z                                     0.060222 .
## Child_Age:HomeEvic_22_R_Z                           0.060010 .
## ---
## Signif. codes:  0 '***' 0.001 '**' 0.01 '*' 0.05 '.' 0.1 ' ' 1
##
## Correlation matrix not shown by default, as p = 28 > 12.
## Use print(x, correlation=TRUE)  or
##     vcov(x)        if you need it
```

\#
modelbased::estimate_slopes(Anxiety_Stringent_Adjusted_Age_Interact, trend = "HomeEvic_22_R_Z", by = "Child_Age",nuisance=c("raceASIA_22_F_L","famstruct5_22_F_L","AdultEduc_22_F_L","povlev4_22_F_L","sex_22_R_L","PlacesLived_22_R_F_L","MotherMH_22_R_F_L","EverHomeless_22_R_F_L","LowBWght_22_R_F_L","BornPre_22_R_F_L","FoodSit_22_R","ACE2more11_22_R_F_L"), length = 100)

```
## Estimated Marginal Effects
##
## Child_Age | Coefficient |   SE |        95% CI |     z |  df |     p
## --------------------------------------------------------------------
## -1.06     |        0.08 | 0.04 | [ 0.01, 0.14] |  2.15 | Inf | 0.032
## -1.03     |        0.07 | 0.03 | [ 0.01, 0.14] |  2.15 | Inf | 0.032
## -1.00     |        0.07 | 0.03 | [ 0.01, 0.14] |  2.15 | Inf | 0.032
## -0.98     |        0.07 | 0.03 | [ 0.01, 0.14] |  2.15 | Inf | 0.032
## -0.95     |        0.07 | 0.03 | [ 0.01, 0.14] |  2.15 | Inf | 0.032
## -0.92     |        0.07 | 0.03 | [ 0.01, 0.14] |  2.15 | Inf | 0.032
## -0.90     |        0.07 | 0.03 | [ 0.01, 0.13] |  2.15 | Inf | 0.032
## -0.87     |        0.07 | 0.03 | [ 0.01, 0.13] |  2.15 | Inf | 0.032
## -0.84     |        0.07 | 0.03 | [ 0.01, 0.13] |  2.15 | Inf | 0.032
## -0.82     |        0.07 | 0.03 | [ 0.01, 0.13] |  2.14 | Inf | 0.032
## -0.79     |        0.07 | 0.03 | [ 0.01, 0.13] |  2.14 | Inf | 0.032
## -0.76     |        0.07 | 0.03 | [ 0.01, 0.12] |  2.14 | Inf | 0.032
## -0.74     |        0.06 | 0.03 | [ 0.01, 0.12] |  2.14 | Inf | 0.032
## -0.71     |        0.06 | 0.03 | [ 0.01, 0.12] |  2.14 | Inf | 0.033
## -0.68     |        0.06 | 0.03 | [ 0.01, 0.12] |  2.13 | Inf | 0.033
## -0.66     |        0.06 | 0.03 | [ 0.00, 0.12] |  2.13 | Inf | 0.033
## -0.63     |        0.06 | 0.03 | [ 0.00, 0.12] |  2.13 | Inf | 0.033
## -0.60     |        0.06 | 0.03 | [ 0.00, 0.11] |  2.12 | Inf | 0.034
## -0.58     |        0.06 | 0.03 | [ 0.00, 0.11] |  2.12 | Inf | 0.034
## -0.55     |        0.06 | 0.03 | [ 0.00, 0.11] |  2.12 | Inf | 0.034
## -0.52     |        0.06 | 0.03 | [ 0.00, 0.11] |  2.11 | Inf | 0.035
## -0.50     |        0.06 | 0.03 | [ 0.00, 0.11] |  2.11 | Inf | 0.035
## -0.47     |        0.05 | 0.03 | [ 0.00, 0.11] |  2.10 | Inf | 0.036
## -0.44     |        0.05 | 0.03 | [ 0.00, 0.10] |  2.09 | Inf | 0.036
## -0.42     |        0.05 | 0.03 | [ 0.00, 0.10] |  2.09 | Inf | 0.037
## -0.39     |        0.05 | 0.03 | [ 0.00, 0.10] |  2.08 | Inf | 0.038
## -0.36     |        0.05 | 0.02 | [ 0.00, 0.10] |  2.07 | Inf | 0.038
## -0.34     |        0.05 | 0.02 | [ 0.00, 0.10] |  2.06 | Inf | 0.039
## -0.31     |        0.05 | 0.02 | [ 0.00, 0.10] |  2.05 | Inf | 0.040
## -0.28     |        0.05 | 0.02 | [ 0.00, 0.09] |  2.04 | Inf | 0.041
## -0.26     |        0.05 | 0.02 | [ 0.00, 0.09] |  2.03 | Inf | 0.042
## -0.23     |        0.05 | 0.02 | [ 0.00, 0.09] |  2.02 | Inf | 0.043
## -0.20     |        0.05 | 0.02 | [ 0.00, 0.09] |  2.01 | Inf | 0.044
## -0.18     |        0.04 | 0.02 | [ 0.00, 0.09] |  2.00 | Inf | 0.046
## -0.15     |        0.04 | 0.02 | [ 0.00, 0.09] |  1.98 | Inf | 0.048
## -0.12     |        0.04 | 0.02 | [ 0.00, 0.09] |  1.97 | Inf | 0.049
## -0.10     |        0.04 | 0.02 | [ 0.00, 0.08] |  1.95 | Inf | 0.051
## -0.07     |        0.04 | 0.02 | [ 0.00, 0.08] |  1.93 | Inf | 0.053
## -0.04     |        0.04 | 0.02 | [ 0.00, 0.08] |  1.91 | Inf | 0.056
## -0.02     |        0.04 | 0.02 | [ 0.00, 0.08] |  1.89 | Inf | 0.058
## 9.73e-03  |        0.04 | 0.02 | [ 0.00, 0.08] |  1.87 | Inf | 0.061
## 0.04      |        0.04 | 0.02 | [ 0.00, 0.08] |  1.85 | Inf | 0.065
## 0.06      |        0.04 | 0.02 | [ 0.00, 0.07] |  1.82 | Inf | 0.068
## 0.09      |        0.04 | 0.02 | [ 0.00, 0.07] |  1.80 | Inf | 0.072
## 0.12      |        0.03 | 0.02 | [ 0.00, 0.07] |  1.77 | Inf | 0.077
## 0.14      |        0.03 | 0.02 | [ 0.00, 0.07] |  1.74 | Inf | 0.082
## 0.17      |        0.03 | 0.02 | [ 0.00, 0.07] |  1.71 | Inf | 0.087
## 0.20      |        0.03 | 0.02 | [-0.01, 0.07] |  1.68 | Inf | 0.093
## 0.22      |        0.03 | 0.02 | [-0.01, 0.07] |  1.64 | Inf | 0.100
## 0.25      |        0.03 | 0.02 | [-0.01, 0.07] |  1.61 | Inf | 0.108
## 0.28      |        0.03 | 0.02 | [-0.01, 0.06] |  1.57 | Inf | 0.116
## 0.30      |        0.03 | 0.02 | [-0.01, 0.06] |  1.53 | Inf | 0.126
## 0.33      |        0.03 | 0.02 | [-0.01, 0.06] |  1.49 | Inf | 0.136
## 0.36      |        0.03 | 0.02 | [-0.01, 0.06] |  1.45 | Inf | 0.148
## 0.38      |        0.02 | 0.02 | [-0.01, 0.06] |  1.40 | Inf | 0.161
## 0.41      |        0.02 | 0.02 | [-0.01, 0.06] |  1.36 | Inf | 0.175
## 0.44      |        0.02 | 0.02 | [-0.01, 0.06] |  1.31 | Inf | 0.191
## 0.46      |        0.02 | 0.02 | [-0.01, 0.06] |  1.26 | Inf | 0.208
## 0.49      |        0.02 | 0.02 | [-0.01, 0.06] |  1.21 | Inf | 0.227
## 0.52      |        0.02 | 0.02 | [-0.01, 0.05] |  1.16 | Inf | 0.247
## 0.54      |        0.02 | 0.02 | [-0.01, 0.05] |  1.10 | Inf | 0.269
## 0.57      |        0.02 | 0.02 | [-0.02, 0.05] |  1.05 | Inf | 0.293
## 0.60      |        0.02 | 0.02 | [-0.02, 0.05] |  1.00 | Inf | 0.319
## 0.62      |        0.02 | 0.02 | [-0.02, 0.05] |  0.94 | Inf | 0.347
## 0.65      |        0.02 | 0.02 | [-0.02, 0.05] |  0.88 | Inf | 0.376
## 0.68      |        0.01 | 0.02 | [-0.02, 0.05] |  0.83 | Inf | 0.407
## 0.70      |        0.01 | 0.02 | [-0.02, 0.05] |  0.77 | Inf | 0.440
## 0.73      |        0.01 | 0.02 | [-0.02, 0.05] |  0.71 | Inf | 0.475
## 0.76      |        0.01 | 0.02 | [-0.02, 0.05] |  0.66 | Inf | 0.511
## 0.78      |        0.01 | 0.02 | [-0.02, 0.05] |  0.60 | Inf | 0.548
## 0.81      |    9.71e-03 | 0.02 | [-0.03, 0.04] |  0.54 | Inf | 0.586
## 0.84      |    8.77e-03 | 0.02 | [-0.03, 0.04] |  0.49 | Inf | 0.626
## 0.86      |    7.83e-03 | 0.02 | [-0.03, 0.04] |  0.43 | Inf | 0.666
## 0.89      |    6.89e-03 | 0.02 | [-0.03, 0.04] |  0.38 | Inf | 0.706
## 0.92      |    5.95e-03 | 0.02 | [-0.03, 0.04] |  0.32 | Inf | 0.747
## 0.94      |    5.01e-03 | 0.02 | [-0.03, 0.04] |  0.27 | Inf | 0.788
## 0.97      |    4.06e-03 | 0.02 | [-0.03, 0.04] |  0.22 | Inf | 0.829
## 1.00      |    3.12e-03 | 0.02 | [-0.03, 0.04] |  0.16 | Inf | 0.870
## 1.02      |    2.18e-03 | 0.02 | [-0.04, 0.04] |  0.11 | Inf | 0.910
## 1.05      |    1.24e-03 | 0.02 | [-0.04, 0.04] |  0.06 | Inf | 0.949
## 1.08      |    3.01e-04 | 0.02 | [-0.04, 0.04] |  0.02 | Inf | 0.988
## 1.10      |   -6.40e-04 | 0.02 | [-0.04, 0.04] | -0.03 | Inf | 0.974
## 1.13      |   -1.58e-03 | 0.02 | [-0.04, 0.04] | -0.08 | Inf | 0.938
## 1.16      |   -2.52e-03 | 0.02 | [-0.04, 0.04] | -0.12 | Inf | 0.902
## 1.18      |   -3.46e-03 | 0.02 | [-0.04, 0.04] | -0.17 | Inf | 0.867
## 1.21      |   -4.40e-03 | 0.02 | [-0.05, 0.04] | -0.21 | Inf | 0.834
## 1.24      |   -5.34e-03 | 0.02 | [-0.05, 0.04] | -0.25 | Inf | 0.802
## 1.26      |   -6.29e-03 | 0.02 | [-0.05, 0.04] | -0.29 | Inf | 0.771
## 1.29      |   -7.23e-03 | 0.02 | [-0.05, 0.04] | -0.33 | Inf | 0.741
## 1.32      |   -8.17e-03 | 0.02 | [-0.05, 0.04] | -0.37 | Inf | 0.713
## 1.34      |   -9.11e-03 | 0.02 | [-0.05, 0.04] | -0.40 | Inf | 0.686
## 1.37      |       -0.01 | 0.02 | [-0.05, 0.03] | -0.44 | Inf | 0.660
## 1.40      |       -0.01 | 0.02 | [-0.06, 0.03] | -0.47 | Inf | 0.635
## 1.42      |       -0.01 | 0.02 | [-0.06, 0.03] | -0.51 | Inf | 0.612
## 1.45      |       -0.01 | 0.02 | [-0.06, 0.03] | -0.54 | Inf | 0.589
## 1.48      |       -0.01 | 0.02 | [-0.06, 0.03] | -0.57 | Inf | 0.568
## 1.50      |       -0.01 | 0.02 | [-0.06, 0.03] | -0.60 | Inf | 0.548
## 1.53      |       -0.02 | 0.02 | [-0.06, 0.03] | -0.63 | Inf | 0.529
## 1.56      |       -0.02 | 0.03 | [-0.07, 0.03] | -0.66 | Inf | 0.510
## 1.58      |       -0.02 | 0.03 | [-0.07, 0.03] | -0.69 | Inf | 0.493
## Marginal effects estimated for HomeEvic_22_R_Z
```

**\# Table Making Related to Anxiety (Table 03)**

\# Table with Anxiety Base Model
Anxiety_base_table <- tbl_regression(Anxiety_Base_Adjusted_Age_Interact, exponentiate = TRUE) %>% bold_p()

\# Table with Anxiety Strict Model
Anxiety_stringent_table <- tbl_regression(Anxiety_Stringent_Adjusted_Age_Interact, exponentiate = TRUE)%>% bold_p()

\# Combining Anxiety Tables
Anxiety_combined_table<-tbl_merge(
 tbls = list(Anxiety_base_table, Anxiety_stringent_table),
 tab_spanner = c("**Base Model**", "**Stringent Model**")
)

Anxiety_combined_table

|  | Base Model | | | Stringent Model | | |
| --- | --- | --- | --- | --- | --- | --- |
| Characteristic | OR*^1^* | 95% CI*^1^* | p-value | OR*^1^* | 95% CI*^1^* | p-value |
| Child's Race/Ethnicity |  |  |  |  |  |  |
| White, non-Hispanic | — | — |  | — | — |  |
| Hispanic | 0.73 | 0.66, 0.81 | <0.001 | 0.72 | 0.65, 0.80 | <0.001 |
| Black, non-Hispanic | 0.33 | 0.27, 0.40 | <0.001 | 0.35 | 0.29, 0.42 | <0.001 |
| Asian, non-Hispanic | 0.26 | 0.21, 0.32 | <0.001 | 0.31 | 0.24, 0.38 | <0.001 |
| Multi-racial, non-Hispanic | 0.84 | 0.74, 0.95 | 0.005 | 0.73 | 0.64, 0.83 | <0.001 |
| Family Structure |  |  |  |  |  |  |
| Two parents, currently married | — | — |  | — | — |  |
| Two parents, not currently married | 1.37 | 1.19, 1.58 | <0.001 | 0.89 | 0.77, 1.03 | 0.12 |
| Single parent (mother or father) | 1.55 | 1.42, 1.69 | <0.001 | 0.86 | 0.78, 0.95 | 0.002 |
| Grandparent household | 2.42 | 0.24, 24.6 | 0.5 | 1.89 | 0.17, 20.7 | 0.6 |
| Other family type | 2.91 | 1.46, 5.80 | 0.002 | 1.36 | 0.67, 2.80 | 0.4 |
| Highest Level of Education in Household |  |  |  |  |  |  |
| Less than high school | — | — |  | — | — |  |
| High school or GED | 1.58 | 1.20, 2.09 | 0.001 | 1.36 | 1.02, 1.82 | 0.035 |
| Some college or technical school | 1.89 | 1.44, 2.49 | <0.001 | 1.60 | 1.20, 2.12 | 0.001 |
| College degree or higher | 2.03 | 1.54, 2.66 | <0.001 | 2.01 | 1.51, 2.67 | <0.001 |
| Household Poverty Status |  |  |  |  |  |  |
| 0-99% FPL | — | — |  | — | — |  |
| 100-199% FPL | 1.07 | 0.95, 1.22 | 0.3 | 1.04 | 0.91, 1.18 | 0.6 |
| 200-399% FPL | 1.03 | 0.91, 1.16 | 0.6 | 1.09 | 0.96, 1.24 | 0.2 |
| 400% FPL or greater | 0.96 | 0.85, 1.09 | 0.5 | 1.19 | 1.04, 1.35 | 0.010 |
| Child's Sex Assigned at Birth |  |  |  |  |  |  |
| Male | — | — |  | — | — |  |
| Female | 1.52 | 1.43, 1.62 | <0.001 | 1.55 | 1.45, 1.66 | <0.001 |
| Child's Age in Years | 2.44 | 2.34, 2.54 | <0.001 | 2.34 | 2.24, 2.44 | <0.001 |
| Eviction Stress/Concern | 1.26 | 1.22, 1.31 | <0.001 | 1.04 | 1.00, 1.08 | 0.060 |
| Child's Age in Years * Eviction Stress/Concern | 0.96 | 0.93, 1.00 | 0.042 | 0.97 | 0.93, 1.00 | 0.060 |
| Places Lived, Last Year |  |  |  |  |  |  |
| 0-2 times |  |  |  | — | — |  |
| 3 or more times |  |  |  | 1.29 | 1.05, 1.60 | 0.018 |
| Mother's Mental health |  |  |  |  |  |  |
| Excellent or very good |  |  |  | — | — |  |
| Good |  |  |  | 1.88 | 1.74, 2.03 | <0.001 |
| Fair or poor |  |  |  | 2.79 | 2.49, 3.12 | <0.001 |
| Ever Homeless |  |  |  |  |  |  |
| No |  |  |  | — | — |  |
| Yes |  |  |  | 1.29 | 1.06, 1.57 | 0.011 |
| Low Birth Weight |  |  |  |  |  |  |
| No |  |  |  | — | — |  |
| Yes |  |  |  | 1.00 | 0.87, 1.14 | >0.9 |
| Born Premature |  |  |  |  |  |  |
| No |  |  |  | — | — |  |
| Yes |  |  |  | 1.26 | 1.12, 1.41 | <0.001 |
| Food Insecurity |  |  |  | 1.22 | 1.14, 1.31 | <0.001 |
| Adverse Childhood Experiences |  |  |  |  |  |  |
| 0 ACEs |  |  |  | — | — |  |
| 1 ACE |  |  |  | 1.78 | 1.62, 1.94 | <0.001 |
| 2+ ACEs |  |  |  | 3.36 | 3.05, 3.70 | <0.001 |
| *^1^*OR = Odds Ratio, CI = Confidence Interval | | | | | | |

\# Output as Word Doc
Anxiety_combined_table %>%
 gtsummary::as_gt() %>%
 gt::gtsave("Table3.docx")

**\# Analyses Related to ADHD**

\# ADHD Base Model
ADHD_Base_Adjusted_Age_Interact<-glmer(ADHD_22_R_Alt ~ raceASIA_22_F_L + famstruct5_22_F_L + AdultEduc_22_F_L + povlev4_22_F_L + sex_22_R_L + Child_Age * HomeEvic_22_R_Z + (1|FIPSST), family=binomial, data=NSCH_Topical_DRC_CAHMI_subset, control = glmerControl(optimizer = "bobyqa",optCtrl=list(maxfun=2e5)),nAGQ = 0)
summary(ADHD_Base_Adjusted_Age_Interact)

```
## Generalized linear mixed model fit by maximum likelihood (Adaptive
##   Gauss-Hermite Quadrature, nAGQ = 0) [glmerMod]
##  Family: binomial  ( logit )
## Formula:
## ADHD_22_R_Alt ~ raceASIA_22_F_L + famstruct5_22_F_L + AdultEduc_22_F_L +
##     povlev4_22_F_L + sex_22_R_L + Child_Age * HomeEvic_22_R_Z +
##     (1 | FIPSST)
##    Data: NSCH_Topical_DRC_CAHMI_subset
## Control: glmerControl(optimizer = "bobyqa", optCtrl = list(maxfun = 2e+05))
##
##      AIC      BIC   logLik deviance df.resid
##  23539.2  23709.4 -11749.6  23499.2    36618
##
## Scaled residuals:
##     Min      1Q  Median      3Q     Max
## -1.3318 -0.3858 -0.2862 -0.2049  9.2470
##
## Random effects:
##  Groups Name        Variance Std.Dev.
##  FIPSST (Intercept) 0.03258  0.1805
## Number of obs: 36638, groups:  FIPSST, 51
##
## Fixed effects:
##                                                      Estimate Std. Error
## (Intercept)                                          -2.24250    0.14197
## raceASIA_22_F_LHispanic                              -0.32980    0.05579
## raceASIA_22_F_LBlack, non-Hispanic                   -0.57060    0.08392
## raceASIA_22_F_LAsian, non-Hispanic                   -1.16837    0.11673
## raceASIA_22_F_LMulti-racial, non-Hispanic            -0.08686    0.06473
## famstruct5_22_F_LTwo parents, not currently married   0.28842    0.07334
## famstruct5_22_F_LSingle parent (mother or father)     0.35598    0.04614
## famstruct5_22_F_LGrandparent household              -10.84821  157.51642
## famstruct5_22_F_LOther family type                    1.62702    0.31933
## AdultEduc_22_F_LHigh school or GED                    0.36892    0.13918
## AdultEduc_22_F_LSome college or technical school      0.41705    0.13694
## AdultEduc_22_F_LCollege degree or higher              0.28857    0.13716
## povlev4_22_F_L100-199% FPL                           -0.11627    0.06505
## povlev4_22_F_L200-399% FPL                           -0.09135    0.06149
## povlev4_22_F_L400% FPL or greater                    -0.13406    0.06409
## sex_22_R_LFemale                                     -0.68235    0.03583
## Child_Age                                             0.62051    0.02132
## HomeEvic_22_R_Z                                       0.17391    0.01827
## Child_Age:HomeEvic_22_R_Z                            -0.03011    0.01803
##                                                     z value Pr(>|z|)
## (Intercept)                                         -15.796  < 2e-16 ***
## raceASIA_22_F_LHispanic                              -5.912 3.38e-09 ***
## raceASIA_22_F_LBlack, non-Hispanic                   -6.799 1.05e-11 ***
## raceASIA_22_F_LAsian, non-Hispanic                  -10.009  < 2e-16 ***
## raceASIA_22_F_LMulti-racial, non-Hispanic            -1.342  0.17967
## famstruct5_22_F_LTwo parents, not currently married   3.932 8.41e-05 ***
## famstruct5_22_F_LSingle parent (mother or father)     7.715 1.21e-14 ***
## famstruct5_22_F_LGrandparent household               -0.069  0.94509
## famstruct5_22_F_LOther family type                    5.095 3.49e-07 ***
## AdultEduc_22_F_LHigh school or GED                    2.651  0.00803 **
## AdultEduc_22_F_LSome college or technical school      3.046  0.00232 **
## AdultEduc_22_F_LCollege degree or higher              2.104  0.03538 *
## povlev4_22_F_L100-199% FPL                           -1.787  0.07389 .
## povlev4_22_F_L200-399% FPL                           -1.486  0.13738
## povlev4_22_F_L400% FPL or greater                    -2.092  0.03647 *
## sex_22_R_LFemale                                    -19.043  < 2e-16 ***
## Child_Age                                            29.106  < 2e-16 ***
## HomeEvic_22_R_Z                                       9.519  < 2e-16 ***
## Child_Age:HomeEvic_22_R_Z                            -1.670  0.09492 .
## ---
## Signif. codes:  0 '***' 0.001 '**' 0.01 '*' 0.05 '.' 0.1 ' ' 1
##
## Correlation matrix not shown by default, as p = 19 > 12.
## Use print(x, correlation=TRUE)  or
##     vcov(x)        if you need it
```

\#
modelbased::estimate_slopes(ADHD_Base_Adjusted_Age_Interact, trend = "HomeEvic_22_R_Z", by ="Child_Age",nuisance=c("raceASIA_22_F_L","famstruct5_22_F_L","AdultEduc_22_F_L","povlev4_22_F_L","sex_22_R_L"),length = 100)

```
## Estimated Marginal Effects
##
## Child_Age | Coefficient |   SE |       95% CI |     z |  df |      p
## --------------------------------------------------------------------
## -1.06     |        0.21 | 0.03 | [0.14, 0.27] |  6.34 | Inf | < .001
## -1.03     |        0.20 | 0.03 | [0.14, 0.27] |  6.40 | Inf | < .001
## -1.00     |        0.20 | 0.03 | [0.14, 0.27] |  6.46 | Inf | < .001
## -0.98     |        0.20 | 0.03 | [0.14, 0.26] |  6.52 | Inf | < .001
## -0.95     |        0.20 | 0.03 | [0.14, 0.26] |  6.58 | Inf | < .001
## -0.92     |        0.20 | 0.03 | [0.14, 0.26] |  6.64 | Inf | < .001
## -0.90     |        0.20 | 0.03 | [0.14, 0.26] |  6.70 | Inf | < .001
## -0.87     |        0.20 | 0.03 | [0.14, 0.26] |  6.77 | Inf | < .001
## -0.84     |        0.20 | 0.03 | [0.14, 0.26] |  6.84 | Inf | < .001
## -0.82     |        0.20 | 0.03 | [0.14, 0.25] |  6.90 | Inf | < .001
## -0.79     |        0.20 | 0.03 | [0.14, 0.25] |  6.97 | Inf | < .001
## -0.76     |        0.20 | 0.03 | [0.14, 0.25] |  7.05 | Inf | < .001
## -0.74     |        0.20 | 0.03 | [0.14, 0.25] |  7.12 | Inf | < .001
## -0.71     |        0.20 | 0.03 | [0.14, 0.25] |  7.19 | Inf | < .001
## -0.68     |        0.19 | 0.03 | [0.14, 0.25] |  7.27 | Inf | < .001
## -0.66     |        0.19 | 0.03 | [0.14, 0.25] |  7.34 | Inf | < .001
## -0.63     |        0.19 | 0.03 | [0.14, 0.24] |  7.42 | Inf | < .001
## -0.60     |        0.19 | 0.03 | [0.14, 0.24] |  7.50 | Inf | < .001
## -0.58     |        0.19 | 0.03 | [0.14, 0.24] |  7.58 | Inf | < .001
## -0.55     |        0.19 | 0.02 | [0.14, 0.24] |  7.66 | Inf | < .001
## -0.52     |        0.19 | 0.02 | [0.14, 0.24] |  7.75 | Inf | < .001
## -0.50     |        0.19 | 0.02 | [0.14, 0.24] |  7.83 | Inf | < .001
## -0.47     |        0.19 | 0.02 | [0.14, 0.23] |  7.92 | Inf | < .001
## -0.44     |        0.19 | 0.02 | [0.14, 0.23] |  8.00 | Inf | < .001
## -0.42     |        0.19 | 0.02 | [0.14, 0.23] |  8.09 | Inf | < .001
## -0.39     |        0.19 | 0.02 | [0.14, 0.23] |  8.18 | Inf | < .001
## -0.36     |        0.18 | 0.02 | [0.14, 0.23] |  8.27 | Inf | < .001
## -0.34     |        0.18 | 0.02 | [0.14, 0.23] |  8.36 | Inf | < .001
## -0.31     |        0.18 | 0.02 | [0.14, 0.23] |  8.45 | Inf | < .001
## -0.28     |        0.18 | 0.02 | [0.14, 0.22] |  8.55 | Inf | < .001
## -0.26     |        0.18 | 0.02 | [0.14, 0.22] |  8.64 | Inf | < .001
## -0.23     |        0.18 | 0.02 | [0.14, 0.22] |  8.73 | Inf | < .001
## -0.20     |        0.18 | 0.02 | [0.14, 0.22] |  8.83 | Inf | < .001
## -0.18     |        0.18 | 0.02 | [0.14, 0.22] |  8.92 | Inf | < .001
## -0.15     |        0.18 | 0.02 | [0.14, 0.22] |  9.01 | Inf | < .001
## -0.12     |        0.18 | 0.02 | [0.14, 0.22] |  9.11 | Inf | < .001
## -0.10     |        0.18 | 0.02 | [0.14, 0.21] |  9.20 | Inf | < .001
## -0.07     |        0.18 | 0.02 | [0.14, 0.21] |  9.29 | Inf | < .001
## -0.04     |        0.18 | 0.02 | [0.14, 0.21] |  9.38 | Inf | < .001
## -0.02     |        0.17 | 0.02 | [0.14, 0.21] |  9.46 | Inf | < .001
## 9.73e-03  |        0.17 | 0.02 | [0.14, 0.21] |  9.55 | Inf | < .001
## 0.04      |        0.17 | 0.02 | [0.14, 0.21] |  9.63 | Inf | < .001
## 0.06      |        0.17 | 0.02 | [0.14, 0.21] |  9.71 | Inf | < .001
## 0.09      |        0.17 | 0.02 | [0.14, 0.21] |  9.79 | Inf | < .001
## 0.12      |        0.17 | 0.02 | [0.14, 0.20] |  9.86 | Inf | < .001
## 0.14      |        0.17 | 0.02 | [0.14, 0.20] |  9.92 | Inf | < .001
## 0.17      |        0.17 | 0.02 | [0.14, 0.20] |  9.98 | Inf | < .001
## 0.20      |        0.17 | 0.02 | [0.14, 0.20] | 10.04 | Inf | < .001
## 0.22      |        0.17 | 0.02 | [0.13, 0.20] | 10.09 | Inf | < .001
## 0.25      |        0.17 | 0.02 | [0.13, 0.20] | 10.13 | Inf | < .001
## 0.28      |        0.17 | 0.02 | [0.13, 0.20] | 10.17 | Inf | < .001
## 0.30      |        0.16 | 0.02 | [0.13, 0.20] | 10.20 | Inf | < .001
## 0.33      |        0.16 | 0.02 | [0.13, 0.20] | 10.22 | Inf | < .001
## 0.36      |        0.16 | 0.02 | [0.13, 0.19] | 10.23 | Inf | < .001
## 0.38      |        0.16 | 0.02 | [0.13, 0.19] | 10.23 | Inf | < .001
## 0.41      |        0.16 | 0.02 | [0.13, 0.19] | 10.22 | Inf | < .001
## 0.44      |        0.16 | 0.02 | [0.13, 0.19] | 10.21 | Inf | < .001
## 0.46      |        0.16 | 0.02 | [0.13, 0.19] | 10.18 | Inf | < .001
## 0.49      |        0.16 | 0.02 | [0.13, 0.19] | 10.14 | Inf | < .001
## 0.52      |        0.16 | 0.02 | [0.13, 0.19] | 10.10 | Inf | < .001
## 0.54      |        0.16 | 0.02 | [0.13, 0.19] | 10.04 | Inf | < .001
## 0.57      |        0.16 | 0.02 | [0.13, 0.19] |  9.98 | Inf | < .001
## 0.60      |        0.16 | 0.02 | [0.13, 0.19] |  9.91 | Inf | < .001
## 0.62      |        0.16 | 0.02 | [0.12, 0.19] |  9.83 | Inf | < .001
## 0.65      |        0.15 | 0.02 | [0.12, 0.19] |  9.74 | Inf | < .001
## 0.68      |        0.15 | 0.02 | [0.12, 0.18] |  9.64 | Inf | < .001
## 0.70      |        0.15 | 0.02 | [0.12, 0.18] |  9.53 | Inf | < .001
## 0.73      |        0.15 | 0.02 | [0.12, 0.18] |  9.42 | Inf | < .001
## 0.76      |        0.15 | 0.02 | [0.12, 0.18] |  9.30 | Inf | < .001
## 0.78      |        0.15 | 0.02 | [0.12, 0.18] |  9.18 | Inf | < .001
## 0.81      |        0.15 | 0.02 | [0.12, 0.18] |  9.05 | Inf | < .001
## 0.84      |        0.15 | 0.02 | [0.12, 0.18] |  8.92 | Inf | < .001
## 0.86      |        0.15 | 0.02 | [0.11, 0.18] |  8.78 | Inf | < .001
## 0.89      |        0.15 | 0.02 | [0.11, 0.18] |  8.64 | Inf | < .001
## 0.92      |        0.15 | 0.02 | [0.11, 0.18] |  8.49 | Inf | < .001
## 0.94      |        0.15 | 0.02 | [0.11, 0.18] |  8.35 | Inf | < .001
## 0.97      |        0.14 | 0.02 | [0.11, 0.18] |  8.20 | Inf | < .001
## 1.00      |        0.14 | 0.02 | [0.11, 0.18] |  8.05 | Inf | < .001
## 1.02      |        0.14 | 0.02 | [0.11, 0.18] |  7.90 | Inf | < .001
## 1.05      |        0.14 | 0.02 | [0.11, 0.18] |  7.75 | Inf | < .001
## 1.08      |        0.14 | 0.02 | [0.11, 0.18] |  7.60 | Inf | < .001
## 1.10      |        0.14 | 0.02 | [0.10, 0.18] |  7.46 | Inf | < .001
## 1.13      |        0.14 | 0.02 | [0.10, 0.18] |  7.31 | Inf | < .001
## 1.16      |        0.14 | 0.02 | [0.10, 0.18] |  7.16 | Inf | < .001
## 1.18      |        0.14 | 0.02 | [0.10, 0.18] |  7.02 | Inf | < .001
## 1.21      |        0.14 | 0.02 | [0.10, 0.18] |  6.87 | Inf | < .001
## 1.24      |        0.14 | 0.02 | [0.10, 0.18] |  6.73 | Inf | < .001
## 1.26      |        0.14 | 0.02 | [0.10, 0.18] |  6.59 | Inf | < .001
## 1.29      |        0.14 | 0.02 | [0.09, 0.18] |  6.45 | Inf | < .001
## 1.32      |        0.13 | 0.02 | [0.09, 0.18] |  6.32 | Inf | < .001
## 1.34      |        0.13 | 0.02 | [0.09, 0.18] |  6.19 | Inf | < .001
## 1.37      |        0.13 | 0.02 | [0.09, 0.18] |  6.06 | Inf | < .001
## 1.40      |        0.13 | 0.02 | [0.09, 0.18] |  5.93 | Inf | < .001
## 1.42      |        0.13 | 0.02 | [0.09, 0.18] |  5.80 | Inf | < .001
## 1.45      |        0.13 | 0.02 | [0.09, 0.18] |  5.68 | Inf | < .001
## 1.48      |        0.13 | 0.02 | [0.08, 0.18] |  5.56 | Inf | < .001
## 1.50      |        0.13 | 0.02 | [0.08, 0.18] |  5.44 | Inf | < .001
## 1.53      |        0.13 | 0.02 | [0.08, 0.17] |  5.33 | Inf | < .001
## 1.56      |        0.13 | 0.02 | [0.08, 0.17] |  5.21 | Inf | < .001
## 1.58      |        0.13 | 0.02 | [0.08, 0.17] |  5.10 | Inf | < .001
## Marginal effects estimated for HomeEvic_22_R_Z
```

\# ADHD Strict Model
ADHD_Stringent_Adjusted_Age_Interact<- glmer(ADHD_22_R_Alt ~ raceASIA_22_F_L + famstruct5_22_F_L + AdultEduc_22_F_L + povlev4_22_F_L + sex_22_R_L + PlacesLived_22_R_F_L + MotherMH_22_R_F_L + EverHomeless_22_R_F_L + LowBWght_22_R_F_L + BornPre_22_R_F_L + FoodSit_22_R + ACE2more11_22_R_F_L + Child_Age * HomeEvic_22_R_Z + (1|FIPSST), family=binomial, data=NSCH_Topical_DRC_CAHMI_subset, control = glmerControl(optimizer = "bobyqa",optCtrl=list(maxfun=2e5)),nAGQ = 0)
summary(ADHD_Stringent_Adjusted_Age_Interact)

```
## Generalized linear mixed model fit by maximum likelihood (Adaptive
##   Gauss-Hermite Quadrature, nAGQ = 0) [glmerMod]
##  Family: binomial  ( logit )
## Formula:
## ADHD_22_R_Alt ~ raceASIA_22_F_L + famstruct5_22_F_L + AdultEduc_22_F_L +
##     povlev4_22_F_L + sex_22_R_L + PlacesLived_22_R_F_L + MotherMH_22_R_F_L +
##     EverHomeless_22_R_F_L + LowBWght_22_R_F_L + BornPre_22_R_F_L +
##     FoodSit_22_R + ACE2more11_22_R_F_L + Child_Age * HomeEvic_22_R_Z +
##     (1 | FIPSST)
##    Data: NSCH_Topical_DRC_CAHMI_subset
## Control: glmerControl(optimizer = "bobyqa", optCtrl = list(maxfun = 2e+05))
##
##      AIC      BIC   logLik deviance df.resid
##  22702.4  22949.1 -11322.2  22644.4    36609
##
## Scaled residuals:
##     Min      1Q  Median      3Q     Max
## -2.4633 -0.3731 -0.2629 -0.1833  8.8960
##
## Random effects:
##  Groups Name        Variance Std.Dev.
##  FIPSST (Intercept) 0.03806  0.1951
## Number of obs: 36638, groups:  FIPSST, 51
##
## Fixed effects:
##                                                      Estimate Std. Error
## (Intercept)                                          -2.87211    0.15562
## raceASIA_22_F_LHispanic                              -0.32873    0.05646
## raceASIA_22_F_LBlack, non-Hispanic                   -0.51718    0.08485
## raceASIA_22_F_LAsian, non-Hispanic                   -1.02497    0.11778
## raceASIA_22_F_LMulti-racial, non-Hispanic            -0.17949    0.06592
## famstruct5_22_F_LTwo parents, not currently married  -0.05319    0.07598
## famstruct5_22_F_LSingle parent (mother or father)    -0.13375    0.05037
## famstruct5_22_F_LGrandparent household              -11.34544  149.90866
## famstruct5_22_F_LOther family type                    0.94290    0.32435
## AdultEduc_22_F_LHigh school or GED                    0.24518    0.14210
## AdultEduc_22_F_LSome college or technical school      0.26456    0.13994
## AdultEduc_22_F_LCollege degree or higher              0.24858    0.14025
## povlev4_22_F_L100-199% FPL                           -0.13756    0.06648
## povlev4_22_F_L200-399% FPL                           -0.05211    0.06298
## povlev4_22_F_L400% FPL or greater                     0.02122    0.06633
## sex_22_R_LFemale                                     -0.71295    0.03648
## PlacesLived_22_R_F_L3 or more times                   0.20576    0.11137
## MotherMH_22_R_F_LGood                                 0.43819    0.04007
## MotherMH_22_R_F_LFair or poor                         0.50495    0.06085
## EverHomeless_22_R_F_LYes                              0.41438    0.09998
## LowBWght_22_R_F_LYes                                  0.25314    0.06709
## BornPre_22_R_F_LYes                                   0.16679    0.06036
## FoodSit_22_R                                          0.13523    0.03549
## ACE2more11_22_R_F_L1 ACE                              0.57845    0.04673
## ACE2more11_22_R_F_L2+ ACEs                            1.01981    0.05145
## Child_Age                                             0.55513    0.02222
## HomeEvic_22_R_Z                                       0.02913    0.02021
## Child_Age:HomeEvic_22_R_Z                            -0.02780    0.01871
##                                                     z value Pr(>|z|)
## (Intercept)                                         -18.456  < 2e-16 ***
## raceASIA_22_F_LHispanic                              -5.822 5.80e-09 ***
## raceASIA_22_F_LBlack, non-Hispanic                   -6.095 1.09e-09 ***
## raceASIA_22_F_LAsian, non-Hispanic                   -8.703  < 2e-16 ***
## raceASIA_22_F_LMulti-racial, non-Hispanic            -2.723 0.006474 **
## famstruct5_22_F_LTwo parents, not currently married  -0.700 0.483848
## famstruct5_22_F_LSingle parent (mother or father)    -2.655 0.007922 **
## famstruct5_22_F_LGrandparent household               -0.076 0.939672
## famstruct5_22_F_LOther family type                    2.907 0.003649 **
## AdultEduc_22_F_LHigh school or GED                    1.725 0.084456 .
## AdultEduc_22_F_LSome college or technical school      1.890 0.058693 .
## AdultEduc_22_F_LCollege degree or higher              1.772 0.076331 .
## povlev4_22_F_L100-199% FPL                           -2.069 0.038545 *
## povlev4_22_F_L200-399% FPL                           -0.827 0.408038
## povlev4_22_F_L400% FPL or greater                     0.320 0.749026
## sex_22_R_LFemale                                    -19.541  < 2e-16 ***
## PlacesLived_22_R_F_L3 or more times                   1.848 0.064670 .
## MotherMH_22_R_F_LGood                                10.935  < 2e-16 ***
## MotherMH_22_R_F_LFair or poor                         8.299  < 2e-16 ***
## EverHomeless_22_R_F_LYes                              4.144 3.41e-05 ***
## LowBWght_22_R_F_LYes                                  3.773 0.000161 ***
## BornPre_22_R_F_LYes                                   2.763 0.005721 **
## FoodSit_22_R                                          3.811 0.000138 ***
## ACE2more11_22_R_F_L1 ACE                             12.379  < 2e-16 ***
## ACE2more11_22_R_F_L2+ ACEs                           19.821  < 2e-16 ***
## Child_Age                                            24.987  < 2e-16 ***
## HomeEvic_22_R_Z                                       1.441 0.149448
## Child_Age:HomeEvic_22_R_Z                            -1.486 0.137340
## ---
## Signif. codes:  0 '***' 0.001 '**' 0.01 '*' 0.05 '.' 0.1 ' ' 1
##
## Correlation matrix not shown by default, as p = 28 > 12.
## Use print(x, correlation=TRUE)  or
##     vcov(x)        if you need it
```

\#
modelbased::estimate_slopes(ADHD_Stringent_Adjusted_Age_Interact, trend = "HomeEvic_22_R_Z", by = "Child_Age",nuisance=c("raceASIA_22_F_L","famstruct5_22_F_L","AdultEduc_22_F_L","povlev4_22_F_L","sex_22_R_L","PlacesLived_22_R_F_L","MotherMH_22_R_F_L","EverHomeless_22_R_F_L","LowBWght_22_R_F_L","BornPre_22_R_F_L","FoodSit_22_R","ACE2more11_22_R_F_L"), length = 100)

```
## Estimated Marginal Effects
##
## Child_Age | Coefficient |   SE |        95% CI |         z |  df |     p
## ------------------------------------------------------------------------
## -1.06     |        0.06 | 0.03 | [-0.01, 0.13] |      1.70 | Inf | 0.089
## -1.03     |        0.06 | 0.03 | [-0.01, 0.12] |      1.70 | Inf | 0.089
## -1.00     |        0.06 | 0.03 | [-0.01, 0.12] |      1.70 | Inf | 0.089
## -0.98     |        0.06 | 0.03 | [-0.01, 0.12] |      1.70 | Inf | 0.089
## -0.95     |        0.06 | 0.03 | [-0.01, 0.12] |      1.70 | Inf | 0.089
## -0.92     |        0.05 | 0.03 | [-0.01, 0.12] |      1.70 | Inf | 0.090
## -0.90     |        0.05 | 0.03 | [-0.01, 0.12] |      1.70 | Inf | 0.090
## -0.87     |        0.05 | 0.03 | [-0.01, 0.11] |      1.70 | Inf | 0.090
## -0.84     |        0.05 | 0.03 | [-0.01, 0.11] |      1.69 | Inf | 0.090
## -0.82     |        0.05 | 0.03 | [-0.01, 0.11] |      1.69 | Inf | 0.091
## -0.79     |        0.05 | 0.03 | [-0.01, 0.11] |      1.69 | Inf | 0.091
## -0.76     |        0.05 | 0.03 | [-0.01, 0.11] |      1.69 | Inf | 0.091
## -0.74     |        0.05 | 0.03 | [-0.01, 0.11] |      1.69 | Inf | 0.092
## -0.71     |        0.05 | 0.03 | [-0.01, 0.11] |      1.68 | Inf | 0.092
## -0.68     |        0.05 | 0.03 | [-0.01, 0.10] |      1.68 | Inf | 0.093
## -0.66     |        0.05 | 0.03 | [-0.01, 0.10] |      1.68 | Inf | 0.093
## -0.63     |        0.05 | 0.03 | [-0.01, 0.10] |      1.67 | Inf | 0.094
## -0.60     |        0.05 | 0.03 | [-0.01, 0.10] |      1.67 | Inf | 0.095
## -0.58     |        0.05 | 0.03 | [-0.01, 0.10] |      1.67 | Inf | 0.096
## -0.55     |        0.04 | 0.03 | [-0.01, 0.10] |      1.66 | Inf | 0.096
## -0.52     |        0.04 | 0.03 | [-0.01, 0.10] |      1.66 | Inf | 0.097
## -0.50     |        0.04 | 0.03 | [-0.01, 0.09] |      1.65 | Inf | 0.098
## -0.47     |        0.04 | 0.03 | [-0.01, 0.09] |      1.65 | Inf | 0.100
## -0.44     |        0.04 | 0.03 | [-0.01, 0.09] |      1.64 | Inf | 0.101
## -0.42     |        0.04 | 0.02 | [-0.01, 0.09] |      1.63 | Inf | 0.102
## -0.39     |        0.04 | 0.02 | [-0.01, 0.09] |      1.63 | Inf | 0.104
## -0.36     |        0.04 | 0.02 | [-0.01, 0.09] |      1.62 | Inf | 0.105
## -0.34     |        0.04 | 0.02 | [-0.01, 0.09] |      1.61 | Inf | 0.107
## -0.31     |        0.04 | 0.02 | [-0.01, 0.08] |      1.60 | Inf | 0.109
## -0.28     |        0.04 | 0.02 | [-0.01, 0.08] |      1.59 | Inf | 0.111
## -0.26     |        0.04 | 0.02 | [-0.01, 0.08] |      1.58 | Inf | 0.113
## -0.23     |        0.04 | 0.02 | [-0.01, 0.08] |      1.57 | Inf | 0.116
## -0.20     |        0.03 | 0.02 | [-0.01, 0.08] |      1.56 | Inf | 0.119
## -0.18     |        0.03 | 0.02 | [-0.01, 0.08] |      1.55 | Inf | 0.122
## -0.15     |        0.03 | 0.02 | [-0.01, 0.08] |      1.53 | Inf | 0.125
## -0.12     |        0.03 | 0.02 | [-0.01, 0.07] |      1.52 | Inf | 0.128
## -0.10     |        0.03 | 0.02 | [-0.01, 0.07] |      1.51 | Inf | 0.132
## -0.07     |        0.03 | 0.02 | [-0.01, 0.07] |      1.49 | Inf | 0.136
## -0.04     |        0.03 | 0.02 | [-0.01, 0.07] |      1.47 | Inf | 0.141
## -0.02     |        0.03 | 0.02 | [-0.01, 0.07] |      1.45 | Inf | 0.146
## 9.73e-03  |        0.03 | 0.02 | [-0.01, 0.07] |      1.43 | Inf | 0.152
## 0.04      |        0.03 | 0.02 | [-0.01, 0.07] |      1.41 | Inf | 0.157
## 0.06      |        0.03 | 0.02 | [-0.01, 0.07] |      1.39 | Inf | 0.164
## 0.09      |        0.03 | 0.02 | [-0.01, 0.06] |      1.37 | Inf | 0.171
## 0.12      |        0.03 | 0.02 | [-0.01, 0.06] |      1.34 | Inf | 0.179
## 0.14      |        0.03 | 0.02 | [-0.01, 0.06] |      1.32 | Inf | 0.187
## 0.17      |        0.02 | 0.02 | [-0.01, 0.06] |      1.29 | Inf | 0.196
## 0.20      |        0.02 | 0.02 | [-0.01, 0.06] |      1.26 | Inf | 0.206
## 0.22      |        0.02 | 0.02 | [-0.01, 0.06] |      1.23 | Inf | 0.217
## 0.25      |        0.02 | 0.02 | [-0.01, 0.06] |      1.20 | Inf | 0.228
## 0.28      |        0.02 | 0.02 | [-0.01, 0.06] |      1.17 | Inf | 0.241
## 0.30      |        0.02 | 0.02 | [-0.01, 0.06] |      1.14 | Inf | 0.255
## 0.33      |        0.02 | 0.02 | [-0.02, 0.06] |      1.10 | Inf | 0.269
## 0.36      |        0.02 | 0.02 | [-0.02, 0.05] |      1.07 | Inf | 0.285
## 0.38      |        0.02 | 0.02 | [-0.02, 0.05] |      1.03 | Inf | 0.302
## 0.41      |        0.02 | 0.02 | [-0.02, 0.05] |      0.99 | Inf | 0.320
## 0.44      |        0.02 | 0.02 | [-0.02, 0.05] |      0.96 | Inf | 0.339
## 0.46      |        0.02 | 0.02 | [-0.02, 0.05] |      0.92 | Inf | 0.360
## 0.49      |        0.02 | 0.02 | [-0.02, 0.05] |      0.88 | Inf | 0.381
## 0.52      |        0.01 | 0.02 | [-0.02, 0.05] |      0.83 | Inf | 0.404
## 0.54      |        0.01 | 0.02 | [-0.02, 0.05] |      0.79 | Inf | 0.428
## 0.57      |        0.01 | 0.02 | [-0.02, 0.05] |      0.75 | Inf | 0.454
## 0.60      |        0.01 | 0.02 | [-0.02, 0.05] |      0.71 | Inf | 0.480
## 0.62      |        0.01 | 0.02 | [-0.02, 0.05] |      0.66 | Inf | 0.507
## 0.65      |        0.01 | 0.02 | [-0.02, 0.05] |      0.62 | Inf | 0.536
## 0.68      |        0.01 | 0.02 | [-0.02, 0.05] |      0.58 | Inf | 0.565
## 0.70      |    9.60e-03 | 0.02 | [-0.03, 0.05] |      0.53 | Inf | 0.595
## 0.73      |    8.86e-03 | 0.02 | [-0.03, 0.04] |      0.49 | Inf | 0.626
## 0.76      |    8.12e-03 | 0.02 | [-0.03, 0.04] |      0.44 | Inf | 0.657
## 0.78      |    7.38e-03 | 0.02 | [-0.03, 0.04] |      0.40 | Inf | 0.689
## 0.81      |    6.64e-03 | 0.02 | [-0.03, 0.04] |      0.36 | Inf | 0.720
## 0.84      |    5.90e-03 | 0.02 | [-0.03, 0.04] |      0.32 | Inf | 0.752
## 0.86      |    5.16e-03 | 0.02 | [-0.03, 0.04] |      0.27 | Inf | 0.785
## 0.89      |    4.42e-03 | 0.02 | [-0.03, 0.04] |      0.23 | Inf | 0.817
## 0.92      |    3.67e-03 | 0.02 | [-0.03, 0.04] |      0.19 | Inf | 0.848
## 0.94      |    2.93e-03 | 0.02 | [-0.04, 0.04] |      0.15 | Inf | 0.880
## 0.97      |    2.19e-03 | 0.02 | [-0.04, 0.04] |      0.11 | Inf | 0.911
## 1.00      |    1.45e-03 | 0.02 | [-0.04, 0.04] |      0.07 | Inf | 0.942
## 1.02      |    7.12e-04 | 0.02 | [-0.04, 0.04] |      0.04 | Inf | 0.972
## 1.05      |   -2.91e-05 | 0.02 | [-0.04, 0.04] | -1.43e-03 | Inf | 0.999
## 1.08      |   -7.70e-04 | 0.02 | [-0.04, 0.04] |     -0.04 | Inf | 0.970
## 1.10      |   -1.51e-03 | 0.02 | [-0.04, 0.04] |     -0.07 | Inf | 0.942
## 1.13      |   -2.25e-03 | 0.02 | [-0.04, 0.04] |     -0.11 | Inf | 0.915
## 1.16      |   -2.99e-03 | 0.02 | [-0.04, 0.04] |     -0.14 | Inf | 0.889
## 1.18      |   -3.73e-03 | 0.02 | [-0.05, 0.04] |     -0.17 | Inf | 0.863
## 1.21      |   -4.47e-03 | 0.02 | [-0.05, 0.04] |     -0.20 | Inf | 0.838
## 1.24      |   -5.21e-03 | 0.02 | [-0.05, 0.04] |     -0.23 | Inf | 0.815
## 1.26      |   -5.96e-03 | 0.02 | [-0.05, 0.04] |     -0.26 | Inf | 0.792
## 1.29      |   -6.70e-03 | 0.02 | [-0.05, 0.04] |     -0.29 | Inf | 0.769
## 1.32      |   -7.44e-03 | 0.02 | [-0.05, 0.04] |     -0.32 | Inf | 0.748
## 1.34      |   -8.18e-03 | 0.02 | [-0.05, 0.04] |     -0.35 | Inf | 0.728
## 1.37      |   -8.92e-03 | 0.02 | [-0.06, 0.04] |     -0.37 | Inf | 0.708
## 1.40      |   -9.66e-03 | 0.02 | [-0.06, 0.04] |     -0.40 | Inf | 0.689
## 1.42      |       -0.01 | 0.02 | [-0.06, 0.04] |     -0.42 | Inf | 0.671
## 1.45      |       -0.01 | 0.02 | [-0.06, 0.04] |     -0.45 | Inf | 0.654
## 1.48      |       -0.01 | 0.03 | [-0.06, 0.04] |     -0.47 | Inf | 0.637
## 1.50      |       -0.01 | 0.03 | [-0.06, 0.04] |     -0.49 | Inf | 0.621
## 1.53      |       -0.01 | 0.03 | [-0.06, 0.04] |     -0.52 | Inf | 0.606
## 1.56      |       -0.01 | 0.03 | [-0.07, 0.04] |     -0.54 | Inf | 0.592
## 1.58      |       -0.01 | 0.03 | [-0.07, 0.04] |     -0.56 | Inf | 0.578
## Marginal effects estimated for HomeEvic_22_R_Z
```

**\# Table Making Related to ADHD (Table 04)**

\# Table with ADHD Base Model
ADHD_base_table <- tbl_regression(ADHD_Base_Adjusted_Age_Interact, exponentiate = TRUE) %>% bold_p()

\# Table with ADHD Strict Model
ADHD_stringent_table <- tbl_regression(ADHD_Stringent_Adjusted_Age_Interact, exponentiate = TRUE)%>% bold_p()

\# Combining ADHD Tables
ADHD_combined_table<-tbl_merge(
 tbls = list(ADHD_base_table, ADHD_stringent_table),
 tab_spanner = c("**Base Model**", "**Stringent Model**")
)

ADHD_combined_table

|  | Base Model | | | Stringent Model | | |
| --- | --- | --- | --- | --- | --- | --- |
| Characteristic | OR*^1^* | 95% CI*^1^* | p-value | OR*^1^* | 95% CI*^1^* | p-value |
| Child's Race/Ethnicity |  |  |  |  |  |  |
| White, non-Hispanic | — | — |  | — | — |  |
| Hispanic | 0.72 | 0.64, 0.80 | <0.001 | 0.72 | 0.64, 0.80 | <0.001 |
| Black, non-Hispanic | 0.57 | 0.48, 0.67 | <0.001 | 0.60 | 0.50, 0.70 | <0.001 |
| Asian, non-Hispanic | 0.31 | 0.25, 0.39 | <0.001 | 0.36 | 0.28, 0.45 | <0.001 |
| Multi-racial, non-Hispanic | 0.92 | 0.81, 1.04 | 0.2 | 0.84 | 0.73, 0.95 | 0.006 |
| Family Structure |  |  |  |  |  |  |
| Two parents, currently married | — | — |  | — | — |  |
| Two parents, not currently married | 1.33 | 1.16, 1.54 | <0.001 | 0.95 | 0.82, 1.10 | 0.5 |
| Single parent (mother or father) | 1.43 | 1.30, 1.56 | <0.001 | 0.87 | 0.79, 0.97 | 0.008 |
| Grandparent household | 0.00 | 0.00, | >0.9 | 0.00 | 0.00, | >0.9 |
| Other family type | 5.09 | 2.72, 9.52 | <0.001 | 2.57 | 1.36, 4.85 | 0.004 |
| Highest Level of Education in Household |  |  |  |  |  |  |
| Less than high school | — | — |  | — | — |  |
| High school or GED | 1.45 | 1.10, 1.90 | 0.008 | 1.28 | 0.97, 1.69 | 0.084 |
| Some college or technical school | 1.52 | 1.16, 1.98 | 0.002 | 1.30 | 0.99, 1.71 | 0.059 |
| College degree or higher | 1.33 | 1.02, 1.75 | 0.035 | 1.28 | 0.97, 1.69 | 0.076 |
| Household Poverty Status |  |  |  |  |  |  |
| 0-99% FPL | — | — |  | — | — |  |
| 100-199% FPL | 0.89 | 0.78, 1.01 | 0.074 | 0.87 | 0.77, 0.99 | 0.039 |
| 200-399% FPL | 0.91 | 0.81, 1.03 | 0.14 | 0.95 | 0.84, 1.07 | 0.4 |
| 400% FPL or greater | 0.87 | 0.77, 0.99 | 0.036 | 1.02 | 0.90, 1.16 | 0.7 |
| Child's Sex Assigned at Birth |  |  |  |  |  |  |
| Male | — | — |  | — | — |  |
| Female | 0.51 | 0.47, 0.54 | <0.001 | 0.49 | 0.46, 0.53 | <0.001 |
| Child's Age in Years | 1.86 | 1.78, 1.94 | <0.001 | 1.74 | 1.67, 1.82 | <0.001 |
| Eviction Stress/Concern | 1.19 | 1.15, 1.23 | <0.001 | 1.03 | 0.99, 1.07 | 0.15 |
| Child's Age in Years * Eviction Stress/Concern | 0.97 | 0.94, 1.01 | 0.095 | 0.97 | 0.94, 1.01 | 0.14 |
| Places Lived, Last Year |  |  |  |  |  |  |
| 0-2 times |  |  |  | — | — |  |
| 3 or more times |  |  |  | 1.23 | 0.99, 1.53 | 0.065 |
| Mother's Mental health |  |  |  |  |  |  |
| Excellent or very good |  |  |  | — | — |  |
| Good |  |  |  | 1.55 | 1.43, 1.68 | <0.001 |
| Fair or poor |  |  |  | 1.66 | 1.47, 1.87 | <0.001 |
| Ever Homeless |  |  |  |  |  |  |
| No |  |  |  | — | — |  |
| Yes |  |  |  | 1.51 | 1.24, 1.84 | <0.001 |
| Low Birth Weight |  |  |  |  |  |  |
| No |  |  |  | — | — |  |
| Yes |  |  |  | 1.29 | 1.13, 1.47 | <0.001 |
| Born Premature |  |  |  |  |  |  |
| No |  |  |  | — | — |  |
| Yes |  |  |  | 1.18 | 1.05, 1.33 | 0.006 |
| Food Insecurity |  |  |  | 1.14 | 1.07, 1.23 | <0.001 |
| Adverse Childhood Experiences |  |  |  |  |  |  |
| 0 ACEs |  |  |  | — | — |  |
| 1 ACE |  |  |  | 1.78 | 1.63, 1.95 | <0.001 |
| 2+ ACEs |  |  |  | 2.77 | 2.51, 3.07 | <0.001 |
| *^1^*OR = Odds Ratio, CI = Confidence Interval | | | | | | |

\# Output as Word Doc
ADHD_combined_table %>%
 gtsummary::as_gt() %>%
 gt::gtsave("Table4.docx")

**\# Analyses Related to Behavioral Problems**

\# Behavioral Problems Base Model
Behavior_Base_Adjusted_Age_Interact<-glmer(behavior_22_R_Alt ~ raceASIA_22_F_L + famstruct5_22_F_L + AdultEduc_22_F_L + povlev4_22_F_L + sex_22_R_L + Child_Age * HomeEvic_22_R_Z + (1|FIPSST), family=binomial, data=NSCH_Topical_DRC_CAHMI_subset, control = glmerControl(optimizer = "bobyqa",optCtrl=list(maxfun=2e5)),nAGQ = 0)
summary(Behavior_Base_Adjusted_Age_Interact)

```
## Generalized linear mixed model fit by maximum likelihood (Adaptive
##   Gauss-Hermite Quadrature, nAGQ = 0) [glmerMod]
##  Family: binomial  ( logit )
## Formula:
## behavior_22_R_Alt ~ raceASIA_22_F_L + famstruct5_22_F_L + AdultEduc_22_F_L +
##     povlev4_22_F_L + sex_22_R_L + Child_Age * HomeEvic_22_R_Z +
##     (1 | FIPSST)
##    Data: NSCH_Topical_DRC_CAHMI_subset
## Control: glmerControl(optimizer = "bobyqa", optCtrl = list(maxfun = 2e+05))
##
##      AIC      BIC   logLik deviance df.resid
##  18503.7  18673.9  -9231.9  18463.7    36618
##
## Scaled residuals:
##     Min      1Q  Median      3Q     Max
## -1.2783 -0.3129 -0.2525 -0.1932  8.7616
##
## Random effects:
##  Groups Name        Variance Std.Dev.
##  FIPSST (Intercept) 0.01146  0.1071
## Number of obs: 36638, groups:  FIPSST, 51
##
## Fixed effects:
##                                                      Estimate Std. Error
## (Intercept)                                         -2.101732   0.145282
## raceASIA_22_F_LHispanic                             -0.169596   0.061899
## raceASIA_22_F_LBlack, non-Hispanic                  -0.213956   0.090250
## raceASIA_22_F_LAsian, non-Hispanic                  -0.885427   0.129143
## raceASIA_22_F_LMulti-racial, non-Hispanic           -0.044336   0.074398
## famstruct5_22_F_LTwo parents, not currently married  0.261938   0.082319
## famstruct5_22_F_LSingle parent (mother or father)    0.421056   0.053425
## famstruct5_22_F_LGrandparent household               0.935312   1.188768
## famstruct5_22_F_LOther family type                   2.245111   0.300651
## AdultEduc_22_F_LHigh school or GED                   0.048440   0.144698
## AdultEduc_22_F_LSome college or technical school     0.128835   0.141714
## AdultEduc_22_F_LCollege degree or higher            -0.007492   0.142393
## povlev4_22_F_L100-199% FPL                          -0.057189   0.071822
## povlev4_22_F_L200-399% FPL                          -0.149583   0.069917
## povlev4_22_F_L400% FPL or greater                   -0.242127   0.073892
## sex_22_R_LFemale                                    -0.897001   0.043880
## Child_Age                                            0.057504   0.023834
## HomeEvic_22_R_Z                                      0.199886   0.017925
## Child_Age:HomeEvic_22_R_Z                           -0.003231   0.018500
##                                                     z value Pr(>|z|)
## (Intercept)                                         -14.467  < 2e-16 ***
## raceASIA_22_F_LHispanic                              -2.740  0.00615 **
## raceASIA_22_F_LBlack, non-Hispanic                   -2.371  0.01775 *
## raceASIA_22_F_LAsian, non-Hispanic                   -6.856 7.07e-12 ***
## raceASIA_22_F_LMulti-racial, non-Hispanic            -0.596  0.55122
## famstruct5_22_F_LTwo parents, not currently married   3.182  0.00146 **
## famstruct5_22_F_LSingle parent (mother or father)     7.881 3.24e-15 ***
## famstruct5_22_F_LGrandparent household                0.787  0.43140
## famstruct5_22_F_LOther family type                    7.467 8.17e-14 ***
## AdultEduc_22_F_LHigh school or GED                    0.335  0.73780
## AdultEduc_22_F_LSome college or technical school      0.909  0.36328
## AdultEduc_22_F_LCollege degree or higher             -0.053  0.95804
## povlev4_22_F_L100-199% FPL                           -0.796  0.42588
## povlev4_22_F_L200-399% FPL                           -2.139  0.03240 *
## povlev4_22_F_L400% FPL or greater                    -3.277  0.00105 **
## sex_22_R_LFemale                                    -20.442  < 2e-16 ***
## Child_Age                                             2.413  0.01583 *
## HomeEvic_22_R_Z                                      11.151  < 2e-16 ***
## Child_Age:HomeEvic_22_R_Z                            -0.175  0.86135
## ---
## Signif. codes:  0 '***' 0.001 '**' 0.01 '*' 0.05 '.' 0.1 ' ' 1
##
## Correlation matrix not shown by default, as p = 19 > 12.
## Use print(x, correlation=TRUE)  or
##     vcov(x)        if you need it
```

\#
modelbased::estimate_slopes(Behavior_Base_Adjusted_Age_Interact, trend = "HomeEvic_22_R_Z", by ="Child_Age",nuisance=c("raceASIA_22_F_L","famstruct5_22_F_L","AdultEduc_22_F_L","povlev4_22_F_L","sex_22_R_L"),length = 100)

```
## Estimated Marginal Effects
##
## Child_Age | Coefficient |   SE |       95% CI |     z |  df |      p
## --------------------------------------------------------------------
## -1.06     |        0.20 | 0.03 | [0.14, 0.26] |  6.61 | Inf | < .001
## -1.03     |        0.20 | 0.03 | [0.14, 0.26] |  6.69 | Inf | < .001
## -1.00     |        0.20 | 0.03 | [0.14, 0.26] |  6.78 | Inf | < .001
## -0.98     |        0.20 | 0.03 | [0.15, 0.26] |  6.87 | Inf | < .001
## -0.95     |        0.20 | 0.03 | [0.15, 0.26] |  6.97 | Inf | < .001
## -0.92     |        0.20 | 0.03 | [0.15, 0.26] |  7.06 | Inf | < .001
## -0.90     |        0.20 | 0.03 | [0.15, 0.26] |  7.16 | Inf | < .001
## -0.87     |        0.20 | 0.03 | [0.15, 0.26] |  7.25 | Inf | < .001
## -0.84     |        0.20 | 0.03 | [0.15, 0.26] |  7.35 | Inf | < .001
## -0.82     |        0.20 | 0.03 | [0.15, 0.26] |  7.46 | Inf | < .001
## -0.79     |        0.20 | 0.03 | [0.15, 0.25] |  7.56 | Inf | < .001
## -0.76     |        0.20 | 0.03 | [0.15, 0.25] |  7.67 | Inf | < .001
## -0.74     |        0.20 | 0.03 | [0.15, 0.25] |  7.78 | Inf | < .001
## -0.71     |        0.20 | 0.03 | [0.15, 0.25] |  7.89 | Inf | < .001
## -0.68     |        0.20 | 0.03 | [0.15, 0.25] |  8.00 | Inf | < .001
## -0.66     |        0.20 | 0.02 | [0.15, 0.25] |  8.11 | Inf | < .001
## -0.63     |        0.20 | 0.02 | [0.15, 0.25] |  8.23 | Inf | < .001
## -0.60     |        0.20 | 0.02 | [0.15, 0.25] |  8.35 | Inf | < .001
## -0.58     |        0.20 | 0.02 | [0.16, 0.25] |  8.47 | Inf | < .001
## -0.55     |        0.20 | 0.02 | [0.16, 0.25] |  8.59 | Inf | < .001
## -0.52     |        0.20 | 0.02 | [0.16, 0.25] |  8.71 | Inf | < .001
## -0.50     |        0.20 | 0.02 | [0.16, 0.25] |  8.83 | Inf | < .001
## -0.47     |        0.20 | 0.02 | [0.16, 0.25] |  8.96 | Inf | < .001
## -0.44     |        0.20 | 0.02 | [0.16, 0.24] |  9.09 | Inf | < .001
## -0.42     |        0.20 | 0.02 | [0.16, 0.24] |  9.22 | Inf | < .001
## -0.39     |        0.20 | 0.02 | [0.16, 0.24] |  9.34 | Inf | < .001
## -0.36     |        0.20 | 0.02 | [0.16, 0.24] |  9.47 | Inf | < .001
## -0.34     |        0.20 | 0.02 | [0.16, 0.24] |  9.61 | Inf | < .001
## -0.31     |        0.20 | 0.02 | [0.16, 0.24] |  9.74 | Inf | < .001
## -0.28     |        0.20 | 0.02 | [0.16, 0.24] |  9.87 | Inf | < .001
## -0.26     |        0.20 | 0.02 | [0.16, 0.24] | 10.00 | Inf | < .001
## -0.23     |        0.20 | 0.02 | [0.16, 0.24] | 10.13 | Inf | < .001
## -0.20     |        0.20 | 0.02 | [0.16, 0.24] | 10.26 | Inf | < .001
## -0.18     |        0.20 | 0.02 | [0.16, 0.24] | 10.38 | Inf | < .001
## -0.15     |        0.20 | 0.02 | [0.16, 0.24] | 10.51 | Inf | < .001
## -0.12     |        0.20 | 0.02 | [0.16, 0.24] | 10.63 | Inf | < .001
## -0.10     |        0.20 | 0.02 | [0.16, 0.24] | 10.75 | Inf | < .001
## -0.07     |        0.20 | 0.02 | [0.16, 0.24] | 10.87 | Inf | < .001
## -0.04     |        0.20 | 0.02 | [0.16, 0.24] | 10.98 | Inf | < .001
## -0.02     |        0.20 | 0.02 | [0.16, 0.24] | 11.09 | Inf | < .001
## 9.73e-03  |        0.20 | 0.02 | [0.16, 0.23] | 11.19 | Inf | < .001
## 0.04      |        0.20 | 0.02 | [0.17, 0.23] | 11.28 | Inf | < .001
## 0.06      |        0.20 | 0.02 | [0.17, 0.23] | 11.38 | Inf | < .001
## 0.09      |        0.20 | 0.02 | [0.17, 0.23] | 11.46 | Inf | < .001
## 0.12      |        0.20 | 0.02 | [0.17, 0.23] | 11.54 | Inf | < .001
## 0.14      |        0.20 | 0.02 | [0.17, 0.23] | 11.60 | Inf | < .001
## 0.17      |        0.20 | 0.02 | [0.17, 0.23] | 11.66 | Inf | < .001
## 0.20      |        0.20 | 0.02 | [0.17, 0.23] | 11.72 | Inf | < .001
## 0.22      |        0.20 | 0.02 | [0.17, 0.23] | 11.76 | Inf | < .001
## 0.25      |        0.20 | 0.02 | [0.17, 0.23] | 11.79 | Inf | < .001
## 0.28      |        0.20 | 0.02 | [0.17, 0.23] | 11.81 | Inf | < .001
## 0.30      |        0.20 | 0.02 | [0.17, 0.23] | 11.83 | Inf | < .001
## 0.33      |        0.20 | 0.02 | [0.17, 0.23] | 11.83 | Inf | < .001
## 0.36      |        0.20 | 0.02 | [0.17, 0.23] | 11.82 | Inf | < .001
## 0.38      |        0.20 | 0.02 | [0.17, 0.23] | 11.81 | Inf | < .001
## 0.41      |        0.20 | 0.02 | [0.17, 0.23] | 11.78 | Inf | < .001
## 0.44      |        0.20 | 0.02 | [0.17, 0.23] | 11.74 | Inf | < .001
## 0.46      |        0.20 | 0.02 | [0.17, 0.23] | 11.69 | Inf | < .001
## 0.49      |        0.20 | 0.02 | [0.16, 0.23] | 11.64 | Inf | < .001
## 0.52      |        0.20 | 0.02 | [0.16, 0.23] | 11.57 | Inf | < .001
## 0.54      |        0.20 | 0.02 | [0.16, 0.23] | 11.50 | Inf | < .001
## 0.57      |        0.20 | 0.02 | [0.16, 0.23] | 11.42 | Inf | < .001
## 0.60      |        0.20 | 0.02 | [0.16, 0.23] | 11.33 | Inf | < .001
## 0.62      |        0.20 | 0.02 | [0.16, 0.23] | 11.23 | Inf | < .001
## 0.65      |        0.20 | 0.02 | [0.16, 0.23] | 11.13 | Inf | < .001
## 0.68      |        0.20 | 0.02 | [0.16, 0.23] | 11.02 | Inf | < .001
## 0.70      |        0.20 | 0.02 | [0.16, 0.23] | 10.91 | Inf | < .001
## 0.73      |        0.20 | 0.02 | [0.16, 0.23] | 10.79 | Inf | < .001
## 0.76      |        0.20 | 0.02 | [0.16, 0.23] | 10.67 | Inf | < .001
## 0.78      |        0.20 | 0.02 | [0.16, 0.23] | 10.54 | Inf | < .001
## 0.81      |        0.20 | 0.02 | [0.16, 0.23] | 10.42 | Inf | < .001
## 0.84      |        0.20 | 0.02 | [0.16, 0.23] | 10.29 | Inf | < .001
## 0.86      |        0.20 | 0.02 | [0.16, 0.24] | 10.15 | Inf | < .001
## 0.89      |        0.20 | 0.02 | [0.16, 0.24] | 10.02 | Inf | < .001
## 0.92      |        0.20 | 0.02 | [0.16, 0.24] |  9.88 | Inf | < .001
## 0.94      |        0.20 | 0.02 | [0.16, 0.24] |  9.75 | Inf | < .001
## 0.97      |        0.20 | 0.02 | [0.16, 0.24] |  9.61 | Inf | < .001
## 1.00      |        0.20 | 0.02 | [0.16, 0.24] |  9.48 | Inf | < .001
## 1.02      |        0.20 | 0.02 | [0.16, 0.24] |  9.34 | Inf | < .001
## 1.05      |        0.20 | 0.02 | [0.15, 0.24] |  9.20 | Inf | < .001
## 1.08      |        0.20 | 0.02 | [0.15, 0.24] |  9.07 | Inf | < .001
## 1.10      |        0.20 | 0.02 | [0.15, 0.24] |  8.94 | Inf | < .001
## 1.13      |        0.20 | 0.02 | [0.15, 0.24] |  8.80 | Inf | < .001
## 1.16      |        0.20 | 0.02 | [0.15, 0.24] |  8.67 | Inf | < .001
## 1.18      |        0.20 | 0.02 | [0.15, 0.24] |  8.54 | Inf | < .001
## 1.21      |        0.20 | 0.02 | [0.15, 0.24] |  8.41 | Inf | < .001
## 1.24      |        0.20 | 0.02 | [0.15, 0.24] |  8.29 | Inf | < .001
## 1.26      |        0.20 | 0.02 | [0.15, 0.24] |  8.16 | Inf | < .001
## 1.29      |        0.20 | 0.02 | [0.15, 0.24] |  8.04 | Inf | < .001
## 1.32      |        0.20 | 0.02 | [0.15, 0.24] |  7.92 | Inf | < .001
## 1.34      |        0.20 | 0.03 | [0.15, 0.24] |  7.80 | Inf | < .001
## 1.37      |        0.20 | 0.03 | [0.15, 0.25] |  7.69 | Inf | < .001
## 1.40      |        0.20 | 0.03 | [0.14, 0.25] |  7.57 | Inf | < .001
## 1.42      |        0.20 | 0.03 | [0.14, 0.25] |  7.46 | Inf | < .001
## 1.45      |        0.20 | 0.03 | [0.14, 0.25] |  7.35 | Inf | < .001
## 1.48      |        0.20 | 0.03 | [0.14, 0.25] |  7.24 | Inf | < .001
## 1.50      |        0.20 | 0.03 | [0.14, 0.25] |  7.14 | Inf | < .001
## 1.53      |        0.19 | 0.03 | [0.14, 0.25] |  7.03 | Inf | < .001
## 1.56      |        0.19 | 0.03 | [0.14, 0.25] |  6.93 | Inf | < .001
## 1.58      |        0.19 | 0.03 | [0.14, 0.25] |  6.83 | Inf | < .001
## Marginal effects estimated for HomeEvic_22_R_Z
```

\# Behavioral Problems Strict Model
Behavior_Stringent_Adjusted_Age_Interact<- glmer(behavior_22_R_Alt ~ raceASIA_22_F_L + famstruct5_22_F_L + AdultEduc_22_F_L + povlev4_22_F_L + sex_22_R_L + PlacesLived_22_R_F_L + MotherMH_22_R_F_L + EverHomeless_22_R_F_L + LowBWght_22_R_F_L + BornPre_22_R_F_L + FoodSit_22_R + ACE2more11_22_R_F_L + Child_Age * HomeEvic_22_R_Z + (1|FIPSST), family=binomial, data=NSCH_Topical_DRC_CAHMI_subset, control = glmerControl(optimizer = "bobyqa",optCtrl=list(maxfun=2e5)),nAGQ = 0)
summary(Behavior_Stringent_Adjusted_Age_Interact)

```
## Generalized linear mixed model fit by maximum likelihood (Adaptive
##   Gauss-Hermite Quadrature, nAGQ = 0) [glmerMod]
##  Family: binomial  ( logit )
## Formula:
## behavior_22_R_Alt ~ raceASIA_22_F_L + famstruct5_22_F_L + AdultEduc_22_F_L +
##     povlev4_22_F_L + sex_22_R_L + PlacesLived_22_R_F_L + MotherMH_22_R_F_L +
##     EverHomeless_22_R_F_L + LowBWght_22_R_F_L + BornPre_22_R_F_L +
##     FoodSit_22_R + ACE2more11_22_R_F_L + Child_Age * HomeEvic_22_R_Z +
##     (1 | FIPSST)
##    Data: NSCH_Topical_DRC_CAHMI_subset
## Control: glmerControl(optimizer = "bobyqa", optCtrl = list(maxfun = 2e+05))
##
##      AIC      BIC   logLik deviance df.resid
##  17310.2  17557.0  -8626.1  17252.2    36609
##
## Scaled residuals:
##     Min      1Q  Median      3Q     Max
## -1.7406 -0.2962 -0.2179 -0.1470  8.6389
##
## Random effects:
##  Groups Name        Variance Std.Dev.
##  FIPSST (Intercept) 0.009981 0.0999
## Number of obs: 36638, groups:  FIPSST, 51
##
## Fixed effects:
##                                                      Estimate Std. Error
## (Intercept)                                         -3.039533   0.163994
## raceASIA_22_F_LHispanic                             -0.151375   0.062838
## raceASIA_22_F_LBlack, non-Hispanic                  -0.088003   0.091813
## raceASIA_22_F_LAsian, non-Hispanic                  -0.645327   0.130724
## raceASIA_22_F_LMulti-racial, non-Hispanic           -0.160628   0.076286
## famstruct5_22_F_LTwo parents, not currently married -0.157806   0.085504
## famstruct5_22_F_LSingle parent (mother or father)   -0.199647   0.058200
## famstruct5_22_F_LGrandparent household               0.580602   1.324875
## famstruct5_22_F_LOther family type                   1.381506   0.317770
## AdultEduc_22_F_LHigh school or GED                  -0.119970   0.150101
## AdultEduc_22_F_LSome college or technical school    -0.071449   0.147271
## AdultEduc_22_F_LCollege degree or higher            -0.055942   0.148047
## povlev4_22_F_L100-199% FPL                          -0.061058   0.074351
## povlev4_22_F_L200-399% FPL                          -0.078010   0.072508
## povlev4_22_F_L400% FPL or greater                    0.006694   0.077485
## sex_22_R_LFemale                                    -0.943549   0.044906
## PlacesLived_22_R_F_L3 or more times                  0.405850   0.117317
## MotherMH_22_R_F_LGood                                0.647943   0.047177
## MotherMH_22_R_F_LFair or poor                        0.926568   0.064474
## EverHomeless_22_R_F_LYes                             0.511822   0.104592
## LowBWght_22_R_F_LYes                                 0.325045   0.076801
## BornPre_22_R_F_LYes                                  0.133824   0.070260
## FoodSit_22_R                                         0.167332   0.039716
## ACE2more11_22_R_F_L1 ACE                             0.695037   0.056919
## ACE2more11_22_R_F_L2+ ACEs                           1.336998   0.060816
## Child_Age                                           -0.058138   0.025538
## HomeEvic_22_R_Z                                      0.002492   0.020426
## Child_Age:HomeEvic_22_R_Z                            0.003092   0.019619
##                                                     z value Pr(>|z|)
## (Intercept)                                         -18.534  < 2e-16 ***
## raceASIA_22_F_LHispanic                              -2.409 0.015997 *
## raceASIA_22_F_LBlack, non-Hispanic                   -0.959 0.337806
## raceASIA_22_F_LAsian, non-Hispanic                   -4.937 7.95e-07 ***
## raceASIA_22_F_LMulti-racial, non-Hispanic            -2.106 0.035239 *
## famstruct5_22_F_LTwo parents, not currently married  -1.846 0.064949 .
## famstruct5_22_F_LSingle parent (mother or father)    -3.430 0.000603 ***
## famstruct5_22_F_LGrandparent household                0.438 0.661218
## famstruct5_22_F_LOther family type                    4.348 1.38e-05 ***
## AdultEduc_22_F_LHigh school or GED                   -0.799 0.424139
## AdultEduc_22_F_LSome college or technical school     -0.485 0.627568
## AdultEduc_22_F_LCollege degree or higher             -0.378 0.705529
## povlev4_22_F_L100-199% FPL                           -0.821 0.411520
## povlev4_22_F_L200-399% FPL                           -1.076 0.281983
## povlev4_22_F_L400% FPL or greater                     0.086 0.931160
## sex_22_R_LFemale                                    -21.012  < 2e-16 ***
## PlacesLived_22_R_F_L3 or more times                   3.459 0.000541 ***
## MotherMH_22_R_F_LGood                                13.734  < 2e-16 ***
## MotherMH_22_R_F_LFair or poor                        14.371  < 2e-16 ***
## EverHomeless_22_R_F_LYes                              4.894 9.91e-07 ***
## LowBWght_22_R_F_LYes                                  4.232 2.31e-05 ***
## BornPre_22_R_F_LYes                                   1.905 0.056819 .
## FoodSit_22_R                                          4.213 2.52e-05 ***
## ACE2more11_22_R_F_L1 ACE                             12.211  < 2e-16 ***
## ACE2more11_22_R_F_L2+ ACEs                           21.984  < 2e-16 ***
## Child_Age                                            -2.277 0.022813 *
## HomeEvic_22_R_Z                                       0.122 0.902888
## Child_Age:HomeEvic_22_R_Z                             0.158 0.874784
## ---
## Signif. codes:  0 '***' 0.001 '**' 0.01 '*' 0.05 '.' 0.1 ' ' 1
##
## Correlation matrix not shown by default, as p = 28 > 12.
## Use print(x, correlation=TRUE)  or
##     vcov(x)        if you need it
```

\#
modelbased::estimate_slopes(Behavior_Stringent_Adjusted_Age_Interact, trend = "HomeEvic_22_R_Z", by = "Child_Age",nuisance=c("raceASIA_22_F_L","famstruct5_22_F_L","AdultEduc_22_F_L","povlev4_22_F_L","sex_22_R_L","PlacesLived_22_R_F_L","MotherMH_22_R_F_L","EverHomeless_22_R_F_L","LowBWght_22_R_F_L","BornPre_22_R_F_L","FoodSit_22_R","ACE2more11_22_R_F_L"), length = 100)

```
## Estimated Marginal Effects
##
## Child_Age | Coefficient |   SE |        95% CI |         z |  df |      p
## -------------------------------------------------------------------------
## -1.06     |   -7.72e-04 | 0.03 | [-0.07, 0.06] |     -0.02 | Inf | 0.982
## -1.03     |   -6.90e-04 | 0.03 | [-0.07, 0.06] |     -0.02 | Inf | 0.983
## -1.00     |   -6.08e-04 | 0.03 | [-0.06, 0.06] |     -0.02 | Inf | 0.985
## -0.98     |   -5.25e-04 | 0.03 | [-0.06, 0.06] |     -0.02 | Inf | 0.987
## -0.95     |   -4.43e-04 | 0.03 | [-0.06, 0.06] |     -0.01 | Inf | 0.989
## -0.92     |   -3.61e-04 | 0.03 | [-0.06, 0.06] |     -0.01 | Inf | 0.991
## -0.90     |   -2.78e-04 | 0.03 | [-0.06, 0.06] | -8.98e-03 | Inf | 0.993
## -0.87     |   -1.96e-04 | 0.03 | [-0.06, 0.06] | -6.41e-03 | Inf | 0.995
## -0.84     |   -1.13e-04 | 0.03 | [-0.06, 0.06] | -3.76e-03 | Inf | 0.997
## -0.82     |   -3.11e-05 | 0.03 | [-0.06, 0.06] | -1.05e-03 | Inf | > .999
## -0.79     |    5.12e-05 | 0.03 | [-0.06, 0.06] |  1.75e-03 | Inf | 0.999
## -0.76     |    1.34e-04 | 0.03 | [-0.06, 0.06] |  4.61e-03 | Inf | 0.996
## -0.74     |    2.16e-04 | 0.03 | [-0.06, 0.06] |  7.56e-03 | Inf | 0.994
## -0.71     |    2.98e-04 | 0.03 | [-0.05, 0.06] |      0.01 | Inf | 0.992
## -0.68     |    3.81e-04 | 0.03 | [-0.05, 0.05] |      0.01 | Inf | 0.989
## -0.66     |    4.63e-04 | 0.03 | [-0.05, 0.05] |      0.02 | Inf | 0.987
## -0.63     |    5.45e-04 | 0.03 | [-0.05, 0.05] |      0.02 | Inf | 0.984
## -0.60     |    6.28e-04 | 0.03 | [-0.05, 0.05] |      0.02 | Inf | 0.981
## -0.58     |    7.10e-04 | 0.03 | [-0.05, 0.05] |      0.03 | Inf | 0.979
## -0.55     |    7.93e-04 | 0.03 | [-0.05, 0.05] |      0.03 | Inf | 0.976
## -0.52     |    8.75e-04 | 0.03 | [-0.05, 0.05] |      0.03 | Inf | 0.973
## -0.50     |    9.57e-04 | 0.03 | [-0.05, 0.05] |      0.04 | Inf | 0.970
## -0.47     |    1.04e-03 | 0.02 | [-0.05, 0.05] |      0.04 | Inf | 0.967
## -0.44     |    1.12e-03 | 0.02 | [-0.05, 0.05] |      0.05 | Inf | 0.964
## -0.42     |    1.20e-03 | 0.02 | [-0.05, 0.05] |      0.05 | Inf | 0.961
## -0.39     |    1.29e-03 | 0.02 | [-0.05, 0.05] |      0.05 | Inf | 0.957
## -0.36     |    1.37e-03 | 0.02 | [-0.05, 0.05] |      0.06 | Inf | 0.954
## -0.34     |    1.45e-03 | 0.02 | [-0.04, 0.05] |      0.06 | Inf | 0.951
## -0.31     |    1.53e-03 | 0.02 | [-0.04, 0.05] |      0.07 | Inf | 0.947
## -0.28     |    1.62e-03 | 0.02 | [-0.04, 0.05] |      0.07 | Inf | 0.944
## -0.26     |    1.70e-03 | 0.02 | [-0.04, 0.05] |      0.08 | Inf | 0.940
## -0.23     |    1.78e-03 | 0.02 | [-0.04, 0.05] |      0.08 | Inf | 0.936
## -0.20     |    1.86e-03 | 0.02 | [-0.04, 0.05] |      0.08 | Inf | 0.933
## -0.18     |    1.95e-03 | 0.02 | [-0.04, 0.04] |      0.09 | Inf | 0.929
## -0.15     |    2.03e-03 | 0.02 | [-0.04, 0.04] |      0.09 | Inf | 0.925
## -0.12     |    2.11e-03 | 0.02 | [-0.04, 0.04] |      0.10 | Inf | 0.921
## -0.10     |    2.19e-03 | 0.02 | [-0.04, 0.04] |      0.10 | Inf | 0.917
## -0.07     |    2.28e-03 | 0.02 | [-0.04, 0.04] |      0.11 | Inf | 0.913
## -0.04     |    2.36e-03 | 0.02 | [-0.04, 0.04] |      0.11 | Inf | 0.909
## -0.02     |    2.44e-03 | 0.02 | [-0.04, 0.04] |      0.12 | Inf | 0.905
## 9.73e-03  |    2.52e-03 | 0.02 | [-0.04, 0.04] |      0.12 | Inf | 0.901
## 0.04      |    2.60e-03 | 0.02 | [-0.04, 0.04] |      0.13 | Inf | 0.897
## 0.06      |    2.69e-03 | 0.02 | [-0.04, 0.04] |      0.13 | Inf | 0.893
## 0.09      |    2.77e-03 | 0.02 | [-0.04, 0.04] |      0.14 | Inf | 0.889
## 0.12      |    2.85e-03 | 0.02 | [-0.04, 0.04] |      0.14 | Inf | 0.885
## 0.14      |    2.93e-03 | 0.02 | [-0.04, 0.04] |      0.15 | Inf | 0.882
## 0.17      |    3.02e-03 | 0.02 | [-0.04, 0.04] |      0.15 | Inf | 0.878
## 0.20      |    3.10e-03 | 0.02 | [-0.04, 0.04] |      0.16 | Inf | 0.874
## 0.22      |    3.18e-03 | 0.02 | [-0.03, 0.04] |      0.16 | Inf | 0.870
## 0.25      |    3.26e-03 | 0.02 | [-0.03, 0.04] |      0.17 | Inf | 0.866
## 0.28      |    3.35e-03 | 0.02 | [-0.03, 0.04] |      0.17 | Inf | 0.863
## 0.30      |    3.43e-03 | 0.02 | [-0.03, 0.04] |      0.18 | Inf | 0.859
## 0.33      |    3.51e-03 | 0.02 | [-0.03, 0.04] |      0.18 | Inf | 0.856
## 0.36      |    3.59e-03 | 0.02 | [-0.03, 0.04] |      0.19 | Inf | 0.852
## 0.38      |    3.68e-03 | 0.02 | [-0.03, 0.04] |      0.19 | Inf | 0.849
## 0.41      |    3.76e-03 | 0.02 | [-0.03, 0.04] |      0.19 | Inf | 0.846
## 0.44      |    3.84e-03 | 0.02 | [-0.03, 0.04] |      0.20 | Inf | 0.843
## 0.46      |    3.92e-03 | 0.02 | [-0.03, 0.04] |      0.20 | Inf | 0.840
## 0.49      |    4.00e-03 | 0.02 | [-0.03, 0.04] |      0.20 | Inf | 0.838
## 0.52      |    4.09e-03 | 0.02 | [-0.03, 0.04] |      0.21 | Inf | 0.835
## 0.54      |    4.17e-03 | 0.02 | [-0.03, 0.04] |      0.21 | Inf | 0.833
## 0.57      |    4.25e-03 | 0.02 | [-0.03, 0.04] |      0.21 | Inf | 0.830
## 0.60      |    4.33e-03 | 0.02 | [-0.03, 0.04] |      0.22 | Inf | 0.828
## 0.62      |    4.42e-03 | 0.02 | [-0.04, 0.04] |      0.22 | Inf | 0.826
## 0.65      |    4.50e-03 | 0.02 | [-0.04, 0.04] |      0.22 | Inf | 0.824
## 0.68      |    4.58e-03 | 0.02 | [-0.04, 0.04] |      0.22 | Inf | 0.823
## 0.70      |    4.66e-03 | 0.02 | [-0.04, 0.05] |      0.23 | Inf | 0.821
## 0.73      |    4.75e-03 | 0.02 | [-0.04, 0.05] |      0.23 | Inf | 0.819
## 0.76      |    4.83e-03 | 0.02 | [-0.04, 0.05] |      0.23 | Inf | 0.818
## 0.78      |    4.91e-03 | 0.02 | [-0.04, 0.05] |      0.23 | Inf | 0.817
## 0.81      |    4.99e-03 | 0.02 | [-0.04, 0.05] |      0.23 | Inf | 0.816
## 0.84      |    5.08e-03 | 0.02 | [-0.04, 0.05] |      0.23 | Inf | 0.815
## 0.86      |    5.16e-03 | 0.02 | [-0.04, 0.05] |      0.24 | Inf | 0.814
## 0.89      |    5.24e-03 | 0.02 | [-0.04, 0.05] |      0.24 | Inf | 0.813
## 0.92      |    5.32e-03 | 0.02 | [-0.04, 0.05] |      0.24 | Inf | 0.812
## 0.94      |    5.41e-03 | 0.02 | [-0.04, 0.05] |      0.24 | Inf | 0.812
## 0.97      |    5.49e-03 | 0.02 | [-0.04, 0.05] |      0.24 | Inf | 0.811
## 1.00      |    5.57e-03 | 0.02 | [-0.04, 0.05] |      0.24 | Inf | 0.811
## 1.02      |    5.65e-03 | 0.02 | [-0.04, 0.05] |      0.24 | Inf | 0.810
## 1.05      |    5.73e-03 | 0.02 | [-0.04, 0.05] |      0.24 | Inf | 0.810
## 1.08      |    5.82e-03 | 0.02 | [-0.04, 0.05] |      0.24 | Inf | 0.810
## 1.10      |    5.90e-03 | 0.02 | [-0.04, 0.05] |      0.24 | Inf | 0.809
## 1.13      |    5.98e-03 | 0.02 | [-0.04, 0.05] |      0.24 | Inf | 0.809
## 1.16      |    6.06e-03 | 0.03 | [-0.04, 0.06] |      0.24 | Inf | 0.809
## 1.18      |    6.15e-03 | 0.03 | [-0.04, 0.06] |      0.24 | Inf | 0.809
## 1.21      |    6.23e-03 | 0.03 | [-0.04, 0.06] |      0.24 | Inf | 0.809
## 1.24      |    6.31e-03 | 0.03 | [-0.04, 0.06] |      0.24 | Inf | 0.809
## 1.26      |    6.39e-03 | 0.03 | [-0.05, 0.06] |      0.24 | Inf | 0.809
## 1.29      |    6.48e-03 | 0.03 | [-0.05, 0.06] |      0.24 | Inf | 0.809
## 1.32      |    6.56e-03 | 0.03 | [-0.05, 0.06] |      0.24 | Inf | 0.810
## 1.34      |    6.64e-03 | 0.03 | [-0.05, 0.06] |      0.24 | Inf | 0.810
## 1.37      |    6.72e-03 | 0.03 | [-0.05, 0.06] |      0.24 | Inf | 0.810
## 1.40      |    6.81e-03 | 0.03 | [-0.05, 0.06] |      0.24 | Inf | 0.810
## 1.42      |    6.89e-03 | 0.03 | [-0.05, 0.06] |      0.24 | Inf | 0.811
## 1.45      |    6.97e-03 | 0.03 | [-0.05, 0.06] |      0.24 | Inf | 0.811
## 1.48      |    7.05e-03 | 0.03 | [-0.05, 0.06] |      0.24 | Inf | 0.811
## 1.50      |    7.13e-03 | 0.03 | [-0.05, 0.07] |      0.24 | Inf | 0.811
## 1.53      |    7.22e-03 | 0.03 | [-0.05, 0.07] |      0.24 | Inf | 0.812
## 1.56      |    7.30e-03 | 0.03 | [-0.05, 0.07] |      0.24 | Inf | 0.812
## 1.58      |    7.38e-03 | 0.03 | [-0.05, 0.07] |      0.24 | Inf | 0.813
## Marginal effects estimated for HomeEvic_22_R_Z
```

**\# Table Making Related to Behavioral Problems (Table 05)**

\# Table with Behavioral Problems Base Model
Behavior_base_table <- tbl_regression(Behavior_Base_Adjusted_Age_Interact, exponentiate = TRUE) %>% bold_p()

\# Table with Behavioral Problems Strict Model
Behavior_stringent_table <- tbl_regression(Behavior_Stringent_Adjusted_Age_Interact, exponentiate = TRUE)%>% bold_p()

\# Combining Behavioral Problems Tables
Behavior_combined_table<-tbl_merge(
 tbls = list(Behavior_base_table, Behavior_stringent_table),
 tab_spanner = c("**Base Model**", "**Stringent Model**")
)

Behavior_combined_table

|  | Base Model | | | Stringent Model | | |
| --- | --- | --- | --- | --- | --- | --- |
| Characteristic | OR*^1^* | 95% CI*^1^* | p-value | OR*^1^* | 95% CI*^1^* | p-value |
| Child's Race/Ethnicity |  |  |  |  |  |  |
| White, non-Hispanic | — | — |  | — | — |  |
| Hispanic | 0.84 | 0.75, 0.95 | 0.006 | 0.86 | 0.76, 0.97 | 0.016 |
| Black, non-Hispanic | 0.81 | 0.68, 0.96 | 0.018 | 0.92 | 0.76, 1.10 | 0.3 |
| Asian, non-Hispanic | 0.41 | 0.32, 0.53 | <0.001 | 0.52 | 0.41, 0.68 | <0.001 |
| Multi-racial, non-Hispanic | 0.96 | 0.83, 1.11 | 0.6 | 0.85 | 0.73, 0.99 | 0.035 |
| Family Structure |  |  |  |  |  |  |
| Two parents, currently married | — | — |  | — | — |  |
| Two parents, not currently married | 1.30 | 1.11, 1.53 | 0.001 | 0.85 | 0.72, 1.01 | 0.065 |
| Single parent (mother or father) | 1.52 | 1.37, 1.69 | <0.001 | 0.82 | 0.73, 0.92 | <0.001 |
| Grandparent household | 2.55 | 0.25, 26.2 | 0.4 | 1.79 | 0.13, 24.0 | 0.7 |
| Other family type | 9.44 | 5.24, 17.0 | <0.001 | 3.98 | 2.14, 7.42 | <0.001 |
| Highest Level of Education in Household |  |  |  |  |  |  |
| Less than high school | — | — |  | — | — |  |
| High school or GED | 1.05 | 0.79, 1.39 | 0.7 | 0.89 | 0.66, 1.19 | 0.4 |
| Some college or technical school | 1.14 | 0.86, 1.50 | 0.4 | 0.93 | 0.70, 1.24 | 0.6 |
| College degree or higher | 0.99 | 0.75, 1.31 | >0.9 | 0.95 | 0.71, 1.26 | 0.7 |
| Household Poverty Status |  |  |  |  |  |  |
| 0-99% FPL | — | — |  | — | — |  |
| 100-199% FPL | 0.94 | 0.82, 1.09 | 0.4 | 0.94 | 0.81, 1.09 | 0.4 |
| 200-399% FPL | 0.86 | 0.75, 0.99 | 0.032 | 0.92 | 0.80, 1.07 | 0.3 |
| 400% FPL or greater | 0.78 | 0.68, 0.91 | 0.001 | 1.01 | 0.86, 1.17 | >0.9 |
| Child's Sex Assigned at Birth |  |  |  |  |  |  |
| Male | — | — |  | — | — |  |
| Female | 0.41 | 0.37, 0.44 | <0.001 | 0.39 | 0.36, 0.43 | <0.001 |
| Child's Age in Years | 1.06 | 1.01, 1.11 | 0.016 | 0.94 | 0.90, 0.99 | 0.023 |
| Eviction Stress/Concern | 1.22 | 1.18, 1.26 | <0.001 | 1.00 | 0.96, 1.04 | >0.9 |
| Child's Age in Years * Eviction Stress/Concern | 1.00 | 0.96, 1.03 | 0.9 | 1.00 | 0.97, 1.04 | 0.9 |
| Places Lived, Last Year |  |  |  |  |  |  |
| 0-2 times |  |  |  | — | — |  |
| 3 or more times |  |  |  | 1.50 | 1.19, 1.89 | <0.001 |
| Mother's Mental health |  |  |  |  |  |  |
| Excellent or very good |  |  |  | — | — |  |
| Good |  |  |  | 1.91 | 1.74, 2.10 | <0.001 |
| Fair or poor |  |  |  | 2.53 | 2.23, 2.87 | <0.001 |
| Ever Homeless |  |  |  |  |  |  |
| No |  |  |  | — | — |  |
| Yes |  |  |  | 1.67 | 1.36, 2.05 | <0.001 |
| Low Birth Weight |  |  |  |  |  |  |
| No |  |  |  | — | — |  |
| Yes |  |  |  | 1.38 | 1.19, 1.61 | <0.001 |
| Born Premature |  |  |  |  |  |  |
| No |  |  |  | — | — |  |
| Yes |  |  |  | 1.14 | 1.00, 1.31 | 0.057 |
| Food Insecurity |  |  |  | 1.18 | 1.09, 1.28 | <0.001 |
| Adverse Childhood Experiences |  |  |  |  |  |  |
| 0 ACEs |  |  |  | — | — |  |
| 1 ACE |  |  |  | 2.00 | 1.79, 2.24 | <0.001 |
| 2+ ACEs |  |  |  | 3.81 | 3.38, 4.29 | <0.001 |
| *^1^*OR = Odds Ratio, CI = Confidence Interval | | | | | | |

\# Output as Word Doc
Behavior_combined_table %>%
 gtsummary::as_gt() %>%
 gt::gtsave("Table5.docx")

### R Markdown (Analytic Code) Related to Analyses to Data Imputation (Three Methods, Noted Above)

**# Load Libraries**

pacman::p_load(haven,tidyverse,sjPlot, ggstatsplot, interactions, ggstats, oddsratio, epiDisplay, Hmisc, gtsummary,easystats,lme4,broom,gt,ggpubr,gridExtra,grid,multiUS,missForest,doParallel, mice)

**\# Imputation with Predictive Mean Matching**

vars<-c('raceASIA_22_F_L','famstruct5_22_F_L','AdultEduc_22_F_L','povlev4_22_F_L','PlacesLived_22_R_F_L','MothPhyH_22_R_F_L','MotherMH_22_R_F_L','ACE2more11_22_R_F_L','sex_22_R_L','EverHomeless_22_R_F_L','LowBWght_22_R_F_L','BornPre_22_R_F_L','SC_AGE_YEARS_Z','HomeEvic_22_R_Z','FIPSST','FoodSit_22_R','depress_22_R_Alt','anxiety_22_R_Alt','ADHD_22_R_Alt','behavior_22_R_Alt')
\#
NSCH_Topical_DRC_CAHMI_IVs<-data.frame(NSCH_Topical_DRC_CAHMI[vars])

\# Actual Imputation Command
NSCH_Topical_DRC_CAHMI_IVs_imputed <- mice(NSCH_Topical_DRC_CAHMI_IVs,m=50,maxit=25,meth='pmm',seed=500)

\# Overwriting Imputed Dependent Variables
NSCH_Topical_DRC_CAHMI_KNN$depress_22_R_Alt<-NSCH_Topical_DRC_CAHMI$depress_22_R_Alt
NSCH_Topical_DRC_CAHMI_KNN$anxiety_22_R_Alt<-NSCH_Topical_DRC_CAHMI$anxiety_22_R_Alt
NSCH_Topical_DRC_CAHMI_KNN$ADHD_22_R_Alt<-NSCH_Topical_DRC_CAHMI$ADHD_22_R_Alt
NSCH_Topical_DRC_CAHMI_KNN$behavior_22_R_Alt<-NSCH_Topical_DRC_CAHMI$behavior_22_R_Alt

**\# Imputation with k-nearest neighbors**

\#
vars<-c('raceASIA_22_F_L','famstruct5_22_F_L','AdultEduc_22_F_L','povlev4_22_F_L','PlacesLived_22_R_F_L','MothPhyH_22_R_F_L','MotherMH_22_R_F_L','ACE2more11_22_R_F_L','sex_22_R_L','EverHomeless_22_R_F_L','LowBWght_22_R_F_L','BornPre_22_R_F_L','SC_AGE_YEARS_Z','HomeEvic_22_R_Z','FIPSST','FoodSit_22_R','depress_22_R_Alt','anxiety_22_R_Alt','ADHD_22_R_Alt','behavior_22_R_Alt')
\#
NSCH_Topical_DRC_CAHMI_for_impute<-data.frame(NSCH_Topical_DRC_CAHMI[vars])

\# Actual Imputation Command
NSCH_Topical_DRC_CAHMI_KNN<-KNNimp(NSCH_Topical_DRC_CAHMI_for_impute)

\# Overwriting Imputed Dependent Variables
NSCH_Topical_DRC_CAHMI_KNN$depress_22_R_Alt<-NSCH_Topical_DRC_CAHMI$depress_22_R_Alt
NSCH_Topical_DRC_CAHMI_KNN$anxiety_22_R_Alt<-NSCH_Topical_DRC_CAHMI$anxiety_22_R_Alt
NSCH_Topical_DRC_CAHMI_KNN$ADHD_22_R_Alt<-NSCH_Topical_DRC_CAHMI$ADHD_22_R_Alt
NSCH_Topical_DRC_CAHMI_KNN$behavior_22_R_Alt<-NSCH_Topical_DRC_CAHMI$behavior_22_R_Alt

**\# Imputation with Random Forest**

\#
vars<-c('raceASIA_22_F_L','famstruct5_22_F_L','AdultEduc_22_F_L','povlev4_22_F_L','PlacesLived_22_R_F_L','MothPhyH_22_R_F_L','MotherMH_22_R_F_L','ACE2more11_22_R_F_L','sex_22_R_L','EverHomeless_22_R_F_L','LowBWght_22_R_F_L','BornPre_22_R_F_L','SC_AGE_YEARS_Z','HomeEvic_22_R_Z','FIPSST','FoodSit_22_R','depress_22_R_Alt','anxiety_22_R_Alt','ADHD_22_R_Alt','behavior_22_R_Alt')
\#
NSCH_Topical_DRC_CAHMI_for_impute<-data.frame(NSCH_Topical_DRC_CAHMI[vars])
\#
registerDoParallel(cores=2)
doRNG::registerDoRNG(seed = 123)

\# Actual Imputation Command
NSCH_Topical_DRC_CAHMI_for_impute_MissForest <- missForest(NSCH_Topical_DRC_CAHMI_for_impute, verbose = TRUE,maxiter = 10,parallelize = "forests")

\# Overwriting Imputed Dependent Variables
NSCH_Topical_DRC_CAHMI_for_impute_MissForest$depress_22_R_Alt<-NSCH_Topical_DRC_CAHMI$depress_22_R_Alt
NSCH_Topical_DRC_CAHMI_for_impute_MissForest$anxiety_22_R_Alt<-NSCH_Topical_DRC_CAHMI$anxiety_22_R_Alt
NSCH_Topical_DRC_CAHMI_for_impute_MissForest$ADHD_22_R_Alt<-NSCH_Topical_DRC_CAHMI$ADHD_22_R_Alt
NSCH_Topical_DRC_CAHMI_for_impute_MissForest$behavior_22_R_Alt<-NSCH_Topical_DRC_CAHMI$behavior_22_R_Alt
